# Multi-protein panel in pancreatic cyst fluid for improved risk stratification for pancreatic cancer

**DOI:** 10.1101/2025.09.17.25335852

**Authors:** Dimitrios Manolis, Darragh P. O’Brien, Benedikt M. Kessler, Eamon C. Faulkner, Pavlos Lykoudis, Farzana Haque, Sam Khulusi, Abdul Razack, Hemant M. Kocher, David K. Chang, Holger Kramer, Anthony Maraveyas, Leonid L. Nikitenko

## Abstract

Pancreatic cystic lesions (PCLs) are the sole radiologically recognisable and highly heterogeneous precursors of pancreatic cancer (PC). The malignant potential of PCLs is inferred from their types, as determined by empirical clinical practice guidelines; however, accurate risk stratification of patients preoperatively presents an unmet clinical need. We performed deep proteomic profiling of pancreatic cyst fluid (PCyF) and identified a first-of-its-kind multi-protein (n=89) panel termed “ASSIGN1” - Early diagnosis and detection of pAncreatic cySt malignancy SIGNature. ASSIGN1 was used for the development and validation of a support vector machine-based model for predicting malignant potential (based on malignancy risk score, zero to one) of individual PCLs using discovery/training and validation/test cohorts. The diagnostic accuracy of the model was evaluated based on histopathology of resected cysts using sensitivity, specificity and area under the receiver-operating-characteristic (AUROC) curve measures and compared to Fukuoka guidelines-based preoperative assessment. ASSIGN1-based malignancy risk score was a cyst type-independent and accurate (sensitivity=1.00, specificity=1.00 and AUROC=1.00) predictor of (i) pancreatic carcinoma and (ii) malignant potential of PCLs, which outperformed international consensus Fukuoka guidelines-based preoperative assessment (sensitivity=1.00; specificity=0.38; AUROC=0.71). Our findings demonstrated that ASSIGN1 holds promise to replace current preoperative laboratory tests, complement existing standard-of-care practices and improve preoperative diagnosis of PCLs and early detection of PC.

## 1. Introduction

The overall poor prognosis observed in pancreatic cancer (PC) is due to late detection and considerable challenges in diagnosis.^1^ Pancreatic cystic lesions (PCLs) are the sole radiologically recognisable and highly heterogeneous precursors of malignant transformation.^2^ However, there is a limited understanding of their biology that ultimately drives different clinical behaviours and outcomes.^2^ According to the 2017 International Association of Pancreatology (IAP; international consensus Fukuoka) guidelines, the current preoperative diagnosis of PCLs relies on imaging and clinical features.^3^ PCLs with “high-risk stigmata” (HRS) of malignancy require surgical resection, whilst the presence of ‘worrisome features’ (WF) is complemented by the performance of endoscopic ultrasound (EUS)-guided fine-needle aspiration (FNA) of pancreatic cyst fluid (PCyF).^3, 4^ Preoperative biochemical and cytological analyses of PCyF are used for the identification of mucinous cysts, which show higher potential for malignant transformation (malignant potential) compared to other PCL types.^5^ However, the insufficient depth in molecular profiling of PCyF offered by these laboratory tests (due to a limited number of biomarkers used) and the high heterogeneity of these cysts result in low accuracy of the prediction of malignant potential of PCLs, with precise diagnosis relying on cyst histopathology following surgery. Molecular profiling of liquid biopsies offers a unique opportunity for early detection and diagnosis of cancer.^6^ Multi-protein biomarker panels can reflect the variety of genomic and transcriptomic changes occurring in early-stage malignancy, allowing for accurate prediction of malignant potential and risk stratification.^7^ Label-free quantitation (LFQ) based mass spectrometry (MS) is a well-established tool to quantify proteomes of complex samples with high accuracy.^8^ Recent advances in LFQ data-independent acquisition (DIA) MS have enhanced throughput, proteome depth, reproducibility and sensitivity for unbiased identification of disease-associated biomarkers.^8^ Previous studies demonstrated some potential of MS-based analysis of PCyF proteome for detection of PC and risk stratification of specific (predominantly mucinous) cyst types.^9–13^ However, the utility of multi-protein panels in PCyF for predicting the malignant potential of PCLs independently of their type remains unexplored. In this study, we performed deep quantitative proteome profiling of PCyF to develop a multi-protein panel-based model for predicting the malignant potential of highly heterogeneous PCLs and compared its diagnostic accuracy to international consensus Fukuoka guidelines-based preoperative assessment.

## 2. Materials and methods

### 2.1 Clinical cohort - Study samples

PCyF samples (n=31; n=15 female; n=16 male; average age at diagnosis - 63 years; Supplementary Table S1) were collected in 2018-2020 for the ethically-approved Tumour Regulatory Molecules as Markers of Malignancy in Pancreatic Cystic Lesions study (TEM-PAC; ClinicalTrials.gov registration: NCT03536793; Integrated Research Application System (IRAS) Project ID: 236870; Research Ethics Committee Reference 18/LO/0736; 24.10.2018-28.02.2029; adopted by the National Institute for Health and Care Research (NIHR), portfolio ID 55297) in the Queens Centre for Oncology and Haematology, Castle Hill Hospital (Hull, UK) and made available for our single center retrospective study through IRAS Substantial Amendment 002 (15.01.2021). There was no perceived selection bias in patient recruitment. All individuals who agreed to participate provided informed consent. PCyF samples were obtained either preoperatively (n=22) using EUS or during surgery (n=9) by FNA along with corresponding follow-up details (clinical, imaging and diagnostic histopathological analysis; Supplementary Table S1). Cyst types and surgical pathology diagnoses were based on radiological, cytological, and biochemical characteristics/features according to the 2017 IAP guidelines.^3^ All PCLs were diagnosed as either “high-risk” (HRS or WF with suspicious, or positive for malignancy cytology and/or biochemistry; recommended for resection with subsequent histopathological analysis performed) or “low-risk” (WF with non-suspicious or negative for malignancy cytology and/or biochemistry; “presumed benign” and therefore unresected, from patients who either underwent follow-up surveillance or were discharged) according to Fukuoka criteria^3^ (Fig. 1 and Supplementary Table S1). Unresectable cysts due to advanced disease progression, along with lesions for which the type could not be assessed due to the presence of high malignant components, were also included in the study. Samples with a confirmed diagnosis of the specific grade (low, intermediate or high) of dysplasia or type of malignancy determined by an expert in gastrointestinal pathology were included in the study. After collection, all PCyF samples were anonymised and stored at −80°C until analysis.

**Fig. 1.**
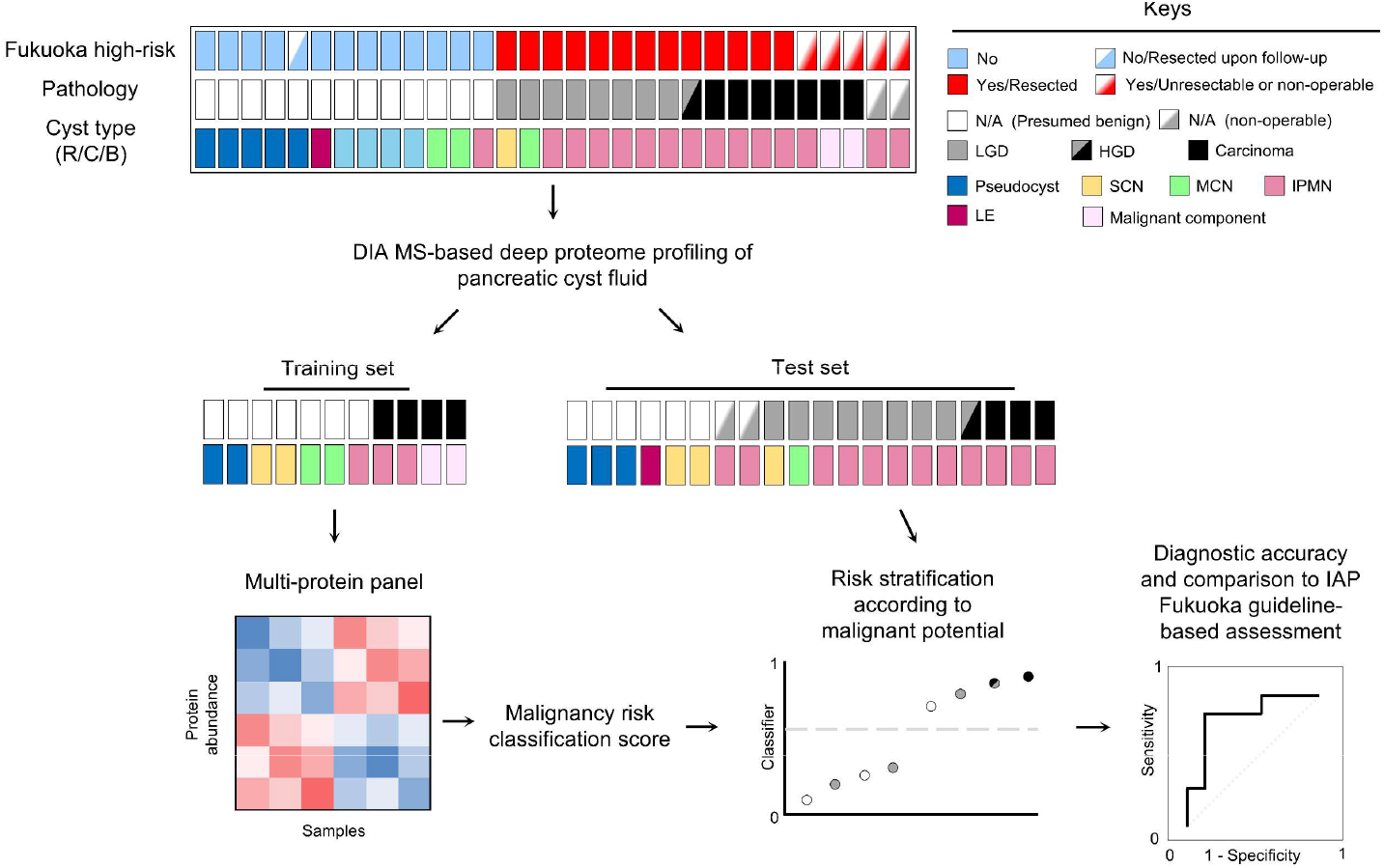
Summary of the analytical approach used to risk-stratify pancreatic cyst fluid samples according to malignant potential based on the signature of cyst fluid proteome. Diagnosis of pancreatic cysts was conducted based on radiological, cytological and biochemical characteristics (Supplementary Table S1; Materials and methods), according to the 2017 IAP Fukuoka guidelines.^3^ Pancreatic cystic lesions (PCLs) were grouped as either “high-risk” or “low-risk” according to Fukuoka criteria.^3^ MS-based proteomic profiling of PCyF samples (n=31) was performed in DIA mode. The MS-based proteomic dataset was divided into a training set and a test set (Materials and methods). A multi-panel of DEPs between groups of the training set was derived upon statistical analysis and inclusion of proteins with expression levels reduced in carcinomas and those specifically detected in all carcinomas but not detected in any presumed benign samples (Supplementary Figs. S1 and S2; Materials and methods). An SVM-based classifier was developed in Perseus software for cross-validation and prediction of the multi-protein panel.^22^ The SVM malignancy risk classification scores were utilised for assigning malignant potential to the cysts and for evaluating the diagnostic performance of the multi-protein panel-based model using the AUROC curve analysis. Abbreviations: R: radiology; C: cytopathology, B: biochemistry; IPMN: intraductal papillary mucinous neoplasm; MCN: mucinous cystic neoplasms; SCN: serous cystic neoplasm; LE: lymphoepithelial cyst; LGD: Low-grade dysplasia; HGD: High-grade dysplasia. N/A: Not available.

### 2.2 Proteomics sample processing

PCyF samples were lysed using sodium dodecyl sulfate (SDS) lysis buffer (5% SDS, 50 mM triethylammonium bicarbonate, TEAB, pH 7.5) before normalisation for protein amount using bicinchoninic acid assay-based quantification. Samples underwent an S-Trap™ micro spin column digestion workflow as described by the manufacturer.^14^ In brief, this included a reduction in 20 mM dithiothreitol for 30 min at room temperature (RT), followed by alkylation with 40 mM of iodoacetamide for 30 min at RT and in the dark. For S-Trap™ binding, proteins were acidified to 1.2% phosphoric acid and diluted 7-fold with 90% methanol/100 mM TEAB. Proteins were loaded onto the S-Trap™ cartridge and digested with a 1:25 ratio of trypsin: protein for 2 hours at 47°C. Peptides were eluted in 50% acetonitrile (ACN), 0.1% trifluoroacetic acid, dried down, and stored at −80°C until further use.

### 2.3 Nano-liquid chromatography-tandem mass spectrometry

Peptides were resuspended in 20mL 2% ACN, 0.1% formic acid and separated using a Dionex Ultimate 3000 nano-ultra high pressure reversed-phase chromatography system (RRID: SCR_019840) synced to an Orbitrap Ascend Tribrid Mass Spectrometer (Thermo Scientific), as previously described.^15^ Briefly, peptides were separated on an EASY-Spray PepMap RSLC C18 column (500 mm x 75 µm, 2 µm particle size; Thermo Scientific) over a 60 min gradient of 2-35% ACN in 5% dimethyl sulfoxide, 0.1% formic acid and at 250 nL/min. The column temperature was maintained at 50°C with the aid of a column oven. The mass spectrometer was operated in positive polarity mode with a capillary temperature of 275°C. DIA mode was utilised for automated switching between MS and MS/MS acquisition.^15^

### 2.4 Mass spectrometry data processing

Data acquired from nano liquid chromatography tandem MS (LC-MS/MS) were acquired and processed using Thermo Scientific Xcalibur (v 4.7; RRID: SCR_014593) and Thermo Scientific FreeStyle (v 1.8.65.0; RRID: SCR_022877). Nano LC-MS/MS data were analysed using DIA-NN (version 1.8; RRID: SCR_022865) with all settings as default.^16^ Specifically, we allowed for a maximum of one tryptic missed cleavage and fixed modifications of N-terminal M excision and carbamidomethylation of Cys residues. No variable modifications were selected. A *Homo sapiens* UniProt database (20,370 entries, retrieved on April 16, 2021) was used for the analysis. A default threshold of 1% false discovery rate (FDR) was used at the peptide and protein levels. We used the library-free mode of DIA-NN to generate precursor and corresponding fragment ions *in silico* from the UniProt database.^16^ The software also generates a library of decoy precursors (negative controls). Retention time alignment was performed using endogenous peptides, and peak scores were calculated by comparison of peak properties between observed and reference spectra. The ‘Match Between Runs’ feature was enabled to match high-resolution MS1 features between runs. LFQ intensity values were calculated upon normalisation of raw label-free quantitative nano-liquid chromatography-tandem MS data (peptide intensities) for each protein group using the MaxLFQ algorithm^17^ in DIA-NN^16^.

### 2.5 Statistical and bioinformatics analysis

For statistical and bioinformatics analysis, the total MS-based proteomic dataset was divided into a training set of 11 PCyF samples (seven presumed benign from four cyst types; pseudocysts, serous cystic neoplasms, mucinous cystic neoplasms, and intraductal papillary mucinous neoplasms; and four malignant from three types of carcinomas; pancreatic ductal adenocarcinoma, basaloid squamous cell carcinoma, and colloid carcinoma), and a test set of 20 anonymized samples (from six presumed benign, nine dysplastic, with respective pathology available, three carcinomas and, two non-operative cysts; Fig. 1 and Supplementary Table S1). All statistical and bioinformatics analyses were done using Perseus (v 2.0.10.0; RRID: SCR_015753).^18^ Protein abundance was calculated based on the LFQ of normalised spectrum intensity and log_2_-transformed. Before downstream analysis, potential contaminants were removed from the LFQ dataset, as previously described.^19^ The steps for deriving the final multi-protein panel are described in Supplementary Table S1. Briefly, the dataset was filtered based on valid LFQ values in both technical duplicates of at least one biological sample and the mean intensity used for normalisation (Z-score) and imputation. Missing values were imputed based on a normal distribution using default (width, 0.15; downshift, 1.8) settings in Perseus. Classical (highly abundant) plasma proteins,^20^ erythrocyte contamination-associated proteins,^21^ and immunoglobulins were excluded from the dataset, which contained at least 75% valid LFQ values within at least one group (Fukuoka high-risk or presumed benign groups) in the training set. For pairwise MS-based proteomic data comparisons, two-tailed Student’s t-tests with a permutation-based on FDR-adjusted p-value of 1% (<0.05; applying 250 randomisations) and S0 value of 2 were conducted to identify statistically significant differentially expressed proteins (DEPs), between Fukuoka high-risk and presumed benign groups of the training set, forming a generic multi-protein panel (Supplementary Figs. S1 and S2). Next, DEPs with reduced abundance in Fukuoka high-risk samples and those specifically detected in all Fukuoka high-risk samples, but not in any of the presumed benign samples, were included in the final multi-protein panel (Supplementary Fig. S1 and Supplementary Table S2).

### 2.6 Prediction of malignant potential using a multi-protein panel of pancreatic cyst fluid

To investigate the capacity of our 89-DEP panel ASSIGN1 to assign malignant potential of PCLs, we developed a classification model by using the training set and applied a supervised machine learning approach based on a built-in support vector machine (SVM) classifier in Perseus.^22^ An SVM-based classifier is considered an optimal (in effectiveness and reliability) solution for the classification of small sample size cohorts.^23^ Before classification (cross-validation and prediction), missing values were imputed based on a normal distribution using default (width, 0.15; downshift, 1.8) settings in Perseus. Firstly, the “classification parameter optimisation” function was used to obtain the C and sigma values (<5% classification error) for the classification model based on the Radial Basis Function kernel, utilising the protein expression data of ASSIGN1 in the training set. Next, a cross-validation procedure based on random sampling was applied (repeated 100 times) to avoid model overfitting. ASSIGN1 ensured <5% classification error. Next, the SVM-based “classification” function was used to assign a classification score (termed “malignancy risk classification score”) to each analysed PCyF sample in the training set. Scores obtained for individual samples were converted to values between zero (lowest) and one (highest). Next, the reproducibility and applicability of our model were determined using the “prediction” function to generate malignancy risk classification scores for all the samples in the test set. The accuracy/performance of the cross-validation and prediction runs for this classification model was determined by area under the receiver-operating-characteristic (AUROC) curve analysis. AUROC plots and optimal cut-off calculations were performed using the RStudio (v 4.2.0; RRID: SCR_000432) and R package pROC (RRID: SCR_024286).^24^ The optimal cutoff for calculating sensitivity and specificity was determined as a value corresponding to the maximum value of balanced accuracy, defined as the average of the sensitivity and specificity. The performance of ASSIGN1 in the prediction of malignant potential was evaluated based on histopathology of resected cysts and compared to the international consensus Fukuoka guidelines-based preoperative assessment (Supplementary Fig. S1 and Supplementary Table S1), with Fukuoka high-risk and presumed benign cases being assigned a “Fukuoka high-risk score” of one and zero, respectively.

### 2.7 Literature and database curation

Investigation of the association of ASSIGN1 members with PC was performed using a combination of literature and gene ontology (GO; RRID: SCR_002811) database.^25^ Curation (Supplementary Table S3). For literature curation, we manually curated information from published reports from the PubMed database (https://pubmed.ncbi.nlm.nih.gov/) and utilised non-MS-based methodology to study the expression and functional roles of these proteins in PC cells and tissues. For this, we integrated findings from existing publications and classified proteins by (i) cell types predicted to express them, (ii) biological process, and (iii) signalling pathway involved. For database curation, we analysed GO “Biological Process” and “Molecular Function” terms to assign protein families and cancer hallmarks^26^ to each protein (Supplementary Table S3).

### 2.8 Data visualisation

Principal component analysis, heat maps and volcano plots with Z-score values of log_2_ LFQ intensities were generated in Perseus.^18^ Dendrograms were built using Euclidean distance and default settings in Perseus. The R package Easyalluvial was used to generate alluvial plots (Easyalluvial version 0.3.2, https://rdrr.io/cran/easyalluvial).

## 3. Results

### 3.1 Design of the study

We performed molecular profiling and prediction of malignant potential for 31 PCyF samples from a heterogeneous cohort of PCLs by deep LFQ-based proteomics using DIA-MS and a machine-learning algorithm/classifier (Fig. 1). PCLs were grouped as either Fukuoka “high-risk” or “low-risk” according to international consensus Fukuoka guidelines (Fig. 1 and Supplementary Table S1; Materials and methods). We analysed all samples in a blinded manner and generated (i) individual comprehensive proteomic profiles for all PCyF samples, exposing the heterogeneity of PCLs, and (ii) a multi-protein panel, enabling the assignment/prediction of the malignant potential of these cysts (Fig. 1).

### 3.2 Deep quantitative proteomic profiling of PCyF

First, we used label-free quantitative MS to quantify the proteomes of 31 anonymised PCyF samples in a blinded fashion and with two technical duplicates (Fig. 2A). This analysis identified 3,461 proteins and demonstrated the variability in the number of proteins (ranging between 631 and 3,160) identified in PCyF from individual PCLs, independently of their post-operative diagnosis or type. Raw nano-liquid chromatography-tandem MS data (identified peptide intensities) were normalised across samples, and the Z-score values of log_2_ LFQ intensities (average of two technical replicates) were used for downstream bioinformatics and statistical analysis (Fig. 2B; Materials and methods). We then investigated whether unsupervised analysis could uncover differences between proteomic profiles of individual PCyF samples and map PCLs according to their (i) type and (ii) malignancy state. Principal component analysis (PCA) of normalised MS data for all identified proteins demonstrated that deep LFQ proteomic profiling of PCyF (i) maps presumed benign cases away from malignant PCLs and (ii) reveals a variable distribution of dysplastic PCLs according to individual comprehensive proteomes and in line with their expected heterogeneity (Fig. 2C).

**Fig. 2.**
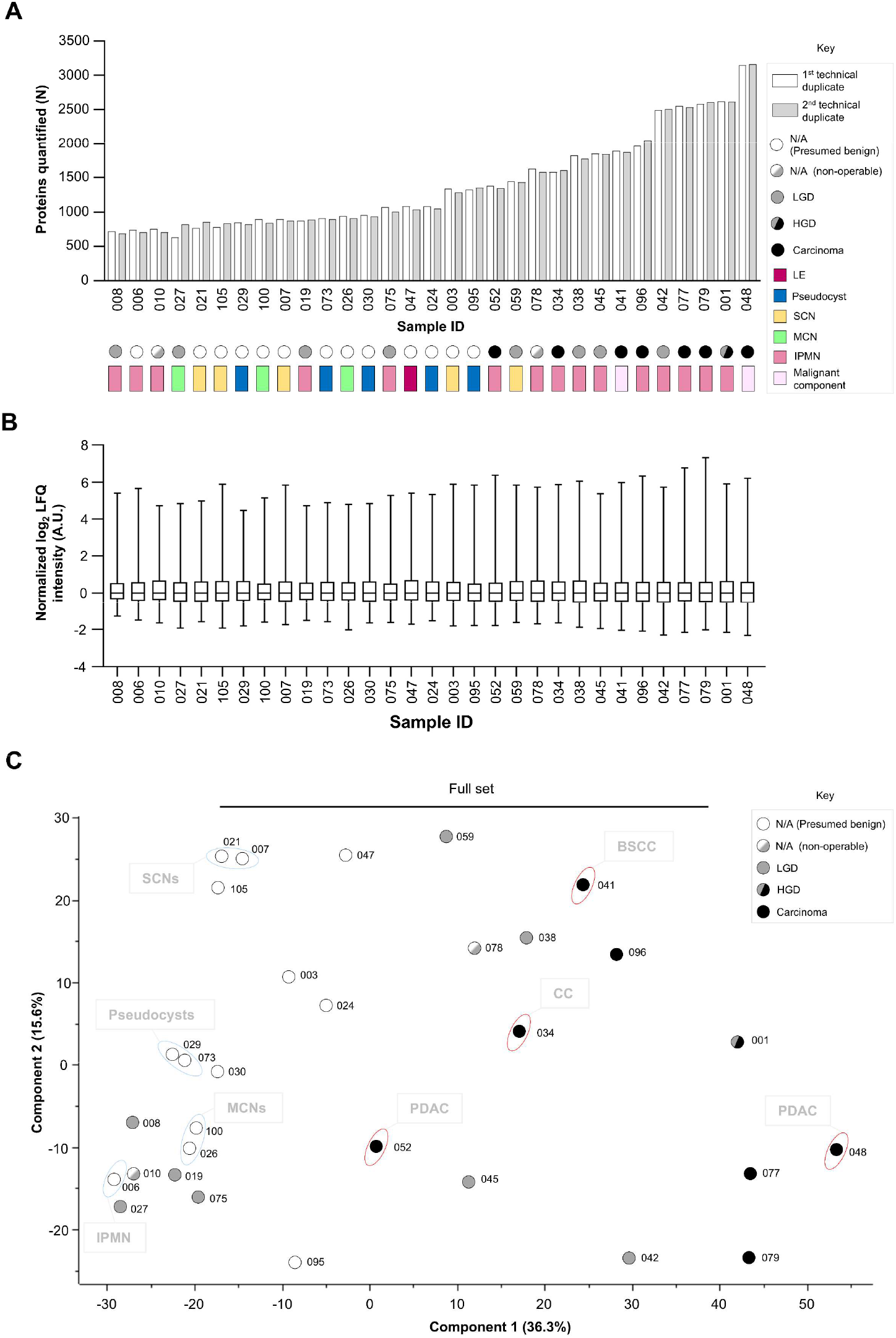
Comprehensive mapping of various pancreatic cysts based on deep proteomic profiling of cyst fluid. **A**. Bar chart shows the number of identified proteins in 31 pancreatic cyst fluid (PCyF) samples from PCLs (Fig. 1; Supplementary Table S1) in technical duplicates (Key) using LFQ-based nano liquid chromatography-tandem mass spectrometry (LC-MS/MS). Corresponding cyst type (rectangular), along with resection and pathology status (circle), are indicated in the Key. **B**. Box and whisker plots of Z-score values of normalised log_2_ LFQ intensity values calculated upon normalisation of raw LFQ nano LC-MS/MS data (peptide intensities) for each protein identified in PCyF samples (Materials and methods). **C**. Principal component analysis (PCA) of PCyF proteomic data presented in a 2D graph of principal components −1 and −2 (PC1 and PC2). PCA was performed using the protein abundance values, represented by Z-score values of normalised log_2_-transformed LFQ intensity, and assigned the largest variance to the difference between 31 PCyF samples acquired from various cyst types (Component 1, 36.3%). Each circle represents the mean LFQ intensity per sample based on two technical replicates. For the training set, four types of presumed benign (blue ellipsoids) and three types of carcinomas (red ellipsoids) samples were used (Fig. 1; Materials and methods). Corresponding resection and pathology status (circle) for each cyst are indicated in the Key. (A and C) Abbreviations: IPMN: intraductal papillary mucinous neoplasm; MCN: mucinous cystic neoplasms; SCN: serous cystic neoplasm; LE: lymphoepithelial cyst; LGD: Low-grade dysplasia; HGD: High-grade dysplasia. PDAC: Pancreatic ductal adenocarcinoma; BSCC: Basaloid squamous cell carcinoma; CC: Colloid carcinoma. N/A: Not available.

### 3.3 Identification of multi-protein panel ASSIGN1 for predicting the malignant potential of heterogeneous PCLs

We next divided samples into training (discovery) and test (validation) sets (Fig. 1). For the training set, we selected PCyF samples from seven presumed benign (four cyst types; Fig. 2C) and four Fukuoka high-risk (three carcinoma types; Fig. 2C) cysts that reflect the high heterogeneity of commonly detected PCLs in the clinic. We used the training set to decipher a generic multi-protein panel that is associated with PC due to the significant differences in the PCyF proteomes between presumed benign and carcinoma-associated cysts (Supplementary Fig. S1). More specifically, statistical analysis revealed a panel of 377 differentially expressed proteins (DEPs) between Fukuoka high-risk and presumed benign groups (Supplementary Fig. S2a and Supplementary Table S2). This generic multi-protein panel efficiently discriminated between two groups of the training set (Supplementary Fig. S2b). Next, DEPs specifically detected in all Fukuoka high-risk samples, but not in any of the presumed benign samples, and also those with reduced abundance in Fukuoka high-risk samples (n=86 and n=3, respectively), were included in the final multi-protein panel (89 DEPs; Fig. 3A; Supplementary Fig. S1 and Table S2). The 89-DEP panel was termed “Early diagnosis and detection of pAncreatic cySt malignancy SIGNature 1” (ASSIGN1). Based on literature and database curation, ASSIGN1 members are either known to be associated with specific protein families and hallmarks of cancer or represent novel biomarkers for PC (Supplementary Fig. S3).

**Fig. 3.**
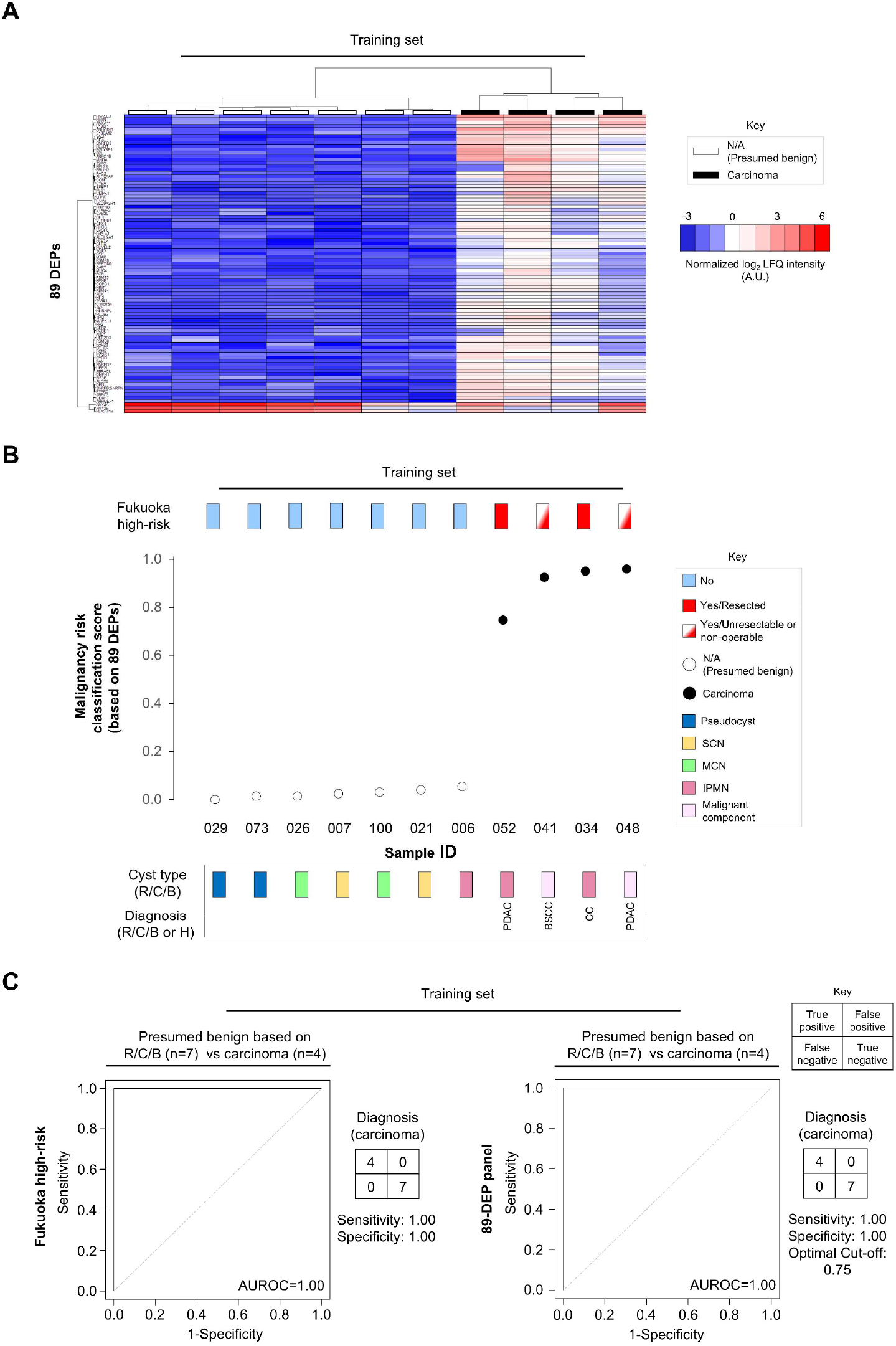
Malignancy risk scoring of pancreatic cystic lesions based on the multi-protein panel ASSIGN1 identified in pancreatic cystic fluid. **A**. Clustered heatmap of 89 DEPs between presumed benign and carcinoma-associated PCLs identified in pancreatic cyst fluid samples of the training set (Supplementary Fig. S2 and Table S2). Data is presented as Z-scores of log_2_ label-free LFQ intensity values, which represent protein abundance according to the colour scale bar at the right of the figure (red-high; low-blue). The dendrograms were built using Euclidean distance in Perseus software. **B**. Dot plot of malignancy classification score distribution for heterogeneous PCLs from the training set with corresponding cyst type, resection, and pathology status indicated (Key). The malignancy score (zero: lowest; one: highest) for each sample was assigned by a support vector machine classifier using an 89-DEP panel (Fig. 3A; Materials and methods). **C**. AUROC curve, sensitivity and specificity were used to measure the diagnostic performance (or accuracy) for predicting the malignant potential of PCLs based on the histopathology of resected cysts and compared between Fukuoka guidelines-based preoperative assessment (left graph) and 89-DEP panel-based malignancy risk classification score (right graph). Diagnostic accuracy was assigned in terms of specificity ([True Negatives] / [True Negatives + False Positives]), sensitivity ([True Positives] / [True Positives + False Negatives]), and AUROC values. (B-C) Abbreviations: R: radiology; C: cytopathology, B: biochemistry; IPMN: intraductal papillary mucinous neoplasm; MCN: mucinous cystic neoplasms; SCN: serous cystic neoplasm; PDAC: Pancreatic ductal adenocarcinoma; BSCC: Basaloid squamous cell carcinoma; CC: Colloid carcinoma; N/A: Not available.

### 3.4 Malignancy risk score based on ASSIGN1 is an independent predictor of pancreatic carcinoma

We then used ASSIGN1 to develop a support vector machine (SVM)-based model, which classified the Fukuoka presumed benign and Fukuoka high-risk samples within the selected training set by assigning a malignancy risk score (Fig. 3B). The diagnostic accuracy of ASSIGN1 in the prediction of malignant potential of heterogeneous PCLs was evaluated based on the histopathology of resected cysts and compared to Fukuoka guidelines-based preoperative assessment using the training set (Figs. 1 and 2B). The sensitivity and specificity of ASSIGN1 were optimal and similar to Fukuoka high-risk preoperative assessment/diagnosis (area under the receiver operating characteristic, AUROC=1 for both; Fig. 3C), demonstrating the ability of our multi-protein panel to comply with current standard-of-care diagnostic modalities utilised for the management of PCLs.

### 3.5 ASSIGN1-based prediction of malignant potential and risk stratification of heterogeneous PCLs

We evaluated the capacity of ASSIGN1 to unbiasedly (independent of cyst type) risk stratify cysts (presumed benign or diagnosed post-operatively with dysplasia, or malignancy; Supplementary Table S1; Materials and methods) within the test set (Fig. 4A). This analysis demonstrated that carcinoma-associated lesions had higher malignancy risk classification scores compared to others (Fig. 4A). Furthermore, the ASSIGN1-based model risk stratified neoplastic PCLs with the same post-operative diagnosis (low-grade dysplasia) and clinical management (resection/surgery) into two distinct cohorts (Fig. 4A) and also differentiated presumed benign cases (Fig. 4A).

**Fig. 4.**
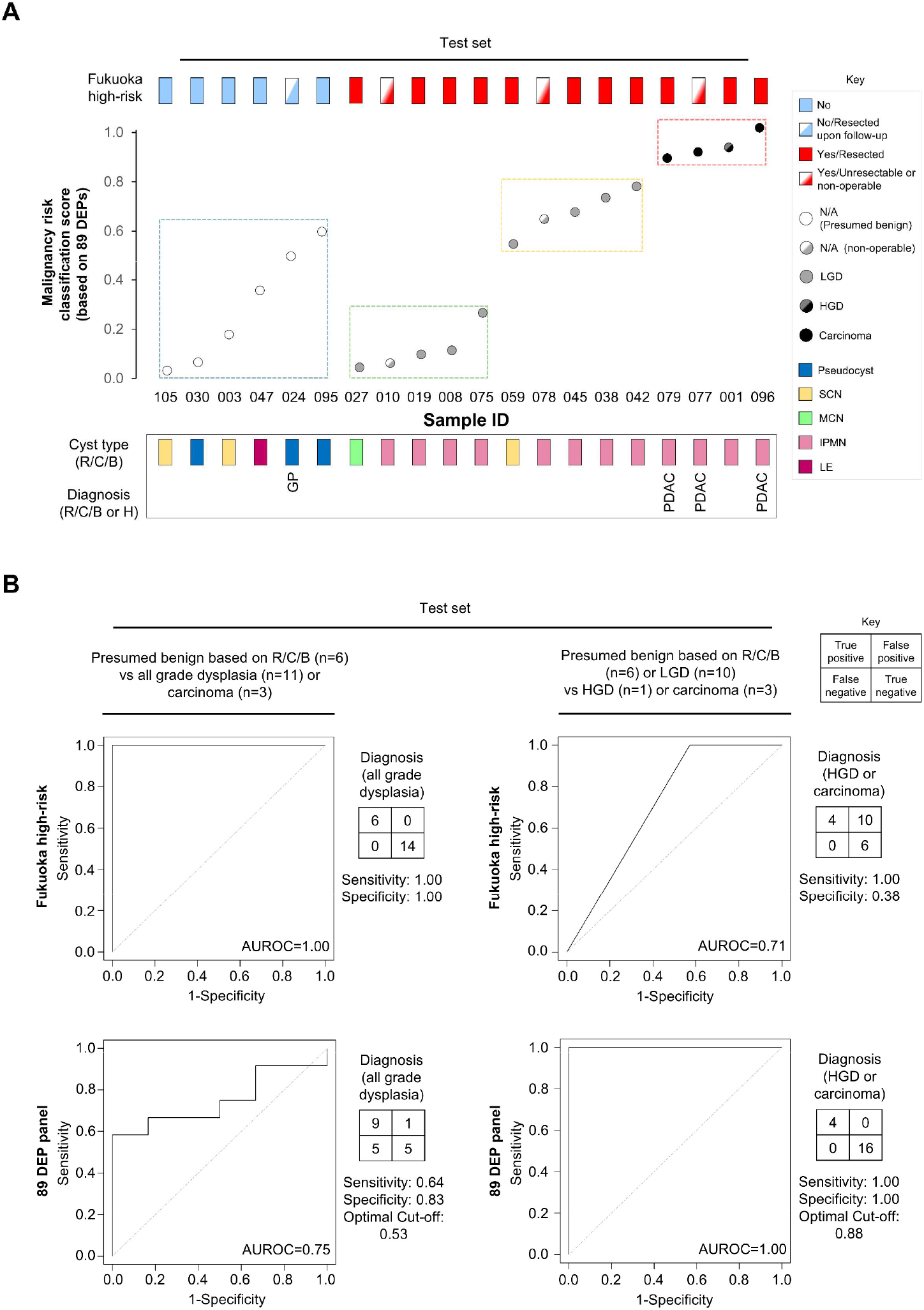
Prediction of malignant potential of pancreatic cystic lesions based on multi-protein panel ASSIGN1 in pancreatic cyst fluid and comparison to Fukuoka guidelines-based diagnosis. **A**. Dot plot of malignancy classification score distribution for heterogeneous PCLs from the test set with corresponding cyst type, resection, and pathology status indicated (Key). The malignancy risk classification score (zero: lowest; one: highest) for each sample was assigned by a support vector machine classifier using the 89-DEP panel (Materials and methods). Cancerous lesions had higher malignancy risk classification scores compared to others (red dotted box). The ASSIGN1-based model showed the potential to risk-stratify neoplastic PCLs with the same post-operative diagnosis (low-grade dysplasia) and clinical management (resection/surgery) into two distinct cohorts (yellow and green dotted boxes). Also, presumed benign cases had variable malignancy risk classification scores (blue dotted box). **B**. The AUROC curve, sensitivity and specificity were used to measure the diagnostic performance (accuracy) from predicting dysplasia (left graphs), and high-grade dysplasia or carcinoma (right panels) of PCLs based on histopathology of resected cysts and compared between preoperative assessments according to Fukuoka guidelines (Fukuoka high-risk; top graphs), and ASSIGN1-based malignancy risk classification score (89 DEP; bottom graphs). Diagnostic accuracy was assigned in terms of specificity ([True Negatives] / [True Negatives + False Positives]) and sensitivity ([True Positives] / [True Positives + False Negatives]), and AUROC values. (A-B) Abbreviations: R: radiology; C: cytopathology, B: biochemistry; IPMN: intraductal papillary mucinous neoplasm; MCN: mucinous cystic neoplasms; SCN: serous cystic neoplasm; LE: lymphoepithelial cyst; PDAC: Pancreatic ductal adenocarcinoma; BSCC: Basaloid squamous cell carcinoma; CC: Colloid carcinoma. GP: Groove pancreatitis; LGD: Low-grade dysplasia; HGD: High-grade dysplasia. N/A: Not available.

### 3.6 Comparison of ASSIGN1-based malignancy risk stratification to international consensus Fukuoka guidelines-based preoperative assessment

The diagnostic accuracy of ASSIGN1-based model was then evaluated based on histopathology of resected cysts and compared to international consensus Fukuoka guidelines-based preoperative assessment using the test set (Fig. 4B). Interestingly, ASSIGN1 had a lower efficacy when discriminating presumed benign from dysplastic (all grade) or malignant/carcinoma cases (AUROC=0.75; Fig. 4B, bottom left), as compared to Fukuoka guidelines-based preoperative assessment/diagnosis (AUROC=1.00; Fig. 4B, top left). This important finding revealed that, according to ASSIGN1, both presumed benign and low-grade dysplastic PCLs are heterogeneous and could be further risk-stratified preoperatively. Further analysis demonstrated that the ASSIGN1-based malignancy risk classification score was superior in assigning high-risk malignant potential to high-grade dysplastic cysts and carcinomas (AUROC=1.00; Fig. 4B, bottom right) over Fukuoka guidelines (AUROC=0.71 and optimal malignancy risk classification score cutoff of 0.53; Fig. 4B, top right). Furthermore, the ASSIGN1-based malignancy risk score demonstrated its capacity to risk-stratify heterogeneous PCLs from both resected and presumed benign cysts (Fig. 4A).

## 4. Discussion

Without a breakthrough in early detection, PC is projected to become the second leading cause of cancer-related mortality before 2040.^27^ Malignant progression in PCLs is a multistep and prolonged process, providing a large window of opportunity for early diagnosis.^1^ The routine monitoring of patients with PCLs under surveillance and the increased number of incidentally detected cysts in asymptomatic individuals highlight the urgent need to balance the prevention of PC with the risk of unnecessary surgery.^5, 28, 29^ The levels of mucin, carcinoembryonic antigen and amylase in PCyF are useful for diagnosing mucinous cysts, which are associated with a higher malignant potential than other types but often have variable post-operative diagnosis. Whilst cytopathological evaluation can be conclusive for detecting malignancy, the whole preoperative diagnostic ‘package’ has low sensitivity for risk-stratifying PCLs. A combination of imaging with a comprehensive molecular test that considers the high cyst heterogeneity could accurately predict malignant potential and improve the clinical management of these precursor lesions of PC. Our findings demonstrate that deep LFQ proteomic profiling of PCyF and multi-protein panel ASSIGN1 have the potential to complement the deployment of imaging and EUS resources for predicting malignant potential of PCLs, thereby reducing the current reliance on resection for precise diagnosis upon histological assessment.

The utilisation of deep proteomics approaches and multi-biomarker panels offers a comprehensive molecular profiling of cancer subtypes and thus a better diagnostic tool for precision medicine, compared to single/individual-biomarker approaches and other diagnostic modalities.^7^ To date, despite the number of MS-based studies of the PCyF proteome, the concept of a multi-protein panel in PCyF for preoperative assignment of malignant potential and risk stratification of PCLs using an unbiased (cyst type-independent) approach remains insufficiently explored. Previous studies analysed the utility of individual proteins or small panels of proteins (n=2-4) in PCyF as biomarkers for detecting malignancy and/or distinguishing the grade of mucinous PCLs.^9–13^ These studies either (i) used only samples collected upon resection (not preoperatively by EUS-guided aspiration), (ii) had low proteomic coverage of PCyF, (iii) focused only on the analysis of PCyF from mucinous cysts, (iv) provided unclear histopathological data, (v) did not validate findings using an independent cohort and, (vi) did not evaluate the diagnostic accuracy of the proposed biomarkers in relation to current guidelines.

Our data indicate that a multi-protein panel, ASSIGN1, derived from deep quantitative proteome profiling of PCyF and consisting of 89 DEPs, can be used preoperatively to successfully predict malignant potential, and risk-stratify a wide range of PCLs with varying histopathology, i.e. cyst-type independently, thus significantly advancing international consensus Fukuoka guidelines-based preoperative assessment. Our results show that ASSIGN1 is capable of: (i) detecting high-grade dysplasia and malignant cases with high efficacy, (ii) risk-stratifying neoplastic PCLs with the same type with inferred malignant potential and post-operative diagnosis (low-grade dysplasia) into two cohorts, and (iii) differentiating presumed benign cases (Fig. 4A). Our findings demonstrate that ASSIGN1-based malignancy risk score can be used as an independent predictor of (i) pancreatic carcinoma and (ii) malignant potential of PCLs, outperforming international consensus Fukuoka guidelines-based preoperative assessment of these precursor lesions of PC. These unprecedented findings and discoveries suggest that our approach and diagnostic tool could improve clinical decision-making by identifying lesions with (i) high malignant potential that should be surgically removed and (ii) lower malignant potential (independent of their type) for reducing unnecessary highly morbid surgery or excessive surveillance.

Critically, the diagnostic accuracy of ASSIGN1 is dependent on 89 DEPs, accounting for the heterogeneity of PCL types^5^ and the complexity of PCL microenvironment, whilst also aligning with the relevant for PC biology top five hallmarks of cancer (tumour-promoting inflammation, unlocking phenotypic plasticity, sustaining proliferative signalling, activating invasion and metastasis, and genome instability and mutation).^30^ This finding highlights the multidimensional nature of ASSIGN1 and emphasises the need for further investigation into the roles of its members in pancreatic carcinogenesis. In particular, the identification of 24 proteins belonging to ASSIGN1 and previously not reported to have an association with PC could inform future studies into their novel roles in PC pathogenesis. Moreover, the discovery of ASSIGN1 proteins in PCyF could guide the search for these (PCL microenvironment-derived and PC-associated) biomarkers of malignant transformation in systemic circulation/blood or urine.

In summary, we presented a proof-of-concept that a multi-protein panel can be used to effectively predict the malignant potential of PCLs independently of cyst type. Our study demonstrates the clinical application of deep quantitative LFQ proteome profiling of PCyF for risk stratification of PCLs, contributing to early diagnosis and guiding the management of PC as a highly heterogeneous disease. After assessing the utility in large prospective cohorts and combined with current guidelines, our approach could provide a unique and valuable diagnostic and improved standard-of-care tool for early detection of PC and improving survival, enabling to avoid unnecessary surgery or excessive surveillance with detrimental effects on patients, and reducing healthcare resource consumption. Thus, our discovery offers a unique and advanced diagnostic tool that could be developed into a point-of-care test that complements current standard-of-care practices with a clear potential for improving cancer patient care.

## CRediT authorship contribution statement

D. Manolis: Investigation, formal analysis, validation, data curation, software, visualisation, writing–original draft, writing–review and editing. D. P. O’Brien: Conceptualisation, investigation, methodology, funding acquisition, resources, formal analysis, data curation, software, writing–original draft, writing–review and editing. B. M. Kessler: Conceptualisation, methodology, funding acquisition, resources, formal analysis, writing–original draft, writing–review and editing. E. C. Faulkner: Methodology, software, visualisation, resources, writing– original draft, writing–review and editing. P. Lykoudis: Methodology, conceptualisation, resources, writing– original draft, writing–review and editing. F. Haque: Methodology, conceptualisation, resources, writing–original draft, writing–review and editing. S. Khulusi: Methodology, conceptualisation, resources, writing–original draft, writing–review and editing. A. Razack: Methodology, conceptualisation, resources, writing–original draft, writing–review and editing. H. M. Kocher: Methodology, conceptualisation, resources, writing–original draft, writing–review and editing. D. K. Chang: Methodology, conceptualisation, resources, writing–original draft, writing–review and editing. H. Kramer: Conceptualisation, methodology, resources, investigation, formal analysis, writing–original draft, writing–review and editing. A. Maraveyas: Project administration, supervision, conceptualisation, methodology, resources, funding acquisition, writing–original draft, writing–review and editing. L.L. Nikitenko: Project administration, supervision, conceptualisation, funding acquisition, resources, methodology, formal analysis, data curation, writing–original draft, writing–review and editing.

## Data Availability

All MS raw files were deposited in the PRoteomics IDEntifications (PRIDE; RRID: SCR_003411) Archive31 under the unique identifier PXD045289.

https://www.ebi.ac.uk/pride/archive

## Data availability

All MS raw files were deposited in the PRoteomics IDEntifications (PRIDE; RRID: SCR_003411) Archive^31^ under the unique identifier PXD045289.

## Declaration of Competing Interest

D. Manolis, D. P. O’Brien, B. M. Kessler, D. K. Chang, A. Maraveyas and L.L. Nikitenko have a patent pending to the University of Hull. Other authors declare that they have no known competing financial interests or personal relationships that could have appeared to influence the work reported in this paper.

## Acknowledgments

The project study was supported in part by the Cancer Research UK Early Detection and Diagnosis Primer Award (grant EDDPMA-May22/100018 EARLY DIAPAC: EARLY DIAgnosis of Pancreatic Cancer combined proteomics and genomics testing of pancreatic cyst fluid to LN, HK, DC, DO’B, BMK and AM) and the Working Independently to Support Hull Hospitals (WISHH) Charity (grant: Quantitative proteomic analysis of pancreatic cyst fluid for early detection of cancer using University of Hull HPC Viper to LN, DM and AM). We are indebted to Dr Aseem Allam and his family for the support and funding of the umbrella biobank study (TEM-PAC; ClinicalTrials.gov registration: NCT03536793, adopted by the NIHR; Materials and methods), which has generated the clinical material accessed by the investigators of the EARLY DIAPAC consortium. We would like to thank all participants in this study, the endoscopy team and the research nurses from Castle Hill Hospital for the collection of the samples, Chris Collins and the support team of the High-Performance Computing Facility from the University of Hull, the members of the Discovery Proteomics Facility from University of Oxford for expert help with the acquisition of mass spectrometry data, and Professor Adrian Harris (University of Oxford), Dr. Alexandre Akoulitchev (Oxford BioDynamics), Dr. Markus Queisser (GlaxoSmithKline), Dr Nischalan Pillay (University College London), Professor Tatsuo Shimosawa (University of Tokyo), Drs Camille Ettelaie, Adenike Adekeye, Shirin Hasan and Sophie Featherby (all – from the University of Hull) for their advice and feedback, and Dr Laura Broughton, James Illingworth and David Hare for their tireless managerial support.

## Notes

### Author Declarations

PCyF samples (n=31; n=15 female; n=16 male; average age at diagnosis - 63 years; Supplementary Table S1) were collected in 2018-2020 for the ethically-approved Tumour Regulatory Molecules as Markers of Malignancy in Pancreatic Cystic Lesions study (TEM-PAC; ClinicalTrials.gov registration: NCT03536793; Integrated Research Application System (IRAS) Project ID: 236870; Research Ethics Committee Reference 18/LO/0736; 24.10.2018-28.02.2029; adopted by the National Institute for Health and Care Research (NIHR), portfolio ID 55297) in the Queens Centre for Oncology and Haematology, Castle Hill Hospital (Hull, UK) and made available for our single center retrospective study through IRAS Substantial Amendment 002 (15.01.2021). There was no perceived selection bias in patient recruitment. All individuals who agreed to participate provided informed consent. PCyF samples were obtained either preoperatively (n=22) using EUS or during surgery (n=9) by FNA along with corresponding follow-up details (clinical, imaging and diagnostic histopathological analysis; Supplementary Table S1).

