## Supplementary Data for "Multi-protein panel in pancreatic cyst fluid for improved risk stratification for pancreatic cancer"

**Contents:**

Supplementary Figs S1-3

Supplementary Tables S1-3

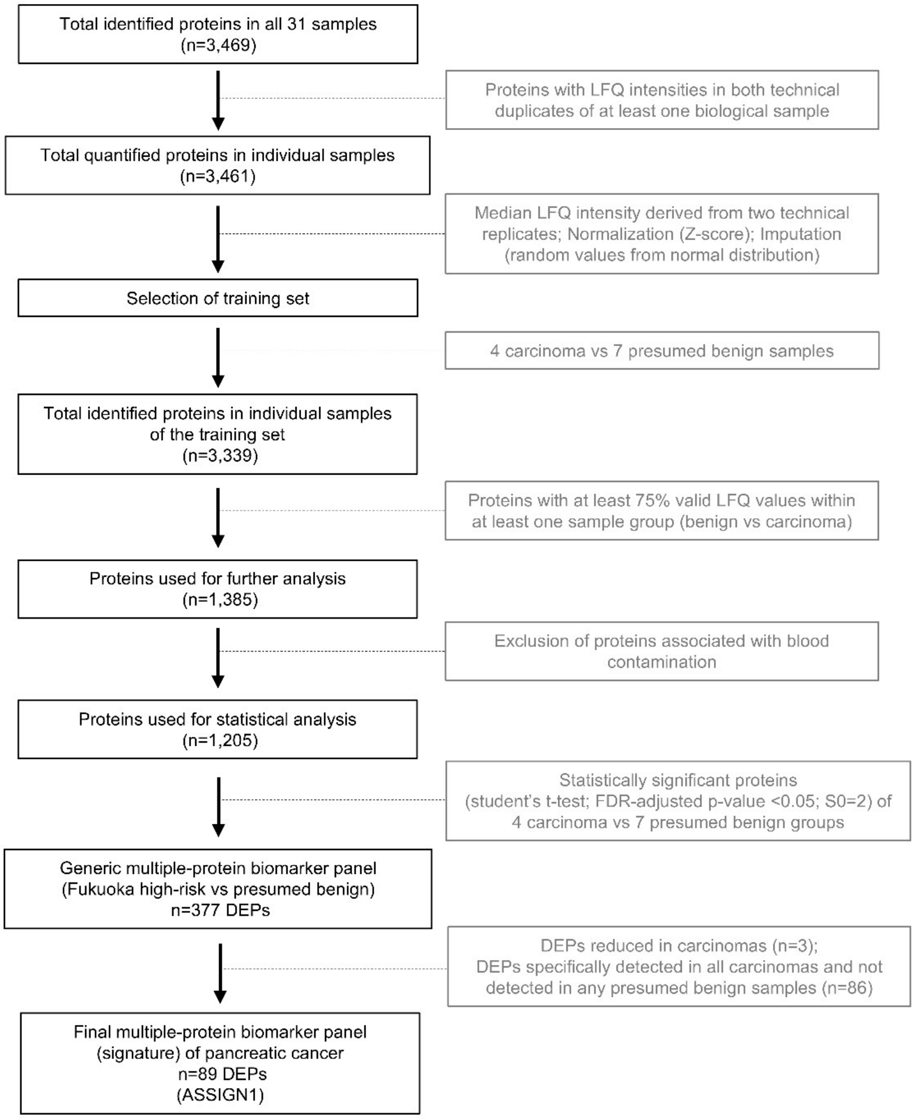

Supplementary Fig S1. Flowchart for the discovery of a multi-protein panel associated with pancreatic cancer based on cyst fluid proteome. A multi-protein panel associated with pancreatic cancer was identified in pancreatic cyst fluid (PCyF) by following an eight-step process. More specifically, of the 3,469 proteins identified, 3,461 were quantified in all individual PCyF samples (n=31; Fig. 1; Materials and methods) and had valid label-free quantitation (LFQ) intensity values in both technical replicates (Fig. 2a; Materials and methods) of any biological sample. Next, 1,385 proteins with at least 75% valid LFQ values within at least one group of the training set (seven presumed benign and four carcinoma-associated samples; Figs. 1 and 3; Materials and methods) were selected for downstream analysis. After removal of proteins identified in abundance in human blood (Materials and methods), 1,205 remaining proteins were used for statistical analysis and 377 differentially expressed proteins (DEPs) were identified (two-tailed Student's t-test; FDR-adjusted p-value <0.05; S0=2.00) between carcinoma and presumed benign cysts in the training set (Figs. 1 and 3; Materials and methods) forming a generic multi-protein panel (Fig. S2 and Table S2). The expression levels of 374 DEPs were increased and 3 were decreased in carcinomas. DEPs reduced in carcinomas (n=3) and DEPs specifically detected in all carcinomas and not detected in any presumed benign samples (n=86) were included in the final multi-protein (n=89) panel, termed "Early diagnosis and detection of pAncreatic cySt malignancy SIGNature 1" (ASSIGN1).

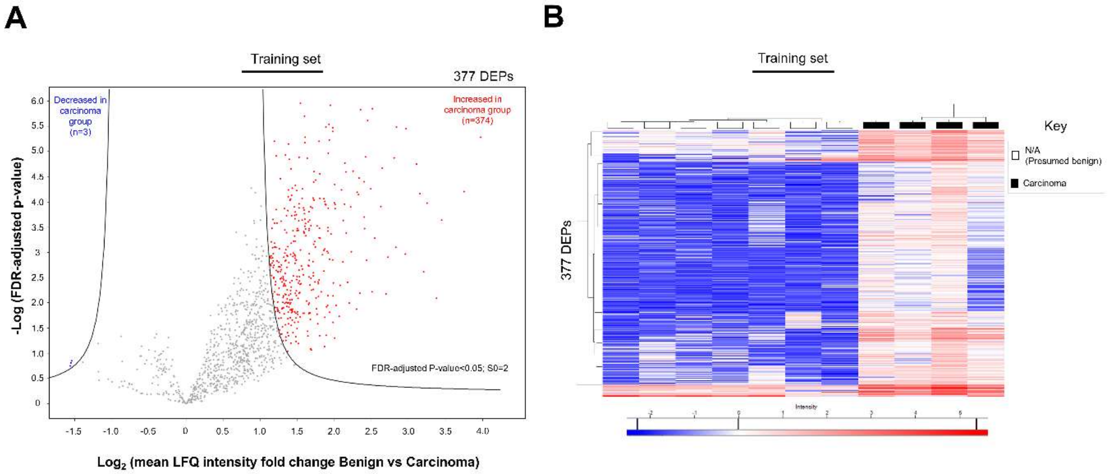

Supplementary Fig S2. Abundance of differentially expressed proteins between carcinoma-associated and presumed benign groups of the training set. Protein abundance of pancreatic cyst fluid (PCyF) samples was assigned by label-free quantitative mass spectrometry (MS) analysis. **A.** Volcano plot of protein abundance, represented by Z-score values of normalized $log_2$ label-free quantitation (LFQ) intensity difference between the two groups of the training set, and plotted against the significance presented by $-log_{10}$ false discovery rate (FDR)-adjusted p-value<0.05 with S0=2.00 (two-tailed Student's t-test; Materials and methods). A generic multi-protein panel consisting of 377 differentially expressed proteins (DEPs) between the two groups (seven presumed benign and four carcinoma-associated) of the training set was identified (Fig. S2 and Table S2). Individual DEPs are indicated as colored dots (red - upregulated and blue -downregulated) in carcinoma-associated lesions. **B.** Cluster analysis of DEPs identified in PCyF samples from presumed benign and carcinoma cases of the training set. Clustered heatmap of 377 DEPs generated upon hierarchical clustering based on Z-scores of normalised $log_2$ LFQ intensity values. Values for each protein (rows) and for each sample (columns) are colored based on the protein abundance indicated in the colour scale bar shown at the bottom of the figure (high-red; low; blue) values are. The dendrogram was built using Euclidean distance in Perseus software.

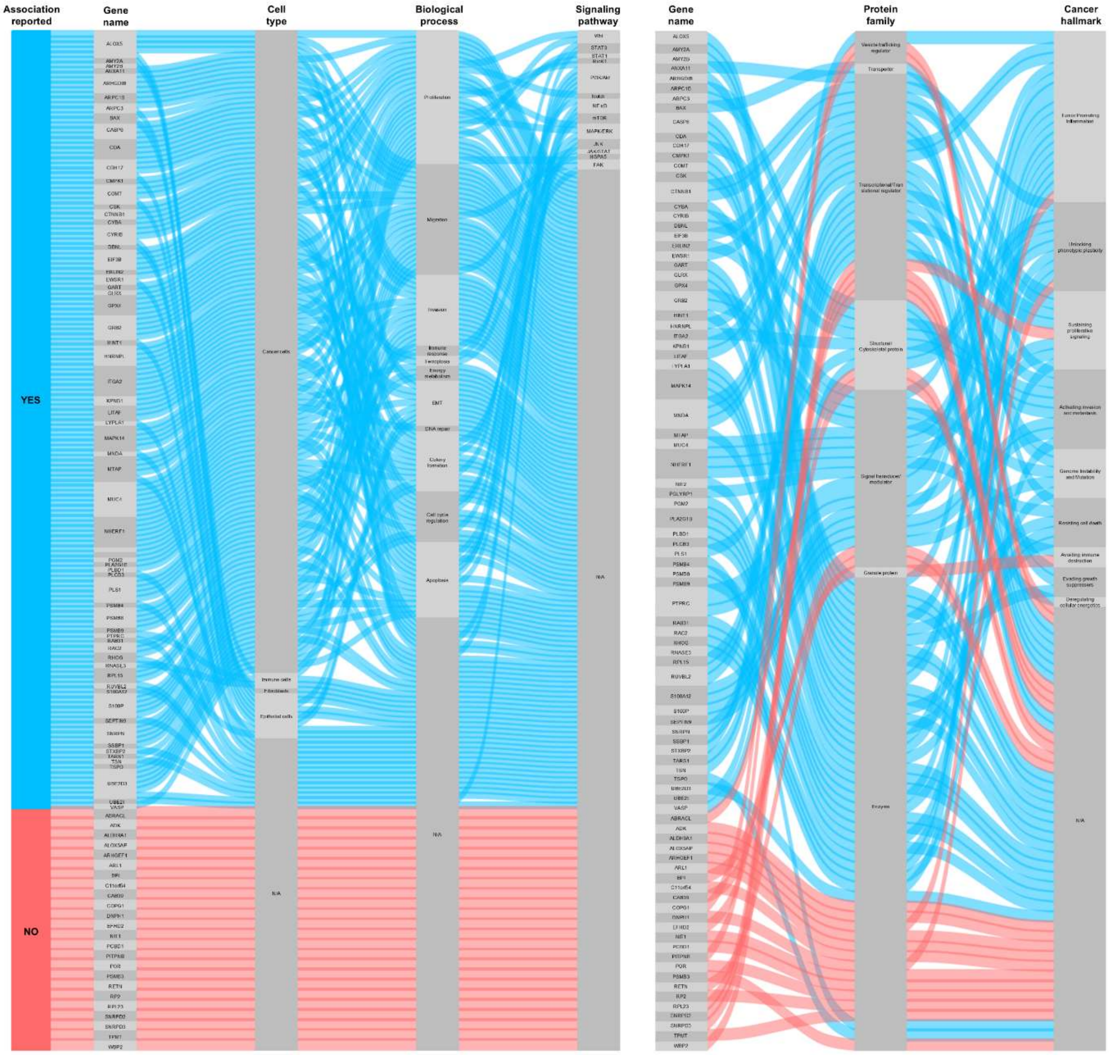

Supplementary Fig S3. Association of pancreatic cyst fluid-derived proteomic signature with pancreatic cancer based on literature and Gene Ontology database curation. ***Left,*** Alluvial plot depicting the patterns in association of the 89 differentially expressed protein (DEP; encoding genes are listed; Fig. 3A and Table S2) panel ASSIGN1, identified in pancreatic cyst fluid (PCyF), with cell type expressing these proteins, biological processes and pathways they are involved (nodes from left to right) in the context of pancreatic cancer (PC), according to previously published reports utilizing non-MS-based methods (Table S3; Materials and methods). Links between nodes are colored based on their reported association (Yes; blue) or not (No; red) with PC. ***Right,*** Alluvial plot showing the classification of ASSIGN1 members according to protein family (middle) and assigned cancer hallmark (right), both based on gene ontology (GO)[26] data curation (Table S3; Materials and methods). The majority of ASSIGN1 proteins are enzymes (48%), transcriptional/translational (20%) and signalling (15%) modulators, which are potentially involved in several hallmarks of cancer -tumour-promotingg inflammation (18%), sustained proliferativsignallingng (9%) and activation of invasion and metastasis (,9%) amongst others. Note that some proteins have been assigned to more than one cancer hallmark. Links between nodes are colored based on the reported association of ASSIGN1 members with pancreatic cancer according to non-MS-based studies (blue) or predicted from our study (red).

| Case ID | Gender | Age range at diagnosis (years) | Fukuoka high-risk | Acquisition method for samples (EUS FNA cyst fluid; EUS FNA cyst fluid and biopsy; EUS FNA cyst fluid before surgery; EUS FNA cyst fluid and biopsy before surgery; FNA cyst fluid during surgery) | Resection (immediate; within 3-6 months; upon follow up; unresectable) or No resection | Histological evaluation (dysplasia: HG - high grade; LG - low grade; CC - colloid carcinoma; BSCC - basaloid squamous cell carcinoma; PDAC - pancreatic ductal adenocarcinoma) or presumed benign | Final cyst type diagnosis |
|---|---|---|---|---|---|---|---|
| 001 | F | 61-65 | Yes | FNA cyst fluid during surgery | Immediate | Dysplasia, HG | IPMN |
| 003 | M | 76-80 | No | EUS FNA cyst fluid | No resection | Presumed benign | SCN |
| 006 | M | 71-75 | No | EUS FNA cyst fluid | No resection | Presumed benign | IPMN |
| 007 | F | 31-35 | No | EUS FNA cyst fluid | No resection | Presumed benign | SCN |
| 008 | F | 66-70 | Yes | EUS FNA cyst fluid before surgery | Within 3-6 months | Dysplasia, LG | IPMN |
| 010 | F | 71-75 | Yes | EUS FNA cyst fluid and biopsy | Non-operable (high risk for operation) | N/A | IPMN |
| 019 | M | 71-75 | Yes | EUS FNA cyst fluid before surgery | Within 3-6 months | Dysplasia, LG | IPMN |
| 021 | F | 76-80 | No | EUS FNA cyst fluid and biopsy | No resection | Presumed benign | SCN |
| 024 | M | 51-55 | No | EUS FNA cyst fluid before surgery | Upon follow-up | Presumed benign / Groove pancreatitis | Pseudocyst / Groove pancreatitis |
| 026 | M | 81-85 | No | EUS FNA cyst fluid | No resection | Presumed benign | MCN |
| 027 | F | 36-40 | Yes | EUS FNA cyst fluid before surgery | Within 3-6 months | Dysplasia, LG | MCN |
| 029 | M | 51-55 | No | EUS FNA cyst fluid and biopsy | No resection | Presumed benign | Pseudocyst |
| 030 | M | 51-55 | No | EUS FNA cyst fluid | No resection | Presumed benign | Pseudocyst |
| 034 | M | 66-70 | Yes | FNA cyst fluid during surgery | Immediate | CC | IPMN |
| 038 | F | 51-55 | Yes | FNA cyst fluid during surgery | Immediate | Dysplasia, LG | IPMN |
| 041 | F | 51-55 | Yes | EUS FNA cyst fluid and biopsy cores | Unresectable | BSCC | Malignant component |
| 042 | F | 66-70 | Yes | FNA cyst fluid during surgery | Immediate | Dysplasia, LG | IPMN |
| 045 | M | 51-55 | Yes | FNA cyst fluid during surgery | Immediate | Dysplasia, LG | IPMN |
| 047 | F | 51-55 | No | EUS FNA cyst fluid | No resection | Presumed benign | Lymphoepithelial cyst |
| 048 | M | 56-60 | Yes | EUS FNA cyst fluid and biopsy cores | Unresectable | PDAC | Malignant component |
| 052 | M | 71-75 | Yes | FNA cyst fluid during surgery | Immediate | PDAC | IPMN |
| 059 | F | 61-65 | Yes | FNA cyst fluid during surgery | Immediate | Dysplasia, LG | SCN (multicystic) |
| 073 | F | 51-55 | No | EUS FNA cyst fluid | No resection | Presumed benign | Pseudocyst |
| 075 | M | 71-75 | Yes | EUS FNA cyst fluid before surgery | Within 3-6 months | Dysplasia, LG | IPMN |
| 077 | M | 46-50 | Yes | FNA during aborted surgery | Non-operable (excessive bleeding) | PDAC | IPMN |
| 078 | F | 76-80 | Yes | EUS FNA cyst fluid | Non-operable (anaphylactic shock) | N/A | IPMN |
| 079 | F | 76-80 | Yes | FNA cyst fluid during surgery | Immediate | PDAC | IPMN |
| 095 | M | 76-80 | No | EUS FNA cyst fluid | No resection | Presumed benign | Pseudocyst |
| 096 | M | 61-65 | Yes | EUS FNA cyst fluid and biopsy cores before surgery | Within 3-6 months | PDAC | IPMN; Malignant component |
| 100 | F | 51-55 | No | EUS FNA cyst fluid | No resection | Presumed benign | MCN |
| 105 | M | 76-80 | No | EUS FNA cyst fluid | No resection | Presumed benign | SCN |

Supplementary Table S1. Patient cohort and cyst clinical information. Pancreatic cyst fluid (PCyF) specimens (n=31) obtained by endoscopic ultrasound fine-needle aspiration, along with corresponding clinical, imaging, and diagnostic surgical pathology follow-up (Materials and methods). Cyst types were diagnosed based on radiological, cytological and biochemical characteristics/features according to the 2017 International Association of Pancreatology (IAP) Fukuoka guidelines[3]. All cysts were diagnosed with either high-risk stigmata (HRS) or worrisome features (WF), according to Fukuoka criteria[3]. Pancreatic cystic lesions with HRS and/or WF with suspicious or positive for malignancy cytology and/or biochemistry were recommended for resection and subsequent histopathological assessment, whilst patients with cysts with WF and non-suspicious or negative for malignancy cytology and/or biochemistry ("presumed benign" and therefore unresected) either underwent follow-up surveillance or were discharged. Histological evaluation fora specific grade (low, intermediate or high) of dysplasia or type of malignancy determined by an expert in gastrointestinal pathology. Abbreviations: F: Female; M: Male; EUS: endoscopic ultrasound; FNA: fine-needle aspiration; IPMN: intraductal papillary mucinous neoplasm; MCN: mucinous cystic neoplasms; SCN: serous cystic neoplasm; N/A: Not available.

| Gene | Uniprot ID | Sample ID | | | | | | | | | | | | | | | | | | | | | | | | | | | | | | |
|---|---|---|---|---|---|---|---|---|---|---|---|---|---|---|---|---|---|---|---|---|---|---|---|---|---|---|---|---|---|---|---|---|
| | | 001 | 003 | 006 | 007 | 008 | 010 | 019 | 021 | 024 | 026 | 027 | 029 | 030 | 034 | 038 | 041 | 042 | 045 | 047 | 048 | 052 | 059 | 073 | 075 | 077 | 078 | 079 | 095 | 096 | 100 | 105 |
| ABRACL | Q9P1F3 | 0.52 | -1.54 | -1.34 | -1.24 | -1.36 | -1.50 | -1.16 | -1.47 | -1.81 | -1.70 | -1.82 | -1.81 | -1.99 | 0.86 | 0.06 | 0.48 | 0.58 | 0.36 | 0.00 | -0.10 | 0.09 | -0.10 | -1.68 | -1.39 | -0.21 | -0.11 | -0.56 | -2.14 | -1.27 | -1.63 | -1.41 |
| ADK | P55263 | 0.10 | -1.74 | -1.69 | -1.99 | -2.08 | -1.67 | -1.69 | -1.24 | -1.01 | -1.39 | -2.19 | -1.64 | -1.66 | 0.18 | 0.30 | -0.19 | -0.48 | -1.88 | -0.36 | 0.06 | -0.74 | 0.02 | -1.49 | -1.29 | 0.23 | 0.01 | 0.13 | -0.81 | -0.45 | -1.45 | -1.68 |
| ALDH9A1 | P49189 | 0.68 | -0.98 | -1.75 | -1.60 | -1.32 | -0.97 | -1.87 | -1.15 | -1.34 | -1.88 | -1.38 | -2.00 | -1.86 | -0.46 | 0.56 | -0.03 | 0.96 | 0.38 | -0.56 | 0.22 | -0.26 | 0.23 | -1.16 | -1.94 | 0.29 | 0.50 | 0.06 | -0.29 | -0.21 | -1.72 | -1.27 |
| ALOX5 | P09917 | -1.68 | -1.76 | -1.66 | -2.07 | -1.31 | -1.45 | -1.32 | -2.03 | -0.40 | -1.66 | -1.65 | -1.09 | -1.98 | 0.88 | -1.56 | 0.81 | -0.57 | -1.76 | -1.41 | 0.70 | -0.35 | -1.78 | -1.66 | -1.81 | 1.56 | -1.79 | 1.68 | -0.35 | -0.03 | -1.38 | -1.91 |
| ALOX5AP | P20292 | -2.07 | -1.56 | -1.54 | -1.16 | -1.77 | -1.47 | -1.40 | -1.23 | -1.11 | -1.70 | -1.40 | -1.59 | -1.54 | 0.48 | -0.70 | 0.29 | -1.91 | -1.36 | -1.82 | 1.65 | -0.18 | -1.14 | -1.73 | -1.65 | 2.50 | -0.70 | 2.50 | 0.28 | 0.63 | -1.20 | -1.60 |
| AMY2A | P04746 | 1.51 | 0.94 | 5.63 | 1.23 | 5.39 | 4.34 | 4.34 | 0.72 | 4.01 | 3.95 | 4.83 | 4.39 | 1.75 | 2.55 | 2.34 | 0.80 | 5.42 | 4.83 | 0.95 | 1.28 | 3.64 | 0.76 | 4.89 | 4.91 | 1.70 | 2.80 | 1.90 | 4.80 | 1.14 | 4.61 | 1.36 |
| AMY2B | P19961 | -0.04 | -0.50 | 3.80 | -0.32 | 3.89 | 2.10 | 2.80 | -0.18 | 2.85 | 3.06 | 3.09 | 2.74 | 1.07 | 1.29 | 0.88 | -0.10 | 3.95 | 3.40 | 0.09 | -0.50 | 2.18 | -0.48 | 3.34 | 3.35 | 0.17 | 1.52 | 0.23 | 3.44 | 0.19 | 3.45 | 0.10 |
| ANXA11 | P50995 | -0.60 | -0.54 | -1.87 | -1.35 | -2.00 | -1.89 | -1.49 | -1.49 | -0.31 | -1.48 | -1.31 | -1.34 | -0.20 | 0.30 | -0.96 | 0.21 | 1.03 | 0.19 | -1.36 | 1.65 | 1.66 | -0.94 | -1.58 | -0.24 | 2.48 | 0.30 | 2.77 | 0.72 | 0.90 | -0.99 | -1.74 |
| ARHGDIB | P52566 | 1.38 | -1.58 | -0.95 | -1.25 | -1.57 | -1.88 | -1.52 | -1.08 | 0.66 | -1.88 | -1.76 | -1.23 | -2.00 | 2.19 | 0.81 | 0.93 | 0.51 | -0.76 | 0.60 | 1.83 | 0.47 | 0.40 | -1.10 | -1.39 | 2.37 | 0.81 | 2.52 | 0.11 | 1.36 | -1.52 | -1.52 |
| ARHGEF1 | Q92888 | -0.19 | -1.73 | -0.94 | -1.63 | -1.01 | -1.74 | -1.09 | -2.38 | -2.14 | -1.66 | -1.48 | -1.13 | -1.67 | 0.28 | -0.71 | -1.07 | -0.21 | -1.65 | -0.48 | 0.06 | -0.78 | -1.72 | -1.56 | -1.36 | 0.87 | -1.22 | 0.91 | -1.34 | -0.22 | -1.50 | -1.34 |
| ARL1 | P40616 | -0.33 | -1.41 | -1.28 | -1.50 | -1.61 | -1.28 | -1.15 | -1.70 | -1.51 | -1.45 | -1.89 | -0.73 | -1.61 | -0.93 | -0.51 | -0.37 | -0.80 | -1.72 | -1.78 | 0.55 | -0.21 | -1.54 | -1.70 | -1.70 | -0.21 | -1.46 | -0.34 | -1.60 | -0.40 | -1.83 | -1.10 |
| ARPC1B | O15143 | 1.19 | -1.48 | -1.81 | -1.73 | -1.11 | -1.47 | -1.38 | -1.88 | 0.74 | -1.76 | -1.29 | -2.15 | -1.79 | 1.91 | 0.29 | 0.59 | 0.90 | 0.46 | 0.24 | 1.56 | 0.02 | -0.19 | -1.62 | -1.58 | 1.73 | 0.21 | 1.60 | 0.05 | 0.95 | -1.55 | -1.27 |
| ARPC3 | O15145 | 1.06 | -1.14 | -1.56 | -1.66 | -1.81 | -1.70 | -1.49 | -2.06 | -0.20 | -0.92 | -1.53 | -1.48 | -2.40 | 0.78 | -0.33 | 0.16 | 1.08 | 0.39 | 0.06 | 1.07 | -0.20 | -0.42 | -1.43 | -1.45 | 0.86 | -0.48 | 0.70 | -0.24 | 0.19 | -1.76 | -1.71 |
| BAX | Q07812 | -0.38 | -1.70 | -0.92 | -1.46 | -1.91 | -1.66 | -1.27 | -1.48 | -1.88 | -1.66 | -1.87 | -1.69 | -1.22 | 0.06 | -1.62 | -0.03 | 0.22 | -1.30 | -1.82 | -0.31 | -0.34 | -1.71 | -1.65 | -0.66 | -0.12 | -1.89 | -0.24 | -1.41 | 0.34 | -1.61 | -1.80 |
| BPI | P17213 | 0.49 | -2.08 | -1.49 | -1.50 | -1.19 | -1.02 | -1.27 | -1.36 | 0.45 | -1.60 | -1.24 | -1.51 | -1.58 | 2.07 | -0.12 | 0.18 | -1.69 | -1.31 | -1.84 | 1.11 | 0.18 | -0.07 | -1.12 | -1.21 | 2.69 | -0.38 | 2.93 | -0.01 | 0.70 | -1.50 | -1.29 |
| C11orf54 | Q9H0W9 | 0.61 | -1.57 | -1.64 | -1.48 | -1.63 | -1.42 | -1.72 | -1.70 | -2.03 | -1.56 | -0.95 | -1.36 | -2.09 | 0.53 | -0.08 | 0.12 | 1.25 | 0.50 | -1.43 | 0.08 | -0.54 | -0.54 | -1.44 | -1.80 | 0.34 | -0.38 | 0.33 | -1.78 | 0.54 | -1.69 | -0.94 |
| CAB39 | Q9Y376 | -0.49 | -1.45 | -1.63 | -1.73 | -1.73 | -1.13 | -1.34 | -1.66 | -0.89 | -2.29 | -1.55 | -1.56 | -0.50 | 0.12 | -1.00 | -0.65 | -0.46 | -0.59 | -1.52 | 0.11 | -0.48 | -1.25 | -0.72 | -1.34 | -0.09 | -1.10 | -0.16 | -1.56 | -0.32 | -1.62 | -1.36 |
| CASP6 | P55212 | 0.48 | -1.00 | -1.75 | -1.80 | -1.63 | -1.49 | -1.78 | -1.35 | -1.81 | -1.69 | -1.60 | -2.40 | -1.65 | 0.57 | -0.21 | -0.70 | -0.05 | -0.59 | -1.46 | 0.51 | 0.10 | -0.82 | -1.37 | -1.62 | 0.65 | -0.22 | 0.72 | -0.02 | -0.15 | -1.80 | -1.73 |
| CDA | P32320 | 0.37 | -1.15 | -2.15 | -1.83 | -1.98 | -1.07 | -1.48 | -1.77 | -0.15 | -1.62 | -1.43 | -1.75 | -1.98 | 1.34 | -0.65 | 0.38 | -1.57 | -1.44 | -1.50 | 0.99 | -0.27 | -1.59 | -2.08 | -2.08 | 1.47 | -0.51 | 1.78 | -2.00 | 0.75 | -1.49 | -1.90 |
| CDH17 | Q12864 | 1.23 | -2.20 | -1.82 | -1.83 | -1.09 | -1.14 | -1.45 | -2.03 | -1.44 | -1.61 | -1.90 | -1.95 | -0.92 | 0.88 | -1.50 | -0.88 | -0.94 | -1.81 | -1.89 | -0.43 | -0.82 | -1.60 | -1.75 | -1.29 | 0.50 | -0.20 | -1.72 | -1.72 | 0.01 | -1.47 | -1.41 |
| CMPK1 | P30085 | 1.69 | -0.34 | -0.99 | -1.69 | -1.57 | -1.29 | -1.75 | -1.93 | 1.65 | -1.50 | -1.72 | -1.32 | -1.42 | -0.77 | 1.10 | 0.13 | 2.09 | 1.30 | -1.06 | 1.66 | 0.35 | 0.55 | -1.95 | -0.12 | 0.31 | 0.59 | 0.73 | 0.32 | 0.87 | -1.66 | -1.47 |
| COMT | P21964 | 0.51 | -1.73 | -1.76 | -1.61 | -1.57 | -1.01 | -0.74 | -1.57 | -1.28 | -1.31 | -1.99 | -1.75 | -1.19 | -0.49 | -1.58 | 0.14 | 0.83 | 0.15 | -1.51 | 1.49 | -0.25 | -0.30 | -1.40 | -1.58 | 0.96 | -0.83 | 0.77 | -0.32 | 0.54 | -1.21 | -1.48 |
| COPG1 | Q9Y678 | 0.88 | -1.38 | -1.65 | -1.59 | -1.63 | -1.18 | -1.28 | -1.45 | -1.97 | -1.27 | -1.91 | -1.73 | -1.56 | -0.32 | -0.19 | 0.23 | 0.15 | 0.08 | -1.50 | 0.49 | -0.71 | -1.50 | -1.61 | -2.09 | 0.09 | -0.25 | 0.08 | -0.58 | -0.34 | -1.69 | -0.28 |
| CSK | P41240 | -0.12 | -1.64 | -1.63 | -1.47 | -1.36 | -1.32 | -1.43 | -1.66 | -1.62 | -1.40 | -1.89 | -1.90 | -1.58 | -0.34 | -0.24 | -0.44 | -0.18 | -0.52 | -0.46 | 0.05 | -1.34 | -1.67 | -2.02 | -1.72 | 0.38 | -1.33 | 0.12 | -0.62 | 0.25 | -1.45 | -1.46 |
| CTNNB1 | P35222 | 0.55 | -2.19 | -1.90 | -1.89 | 0.37 | -1.23 | -1.53 | -1.65 | -1.55 | -1.51 | -1.47 | -1.88 | -2.13 | -0.30 | -0.97 | -0.49 | 0.58 | -0.28 | -1.78 | 0.12 | 0.22 | -0.88 | -1.86 | -1.26 | -1.22 | -1.80 | -0.97 | -0.71 | -0.35 | -1.34 | -1.42 |
| CYBA | P13498 | -1.63 | -1.64 | -1.24 | -1.37 | -1.05 | -1.46 | -1.42 | -1.26 | -0.57 | -1.41 | -1.38 | -1.54 | -1.87 | -0.28 | -1.94 | 0.29 | -2.12 | -1.23 | -1.76 | 1.29 | -0.37 | -1.68 | -1.50 | -1.23 | 1.75 | -1.56 | 1.71 | -1.28 | 0.72 | -1.84 | -1.41 |
| CYRIB | Q9NUQ9 | 0.35 | -1.35 | -0.89 | -1.67 | -0.97 | -1.37 | -1.65 | -1.22 | -1.68 | -2.25 | -1.17 | -2.16 | -1.49 | 0.83 | 0.09 | 0.39 | 0.31 | 0.43 | -0.04 | 0.60 | -0.21 | -0.20 | -1.27 | -1.30 | 0.61 | -0.06 | 0.41 | -1.37 | 0.39 | -1.75 | -1.89 |
| DBNL | Q9UJU6 | 0.39 | -1.62 | -1.01 | -1.59 | -1.70 | -1.43 | -1.36 | -1.80 | -0.29 | -0.87 | -1.79 | -1.51 | -1.49 | 0.95 | 0.00 | -0.13 | 0.51 | 0.32 | -0.21 | 0.31 | 0.05 | -0.21 | -1.87 | 1.04 | 0.43 | -0.08 | 0.69 | -0.92 | -0.12 | -1.36 | -1.43 |
| DNPH1 | O43598 | 0.58 | -1.87 | -1.43 | -1.33 | -1.50 | -1.83 | -1.17 | -1.58 | -0.94 | -1.96 | -1.02 | -2.26 | -1.88 | 0.48 | 1.05 | 0.35 | 1.11 | 0.63 | -1.56 | 0.27 | 0.03 | 0.82 | -1.52 | -1.71 | -0.30 | 0.86 | 0.25 | -0.83 | 0.60 | -1.41 | -1.78 |
| EFHD2 | Q96C19 | 0.75 | -2.02 | -1.89 | -2.14 | -1.37 | -1.83 | -1.58 | -1.96 | -1.48 | -1.21 | -1.37 | -1.70 | -1.17 | 0.50 | -0.59 | 0.09 | 0.60 | 0.41 | -1.08 | 0.56 | -1.02 | -1.60 | -2.07 | -1.37 | 0.69 | -1.01 | 0.64 | 0.17 | 0.15 | -1.37 | -1.34 |
| EIF3B | P55884 | 0.23 | -2.00 | -1.94 | -1.84 | 0.47 | -1.31 | -0.80 | -1.42 | -1.24 | -1.29 | -1.29 | -1.07 | -1.07 | 0.27 | -1.51 | 0.05 | -0.22 | -0.07 | -0.25 | 0.35 | -0.27 | -1.62 | -0.73 | -1.68 | -0.01 | -0.63 | 0.14 | -0.22 | -0.22 | -1.34 | -1.72 |
| ERLIN2 | O94905 | -0.83 | -1.88 | -2.06 | -1.37 | -1.78 | -1.26 | -1.41 | -1.69 | -1.27 | -1.67 | -1.05 | -1.43 | -2.26 | -1.28 | -0.88 | 0.04 | -0.05 | -1.56 | -1.22 | 0.71 | -0.19 | -1.81 | -1.93 | -2.15 | 0.56 | -1.58 | 0.37 | -1.71 | -0.23 | -1.51 | -1.84 |
| EWSR1 | Q01844 | 0.78 | -1.14 | -1.09 | -1.41 | -0.95 | -1.32 | -1.43 | -1.02 | -1.74 | -1.38 | -1.37 | -1.33 | -1.73 | 0.81 | -1.59 | 0.41 | 0.75 | -0.15 | 0.23 | 0.14 | -1.28 | -1.49 | -1.16 | -2.03 | 0.52 | -1.44 | 0.52 | -1.99 | 0.43 | -1.86 | -1.30 |
| GART | P22102 | -0.04 | -2.04 | -1.33 | -1.79 | -1.22 | -1.91 | -0.90 | -1.36 | -1.82 | -1.93 | -1.51 | -1.93 | -1.18 | -0.60 | -0.25 | -0.28 | 0.11 | -0.50 | -1.62 | 0.24 | -0.86 | -0.61 | -1.47 | -1.66 | 0.03 | -0.79 | 0.21 | 0.28 | -0.26 | -1.79 | -1.13 |
| GLRX | P35754 | 0.89 | -0.76 | -1.60 | -2.22 | -1.53 | -1.73 | -1.45 | -1.33 | -1.54 | -2.19 | -1.63 | -1.58 | -1.13 | -0.28 | 0.76 | 0.29 | 0.07 | 0.09 | -1.36 | 0.15 | -0.02 | 0.26 | -1.75 | -1.60 | 0.88 | 0.01 | 1.02 | -1.49 | 0.22 | -1.64 | -1.18 |
| GPX4 | P36969 | -0.53 | -1.33 | -1.88 | -1.26 | -1.55 | -1.60 | -1.66 | -1.61 | -0.79 | -1.56 | -0.50 | -2.38 | -1.75 | 0.43 | 0.11 | -0.85 | 0.41 | -0.49 | -1.41 | 0.13 | -0.47 | -0.60 | -1.65 | -1.36 | 0.30 | -0.22 | 0.51 | -0.55 | -0.48 | -1.30 | -1.90 |
| GRB2 | P62993 | 0.11 | -1.49 | -1.08 | -1.45 | -1.53 | -1.34 | -1.81 | -1.43 | -1.66 | -1.65 | -1.64 | -0.88 | -1.37 | -0.36 | -0.73 | -0.03 | -0.27 | -0.67 | -0.65 | 0.23 | -0.84 | -1.88 | -1.76 | -0.91 | 0.31 | -0.89 | 0.12 | -1.64 | -0.08 | -1.68 | -1.28 |
| HINT1 | P49773 | 1.18 | -0.84 | -1.56 | -1.63 | -1.38 | -1.61 | -1.21 | -1.71 | -0.49 | -1.10 | -1.67 | -1.88 | -1.66 | -0.12 | 1.17 | 0.35 | 1.14 | 0.74 | -0.13 | 0.50 | -0.68 | 0.77 | -1.35 | -2.04 | 0.37 | 1.05 | 0.29 | -0.03 | 0.38 | -1.64 | -1.67 |
| HNRNPL | P14866 | 0.65 | -1.36 | -1.60 | -1.48 | -1.63 | -1.40 | -1.42 | -1.60 | -1.15 | -1.29 | -1.59 | -1.05 | -1.06 | 0.17 | -0.60 | -0.04 | 0.66 | -0.55 | -0.05 | 0.50 | -0.77 | -0.72 | -1.19 | -1.21 | 0.48 | -1.15 | 0.64 | -1.15 | -0.18 | -1.53 | -2.19 |
| ITGA2 | P17301 | -0.54 | -0.32 | -1.21 | -1.69 | -1.62 | -1.87 | -0.89 | -1.86 | -2.01 | -1.68 | -1.34 | -1.31 | -1.18 | -0.13 | -1.32 | -0.38 | -0.51 | -1.50 | 0.01 | 1.03 | 0.26 | -0.55 | -1.62 | -1.45 | -0.25 | -2.11 | 0.06 | -0.99 | -0.10 | -1.28 | -1.80 |
| KPNB1 | Q14974 | 0.54 | -1.50 | -1.50 | -1.57 | -1.73 | -1.20 | -1.73 | -1.82 | 0.08 | -1.40 | -1.25 | -1.49 | -2.38 | -0.24 | 0.74 | 0.20 | 0.52 | 0.03 | -0.34 | 0.29 | -0.62 | 0.42 | -1.88 | -1.20 | 0.01 | 0.50 | -0.05 | -1.73 | 0.38 | -1.65 | -1.56 |
| LITAF | Q99732 | -0.89 | -0.12 | -0.85 | -0.98 | -1.82 | -1.52 | -1.67 | -1.72 | -1.19 | -1.44 | -1.14 | -1.49 | -0.73 | 0.18 | -1.23 | -0.14 | -0.44 | -1.27 | -1.43 | 1.08 | 0.32 | -0.81 | -1.70 | -0.73 | 1.85 | -1.35 | 2.03 | -0.56 | 0.31 | -1.30 | -1.44 |
| LYPLA1 | O75608 | 0.35 | -1.87 | -1.92 | -2.04 | -1.52 | -1.31 | -1.52 | -1.59 | -1.32 | -1.55 | -1.57 | -1.95 | -1.82 | 0.37 | -0.01 | -1.08 | 0.61 | -0.07 | -1.13 | 0.19 | -0.37 | -0.58 | -1.36 | -1.81 | 0.13 | -0.36 | -0.33 | -1.65 | -0.52 | -1.67 | -1.19 |
| MAPK14 | Q16539 | 0.19 | -1.09 | -1.67 | -1.48 | -1.08 | -1.77 | -1.46 | -1.61 | -1.35 | -1.76 | -2.24 | -1.40 | -1.60 | 0.14 | -0.73 | -0.34 | 0.00 | -0.53 | -0.87 | 0.57 | -0.82 | -1.63 | -1.87 | -1.98 | 0.99 | -0.70 | 0.72 | -0.44 | -1.35 | -2.15 | -1.55 |
| MNDA | P41218 | 1.34 | -1.41 | -2.15 | -1.75 | -1.54 | -1.60 | -1.12 | -1.96 | 1.15 | -1.65 | -1.37 | -1.12 | -1.49 | 1.86 | 0.42 | 1.06 | -1.80 | -2.03 | -1.37 | 2.28 | 0.16 | 1.95 | -1.82 | -1.76 | 2.56 | 0.04 | 2.72 | 0.33 | 1.32 | -1.91 | -1.78 |
| MTAP | Q13126 | -0.03 | -1.62 | -1.66 | -1.38 | -1.99 | -1.29 | -2.22 | -1.87 | -1.36 | -1.84 | -1.46 | -1.45 | -1.69 | -0.23 | 0.25 | -0.49 | 0.48 | -0.22 | -0.52 | 0.12 | -0.84 | 0.02 | -1.29 | -1.50 | -0.07 | 0.13 | -0.09 | -1.47 | -0.36 | -1.12 | -1.90 |
| MUC4 | Q99102 | 0.18 | 0.28 | -1.20 | -1.33 | -1.69 | -1.18 | -1.03 | -1.56 | -2.09 | -1.94 | -1.55 | -1.55 | -1.12 | 0.00 | -1.48 | -0.52 | -1.28 | -1.48 | -1.95 | 0.89 | -1.13 | -2.04 | -1.66 | -1.50 | -0.63 | 0.10 | 0.84 | -1.25 | -0.76 | -1.29 | -1.80 |
| NIT1 | Q86X76 | 0.20 | -1.67 | -1.30 | -1.74 | -1.86 | -1.30 | -2.34 | -1.55 | -1.31 | -1.32 | -1.50 | -1.68 | -1.65 | -0.01 | 0.12 | -0.30 | 0.42 | 0.20 | -1.29 | 0.02 | 0.34 | 0.44 | -1.70 | -2.27 | 0.44 | 0.27 | 0.51 | -1.36 | -0.08 | -1.41 | -1.20 |
| NIT2 | Q9NQR4 | 0.34 | -2.22 | -1.27 | -1.69 | -1.82 | -1.81 | -1.53 | -1.49 | -1.68 | -1.40 | -1.72 | -1.46 | -1.35 | -0.31 | -0.25 | -0.08 | 0.53 | -0.22 | -1.25 | 0.01 | -0.68 | 0.03 | -1.46 | -1.62 | 0.10 | -0.38 | -0.23 | -1.04 | -0.87 | -1.44 | -1.26 |
| PCBD1 | P61457 | 1.05 | -0.02 | -0.89 | -1.61 | -1.19 | -1.68 | -1.30 | -1.54 | -1.39 | -1.65 | -1.36 | -1.08 | -1.32 | -1.04 | -0.13 | 0.04 | 0.72 | 0.33 | -0.97 | 0.19 | -0.20 | -0.26 | -1.74 | -1.42 | -0.47 | -0.77 | -0.12 | -0.82 | 0.15 | -1.24 | -1.82 |
| PGLYRP1 | O75594 | 0.96 | -1.18 | -1.76 | -1.87 | -1.35 | -1.62 | -1.37 | -1.61 | 0.16 | -1.64 | -1.74 | -1.08 | -1.56 | 2.06 | -0.40 | 0.65 | -1.77 | -1.70 | -1.78 | 1.02 | -0.40 | -0.51 | -1.80 | -1.41 | 2.32 | -0.23 | 2.75 | 0.07 | 1.01 | -1.22 | -1.35 |
| PGM2 | Q96G03 | 0.52 | 0.25 | -1.07 | -1.49 | -1.27 | -1.48 | -1.19 | -1.34 | -1.53 | -1.46 | -1.55 | -1.33 | -1.32 | 0.72 | 0.67 | 0.16 | 0.58 | -0.24 | -0.10 | 0.28 | -0.81 | 0.03 | -1.50 | -1.87 | 0.67 | 0.49 | 0.57 | -1.57 | 0.18 | -1.49 | -1.52 |
| PITPNB | P48739 | 0.30 | -2.26 | -2.51 | -1.32 | -1.66 | -1.51 | -1.53 | -1.43 | -1.47 | -1.86 | -1.45 | -2.12 | -1.37 | 0.02 | 0.15 | 0.02 | -0.14 | -0.62 | -1.59 | -0.09 | -0.38 | -0.37 | -1.87 | 0.52 | -0.40 | -0.02 | -0.21 | -0.44 | -0.10 | -1.70 | -1.64 |
| PLA2G1B | P04054 | 0.72 | -0.24 | 4.12 | 0.42 | 4.56 | 2.15 | 3.72 | -0.44 | 2.59 | 2.68 | 3.63 | 2.21 | 0.49 | 1.16 | 1.53 | -0.34 | 4.66 | 4.50 | 0.13 | -0.23 | 2.15 | -0.07 | 3.14 | 4.36 | 1.07 | 1.84 | 0.85 | 4.70 | 0.34 | 3.48 | 0.21 |
| PLBD1 | Q6P4A8 | 0.30 | -1.43 | -2.18 | -0.96 | -1.71 | -2.06 | -1.09 | -1.52 | -0.08 | -1.22 | -1.85 | -2.00 | -1.58 | 1.64 | 0.00 | 0.02 | 0.30 | 0.06 | -1.69 | 0.75 | 0.42 | -1.20 | -1.59 | 1.15 | 2.22 | -1.42 | 3.06 | -0.09 | 0.55 | -1.72 | -1.84 |
| PLCB3 | Q01970 | -0.63 | -1.66 | -1.53 | -1.51 | -1.35 | -1.27 | -1.69 | -1.18 | -1.52 | -1.22 | -1.44 | -1.40 | -1.22 | 0.31 | -1.49 | -0.74 | -0.65 | -1.98 | -1.41 | 0.29 | -0.32 | -1.51 | -1.72 | -1.82 | -1.03 | -1.36 | -0.22 | -2.00 | 0.09 | -1.57 | -1.54 |

| | | | | | | | | | | | | | | | | | | | | | | | | | | | | | | | | |
|---|---|---|---|---|---|---|---|---|---|---|---|---|---|---|---|---|---|---|---|---|---|---|---|---|---|---|---|---|---|---|---|---|
| PLS1 | Q14651 | 1.09 | -1.05 | -1.63 | -1.85 | -1.16 | -1.21 | -1.63 | -1.34 | -1.62 | -1.42 | -1.81 | -1.33 | -1.62 | -0.14 | -0.34 | 0.68 | 0.97 | 0.52 | -1.42 | 0.64 | 0.56 | -1.21 | -1.25 | -1.87 | 0.62 | -0.45 | 0.25 | 0.06 | 0.29 | -1.98 | -1.72 |
| POR | P16435 | -0.11 | -1.55 | -0.99 | -1.38 | -1.74 | -1.27 | -1.80 | -1.51 | -1.39 | -1.76 | -1.62 | -1.67 | -1.41 | -0.38 | -1.67 | -0.30 | 0.00 | -1.53 | -1.43 | 0.70 | -1.01 | -1.12 | -1.46 | -1.74 | 0.07 | -1.86 | 0.14 | -1.23 | -0.37 | -1.71 | -2.29 |
| PSMB3 | P49720 | -0.01 | -1.40 | -1.86 | -1.56 | -1.67 | -1.30 | -0.04 | -1.81 | -2.09 | -1.51 | -1.66 | -1.99 | -1.58 | -0.45 | 0.28 | -0.06 | -0.31 | -1.10 | -1.81 | 0.16 | -0.73 | -0.75 | -1.16 | 0.02 | -0.46 | 0.08 | -0.26 | -0.24 | -0.46 | -1.63 | -1.39 |
| PSMB4 | P28070 | 0.47 | -1.13 | -1.52 | -1.68 | -2.14 | -1.40 | -1.56 | -1.38 | -1.47 | -1.53 | -1.86 | -1.72 | -1.46 | -0.41 | 0.64 | 0.35 | 0.34 | -0.30 | -1.69 | 0.38 | -0.58 | 0.13 | -1.28 | -1.04 | -0.04 | 0.54 | 0.02 | -1.03 | 0.27 | -1.37 | -1.31 |
| PSMB8 | P28062 | 0.49 | -1.93 | -1.77 | -1.04 | -0.76 | -1.79 | -1.11 | -1.29 | -0.43 | -1.31 | -1.80 | -1.70 | -1.92 | 0.40 | -0.92 | -0.34 | -0.20 | -0.41 | -1.20 | 0.40 | -0.64 | -1.19 | -1.73 | -1.99 | 0.31 | -1.98 | 0.62 | -0.94 | -0.39 | -0.49 | -2.11 |
| PSMB9 | P28065 | 0.48 | -1.48 | -1.65 | -1.64 | -1.42 | -1.52 | -1.42 | -1.53 | -1.16 | -1.70 | -1.75 | -1.79 | -1.96 | 0.00 | -0.99 | -0.06 | -0.20 | -0.85 | -0.23 | 0.33 | -0.97 | -1.19 | -1.47 | -1.68 | 0.25 | -0.97 | 0.59 | -1.11 | -0.17 | -0.93 | -1.90 |
| PTPRC | P08575 | -1.94 | -0.41 | -1.45 | -2.02 | -1.59 | -1.64 | -1.80 | -1.57 | 0.08 | -1.13 | -1.70 | -1.46 | -1.26 | 0.65 | -1.60 | -0.02 | -0.97 | -1.09 | -0.11 | 0.68 | -0.25 | -1.37 | -1.10 | -2.33 | 1.59 | -1.75 | 1.81 | -0.46 | 0.08 | -1.45 | -1.60 |
| RAB31 | Q13636 | -0.25 | -2.06 | -1.70 | -1.48 | -1.45 | -1.43 | -1.65 | -1.62 | -1.53 | -0.76 | -1.47 | -2.26 | -1.75 | 0.31 | -1.26 | -0.56 | -1.31 | -1.94 | -1.98 | 0.58 | -0.88 | -2.03 | -2.11 | -1.90 | 1.28 | -1.42 | 1.44 | -0.46 | -1.34 | -1.93 | -1.49 |
| RAC2 | P15153 | 0.91 | -1.50 | -1.14 | -1.12 | -1.57 | -0.96 | -1.44 | -1.93 | -0.74 | -1.39 | -1.65 | -2.11 | -1.79 | 0.01 | -0.02 | 0.53 | -0.90 | -1.13 | 0.70 | 1.72 | 0.04 | -0.07 | -1.24 | -1.32 | 2.36 | 0.01 | 2.35 | -0.61 | 1.19 | -1.90 | -1.69 |
| RETN | Q9HD89 | 1.06 | -0.67 | -1.86 | -1.77 | -1.70 | -1.41 | -1.67 | -1.40 | 0.69 | -1.04 | -1.51 | -1.40 | -1.33 | 1.58 | -0.44 | 0.82 | -1.82 | 0.79 | -1.17 | 1.11 | 0.98 | -0.67 | -0.83 | -1.28 | 2.57 | -0.22 | 2.58 | -0.04 | 0.90 | -1.01 | -1.74 |
| RHOG | P84095 | -0.34 | -0.76 | -1.92 | -1.69 | -0.96 | -1.78 | -1.28 | -1.43 | 0.27 | -1.11 | -1.61 | -1.88 | -1.69 | 0.53 | -1.59 | -0.58 | -0.46 | -0.77 | -0.49 | 0.56 | -0.16 | -1.25 | -1.22 | -1.66 | 1.19 | -1.00 | 1.01 | -1.07 | 0.18 | -1.33 | -1.86 |
| RNASE3 | P12724 | 1.18 | -1.13 | -1.66 | -1.45 | -0.94 | -1.73 | -1.04 | -1.44 | 1.15 | -1.43 | -1.93 | -1.09 | -1.46 | 2.23 | -0.21 | 0.57 | -1.97 | -1.35 | -1.88 | 1.90 | 1.78 | -1.37 | -1.44 | 1.40 | 3.57 | 0.33 | 3.58 | 1.09 | 1.41 | -0.88 | -2.26 |
| RP2 | O75695 | -0.90 | -2.03 | -1.75 | -1.72 | -1.53 | -1.85 | -1.52 | -1.72 | 0.21 | -1.60 | -1.30 | -1.77 | -1.84 | 0.34 | -0.32 | -0.58 | -0.73 | -1.74 | -1.99 | 0.36 | -0.88 | -1.50 | -1.62 | -1.52 | 0.70 | -0.51 | 0.65 | -1.39 | -0.54 | -1.81 | -1.08 |
| RPL15 | P61313 | 0.37 | -0.82 | -0.98 | -1.90 | -1.51 | -1.69 | -1.63 | -1.64 | -1.77 | -2.40 | -1.90 | -1.97 | -1.73 | -0.27 | -0.56 | -0.06 | 0.14 | -0.46 | -0.48 | 0.45 | -0.46 | -0.97 | -1.73 | -1.36 | -0.04 | -1.95 | 0.35 | 0.26 | -0.40 | -1.72 | -1.28 |
| RPL23 | P62829 | 0.07 | -0.70 | -1.78 | -1.56 | -1.32 | -0.92 | -1.74 | -1.50 | -1.58 | -1.82 | -1.17 | -1.48 | -1.24 | -1.27 | -0.54 | -0.06 | 0.56 | -0.22 | -0.32 | 0.73 | -0.11 | -1.04 | -1.47 | -2.32 | 0.13 | -1.75 | 0.65 | 0.08 | -0.22 | -1.36 | -2.04 |
| RUVBL2 | Q9Y230 | 0.32 | -1.42 | -2.10 | -1.66 | -0.17 | -1.51 | -0.95 | -1.89 | -1.74 | -1.88 | -1.57 | -1.40 | -0.90 | -0.63 | 0.32 | 0.19 | 0.22 | -0.19 | -0.81 | 0.27 | -0.84 | -0.27 | -1.59 | -1.92 | -0.20 | 0.44 | -0.05 | -1.39 | 0.06 | -1.51 | -1.43 |
| S100A12 | P80511 | 0.69 | -1.93 | -0.85 | -1.09 | -1.50 | -1.13 | -1.36 | -1.79 | 0.47 | -1.65 | -1.14 | -0.77 | -0.79 | 1.33 | -1.35 | 0.55 | -1.79 | -1.61 | -1.47 | 0.79 | 0.52 | -1.35 | -1.69 | -2.02 | 2.79 | -1.83 | 2.54 | 0.24 | 0.27 | -1.58 | -1.22 |
| S100P | P25815 | 1.64 | -1.47 | -1.55 | -1.59 | -1.11 | -1.48 | -1.18 | -1.68 | -0.09 | -1.89 | -0.63 | -1.50 | -2.04 | 0.93 | 0.49 | 0.02 | 1.32 | 1.57 | -1.84 | 1.42 | 1.06 | -1.79 | -1.74 | 0.13 | 1.38 | -0.56 | 1.10 | 0.79 | 0.37 | -1.64 | -1.59 |
| SEPTIN9 | Q9UHD8 | 0.51 | -1.23 | -1.41 | -1.66 | -1.40 | -1.10 | -1.05 | -1.48 | -1.20 | -1.94 | -1.84 | -1.58 | -1.86 | -0.31 | -1.20 | -0.61 | 0.38 | -0.52 | -0.52 | 0.32 | -0.85 | -1.11 | -1.32 | -1.58 | 0.10 | -1.74 | -0.17 | -2.14 | -0.54 | -1.73 | -1.37 |
| SLC9A3R1 | O14745 | 0.47 | 0.21 | -1.19 | -1.52 | -1.24 | -1.79 | -1.67 | -1.34 | -0.99 | -1.26 | -1.42 | -1.19 | 0.79 | -0.03 | -0.75 | -0.54 | 1.17 | 0.88 | 0.24 | 1.09 | 0.64 | -0.47 | -1.21 | 0.13 | -0.28 | -0.45 | -0.15 | 0.21 | 0.45 | -1.49 | -1.80 |
| SNRPD2 | P62316 | 0.43 | -1.14 | -0.76 | -1.21 | -0.87 | -1.92 | -0.94 | -1.48 | -1.02 | -1.77 | -1.53 | -1.67 | -0.84 | 0.55 | -1.15 | 0.05 | 0.32 | -0.72 | -0.31 | 0.17 | -0.47 | -0.35 | -1.51 | -1.40 | 0.52 | -1.90 | 0.74 | -1.61 | -0.29 | -1.75 | -1.44 |
| SNRPD3 | P62318 | 0.59 | -1.11 | -1.96 | -1.20 | -1.21 | -0.05 | -1.66 | -1.01 | -0.64 | -1.80 | -1.57 | -1.62 | -1.30 | 1.19 | -1.18 | 0.31 | 0.39 | -0.74 | -0.49 | 0.74 | -0.52 | -1.73 | -1.72 | -1.50 | 0.81 | -1.66 | 1.10 | -1.47 | -0.45 | -1.74 | -1.11 |
| SNRPN | P63162 | 0.45 | -1.82 | -1.66 | -1.93 | -1.65 | -1.60 | -1.72 | -1.67 | -0.77 | -1.09 | -1.88 | -1.84 | -1.88 | 0.82 | -0.93 | 0.18 | 0.29 | -0.76 | -0.87 | 0.59 | -0.17 | -1.30 | -1.36 | -1.45 | 1.02 | -1.44 | 1.21 | -1.62 | -0.50 | -1.39 | -1.32 |
| SSBP1 | Q04837 | 0.64 | -1.16 | -1.37 | -1.20 | -1.26 | -1.30 | -1.54 | -1.89 | -0.33 | -1.64 | -1.74 | -1.45 | -1.37 | -0.13 | -1.61 | 0.49 | 0.57 | -0.06 | -1.33 | 0.81 | 0.35 | -1.75 | -1.78 | -1.43 | 1.04 | -1.22 | 1.29 | -0.47 | -0.19 | -1.87 | -1.80 |
| STXBP2 | Q15833 | 0.04 | -0.51 | -1.15 | -1.64 | -1.00 | -1.58 | -1.60 | -1.63 | 0.26 | -1.63 | -1.34 | -1.74 | -0.38 | 0.30 | -0.82 | -0.89 | 0.20 | 0.04 | -1.41 | 0.67 | -0.01 | -1.62 | -0.74 | -0.49 | 0.24 | -0.31 | 0.29 | -0.76 | 0.17 | -1.47 | -2.00 |
| TARS1 | P26639 | 0.74 | -1.89 | -1.20 | -1.72 | -1.48 | -1.42 | -2.13 | -1.66 | -0.03 | -1.36 | -1.36 | -1.74 | -1.47 | 0.26 | -0.46 | -0.34 | -0.13 | 0.22 | -1.71 | 0.01 | -0.54 | -0.97 | -1.50 | -1.01 | 0.00 | -0.48 | 0.21 | -0.40 | -1.48 | -1.61 | -0.90 |
| TPMT | P51580 | 0.44 | -1.40 | -1.74 | -1.44 | -1.66 | -1.65 | -1.44 | -1.52 | -1.72 | -1.18 | -2.09 | -1.40 | -2.20 | -0.19 | -0.18 | -0.57 | 0.04 | -1.28 | -1.70 | 0.85 | -0.28 | -1.67 | -1.85 | -1.39 | 0.40 | -0.69 | 0.26 | -0.48 | -0.37 | -1.85 | -1.89 |
| TSN | Q15631 | -0.16 | -0.58 | -1.69 | -1.44 | -1.53 | -1.29 | -1.81 | -1.64 | -2.24 | -1.08 | -1.53 | -1.65 | -1.59 | 0.22 | 0.05 | 0.14 | -0.11 | -0.76 | -1.65 | 0.05 | -0.82 | -0.58 | -1.35 | -1.59 | -0.07 | -0.15 | 0.27 | -1.40 | -0.48 | -1.68 | -1.45 |
| TSPO | P30536 | -1.52 | -1.88 | -1.96 | -1.40 | -1.82 | -1.59 | -1.52 | -1.34 | -1.57 | -1.46 | -1.68 | -1.85 | -1.72 | -0.86 | -1.84 | -0.21 | 0.55 | -1.36 | -1.49 | 1.29 | 0.14 | -1.58 | -1.91 | -1.42 | 0.74 | -2.00 | 1.20 | 0.34 | 0.64 | -1.67 | -1.69 |
| UBE2D3 | P61077 | 0.55 | -1.82 | -1.73 | -1.70 | -1.22 | -1.56 | -1.29 | -1.03 | -1.84 | -1.57 | -1.80 | -1.64 | -1.74 | 0.51 | 0.54 | -0.28 | 0.44 | -0.06 | -1.03 | 0.49 | -1.14 | -0.18 | -1.71 | -1.41 | 0.59 | 0.31 | 0.47 | -1.55 | -0.11 | -0.77 | -1.46 |
| UBE2I | P63279 | 0.83 | -1.63 | -1.58 | -1.73 | -1.66 | -1.74 | -1.62 | -1.69 | -0.95 | -1.57 | -1.37 | -1.85 | -1.64 | 0.58 | 0.37 | 0.48 | 0.81 | -0.07 | 0.26 | 0.32 | -0.39 | 0.19 | -1.61 | -1.51 | 0.45 | 0.30 | 0.58 | -2.10 | 0.29 | -1.87 | -1.39 |
| VASP | P50552 | 1.12 | -1.81 | -1.84 | -1.50 | -2.01 | -1.45 | -0.26 | -1.88 | -0.89 | -2.00 | -1.16 | -2.03 | -1.63 | 0.76 | 0.15 | 0.21 | 1.02 | 0.43 | -0.15 | 1.01 | -0.04 | -0.24 | -2.19 | 1.25 | 1.27 | 0.19 | 1.08 | -0.39 | 0.78 | -1.65 | -1.71 |
| WBP2 | Q969T9 | -0.41 | -1.23 | -1.94 | -1.81 | -2.14 | -1.46 | -1.46 | -2.35 | -0.59 | -1.50 | -1.83 | -1.57 | -1.50 | -0.52 | 0.45 | -0.42 | -0.73 | -0.73 | -1.51 | -0.31 | -0.38 | -0.14 | -1.70 | -1.97 | 0.03 | 0.25 | -0.13 | 0.54 | -0.42 | -1.67 | -1.27 |
| ABCB6 | Q9NP58 | 1.82 | -0.22 | -1.25 | -1.08 | -1.26 | -1.58 | -0.70 | -1.49 | -0.67 | -0.70 | -1.79 | -1.21 | -1.53 | 0.85 | 1.46 | 1.31 | 1.73 | 0.94 | 0.05 | 2.18 | 0.86 | 0.96 | -0.80 | -0.67 | 1.36 | 1.36 | 1.21 | 0.30 | 1.46 | 0.07 | -0.94 |
| ABHD14B | Q96IU4 | 0.74 | -0.72 | -1.50 | -1.44 | -1.13 | -1.51 | -1.48 | -1.30 | -1.68 | -1.03 | -1.07 | -1.27 | -1.79 | 0.50 | 0.74 | -0.02 | 1.07 | 0.37 | -0.08 | 0.86 | -0.54 | 0.19 | -1.08 | -0.80 | 0.51 | 0.58 | 0.71 | -0.19 | 0.16 | -2.13 | -1.43 |
| ACTBL2 | Q562R1 | 0.03 | -1.61 | -1.11 | -1.11 | -1.58 | -1.09 | -1.62 | -1.26 | -1.19 | -1.79 | -1.92 | -2.02 | -1.49 | -0.18 | -0.59 | 0.33 | 0.71 | 0.47 | -1.53 | 1.06 | -0.24 | -1.47 | -0.79 | 0.24 | 0.28 | -0.23 | 0.74 | 0.42 | 0.37 | -0.09 | -1.71 |
| ACTG1 | P63261 | 3.94 | 2.69 | 2.73 | 1.79 | 2.40 | 2.83 | 2.04 | 1.52 | 1.88 | 2.33 | 1.99 | 2.00 | 2.67 | 3.04 | 2.90 | 2.98 | 4.08 | 3.19 | 2.42 | 4.22 | 2.75 | 2.63 | 2.04 | 2.06 | 4.02 | 2.91 | 4.27 | 2.62 | 3.59 | 1.88 | 1.48 |
| ACTN1 | P12814 | 1.32 | 0.88 | -1.85 | 1.18 | -1.23 | -1.70 | -1.79 | -1.60 | 0.10 | -1.25 | -1.88 | -1.35 | -1.69 | 1.02 | 0.40 | 0.92 | 0.81 | 0.05 | 0.08 | 1.44 | 0.30 | 0.01 | 0.30 | -1.46 | 1.25 | 0.50 | 1.29 | -0.86 | 1.13 | -1.36 | -1.95 |
| ACTN4 | O43707 | 2.06 | 0.13 | -1.45 | -1.65 | -1.70 | 0.00 | -2.19 | -0.01 | 0.09 | 0.33 | -1.45 | -0.29 | -0.26 | 1.07 | 0.58 | 1.15 | 1.70 | 1.00 | -0.11 | 2.03 | 0.92 | 0.36 | -0.48 | 0.23 | 1.32 | 0.31 | 1.29 | 0.55 | 1.37 | 0.98 | -1.16 |
| ACTR2 | P61160 | 0.96 | 0.23 | 0.48 | -1.64 | -0.30 | 0.62 | -0.19 | -1.95 | -0.40 | -0.40 | 0.06 | -0.59 | -0.14 | 1.11 | -0.07 | 0.26 | 0.70 | 0.39 | 0.16 | 1.02 | -0.15 | -0.41 | -0.32 | -0.21 | 0.90 | -0.17 | 0.97 | 0.01 | 0.73 | -1.50 | -1.21 |
| ADH5 | P11766 | 0.93 | 0.16 | -1.50 | 0.97 | -1.76 | -1.63 | -1.56 | -1.41 | -0.30 | -1.68 | -1.42 | -1.10 | -0.78 | 0.63 | 0.40 | -0.19 | 1.19 | 0.48 | -0.42 | 0.82 | 0.22 | 0.15 | -1.76 | -0.22 | 0.41 | -0.04 | 1.02 | 0.16 | 0.17 | -1.49 | -1.55 |
| ADSL | P30566 | -0.42 | -1.29 | -1.87 | -1.59 | -1.34 | -1.60 | -1.24 | -1.73 | -1.47 | -1.58 | -1.35 | -1.62 | -1.33 | -0.32 | 0.96 | 0.04 | -0.78 | -1.30 | -1.31 | -0.16 | -1.66 | 0.40 | -1.59 | -1.13 | -0.20 | 0.85 | -0.62 | -1.64 | -0.23 | -1.77 | -1.97 |
| AHCYL2 | Q96HN2 | -0.16 | -1.47 | -1.12 | -2.30 | -0.82 | -1.23 | -1.13 | -1.86 | -1.68 | -1.31 | -1.29 | -1.26 | -1.22 | -0.72 | -1.03 | -0.11 | 0.29 | -0.37 | -1.68 | 0.30 | -0.23 | -1.28 | -1.61 | -2.01 | 0.01 | -1.87 | 0.05 | -0.28 | -0.74 | -0.74 | -1.48 |
| AK1 | P00568 | 0.95 | -0.53 | -1.25 | -1.04 | -1.26 | -1.30 | -0.40 | -1.26 | -1.62 | -1.88 | -1.08 | -1.54 | -1.87 | -0.14 | 1.90 | 1.04 | 0.79 | 0.82 | -1.82 | 0.85 | -0.77 | 1.51 | -1.55 | -0.50 | -0.96 | 1.81 | -0.35 | -0.06 | 1.09 | 0.26 | -2.04 |
| AKR1A1 | P14550 | 1.42 | 0.08 | -1.59 | -1.27 | -1.40 | -0.88 | -0.02 | -1.58 | -1.06 | -0.52 | -1.42 | -1.37 | -1.72 | 0.10 | 0.57 | 0.08 | 1.45 | 1.05 | -0.10 | 0.82 | -0.17 | 0.18 | -0.36 | -0.72 | 0.30 | 0.34 | 0.30 | -0.15 | 0.18 | -1.31 | -0.39 |
| AKR1E2 | Q96JD6 | 1.36 | -0.16 | -1.37 | -1.46 | -1.42 | -1.52 | -1.14 | -2.27 | -1.21 | -0.79 | -1.45 | -1.44 | -1.63 | 0.38 | 0.56 | 0.21 | 1.34 | 0.97 | -0.98 | 1.10 | 0.06 | 0.09 | -1.61 | -0.70 | 0.72 | 0.24 | 0.57 | 0.28 | 0.02 | -1.11 | -1.44 |
| ALDOC | P09972 | 0.90 | 0.18 | -0.93 | -1.27 | -0.49 | -1.06 | -0.43 | -2.12 | -1.36 | -0.27 | -0.42 | 0.12 | 0.01 | 0.54 | 0.42 | 0.43 | 0.39 | 0.07 | 0.52 | 1.03 | 1.22 | 0.38 | -0.12 | -0.39 | 0.84 | 0.09 | 0.81 | -0.39 | 0.66 | -0.21 | -0.78 |
| ANPEP | P15144 | -0.19 | 0.93 | -1.75 | -1.32 | 0.74 | -1.19 | 0.02 | -1.89 | -0.14 | -1.83 | 0.57 | -1.81 | 1.85 | 0.17 | -0.53 | -0.24 | 1.68 | 1.20 | -0.99 | 0.71 | 1.37 | -1.25 | 0.35 | -0.19 | 0.79 | -0.97 | 1.53 | 0.25 | -0.22 | 0.14 | -0.15 |
| ANXA1 | P04083 | -0.04 | 0.17 | 0.06 | -0.45 | -0.22 | 0.00 | 0.05 | -0.41 | 0.55 | 0.07 | -0.37 | -0.34 | 0.00 | 0.63 | 0.03 | 1.41 | 1.11 | 0.76 | -0.27 | 2.14 | 1.31 | -0.23 | -0.29 | 0.00 | 2.39 | -0.03 | 2.21 | 1.11 | 1.65 | -0.14 | -0.26 |
| ANXA3 | P12429 | 0.15 | -0.39 | -2.08 | -1.86 | 1.45 | -1.57 | -0.20 | -1.64 | 0.51 | -0.21 | -1.65 | -1.61 | -0.36 | 1.10 | -0.03 | 0.92 | 1.86 | 1.04 | -1.34 | 2.00 | 1.72 | -0.34 | -0.02 | -0.14 | 3.55 | 0.13 | 3.15 | 1.14 | 1.48 | -0.35 | -0.69 |
| ANXA4 | P09525 | 0.40 | 0.30 | -1.58 | -1.66 | 0.24 | -0.45 | -0.55 | -2.37 | -0.31 | -0.34 | -0.42 | -2.29 | 1.36 | 0.49 | 0.02 | 0.73 | 2.75 | 1.80 | -1.25 | 2.48 | 2.23 | 0.50 | -0.86 | 0.09 | 2.09 | -0.24 | 2.80 | 1.36 | 1.48 | -0.32 | -1.48 |
| ANXA7 | P20073 | -1.27 | -0.69 | -1.08 | -1.38 | -1.55 | -1.06 | -1.30 | -1.83 | -0.75 | -0.39 | -1.86 | -1.20 | -0.36 | -0.62 | 0.63 | 0.20 | 0.33 | -0.39 | -1.40 | 0.63 | 0.70 | 0.23 | -1.41 | -0.42 | 1.32 | 0.55 | 1.60 | -0.25 | 0.30 | 0.16 | -0.43 |
| AP2S1 | P53680 | 0.21 | -1.56 | -0.34 | -1.50 | -0.62 | -0.26 | -0.44 | -0.78 | -1.35 | -1.37 | -1.75 | -2.06 | -0.34 | 0.75 | 0.29 | -0.05 | 0.13 | -0.35 | -1.83 | 0.79 | -0.12 | -0.34 | -0.46 | -0.43 | 0.75 | -0.03 | 0.81 | 0.05 | 0.26 | -0.45 | -1.54 |
| APCS | P02743 | 1.13 | -0.54 | -2.19 | 0.56 | -1.83 | -1.58 | -1.74 | 1.25 | 1.90 | -0.87 | 1.89 | 1.99 | 1.61 | 1.66 | 1.42 | 1.90 | 0.54 | -0.10 | 1.38 | 1.11 | 1.30 | 1.72 | 0.46 | 0.98 | 1.87 | 1.43 | 1.11 | -0.26 | 1.83 | -0.33 | 0.15 |
| APEX1 | P27695 | 0.49 | -1.76 | -1.44 | 0.06 | -1.16 | -0.62 | -0.59 | -1.52 | -1.47 | -1.87 | -1.34 | -1.55 | -0.75 | 0.14 | -0.48 | -0.10 | 0.76 | -0.18 | 0.23 | 0.25 | -0.40 | -0.14 | -0.65 | -1.33 | 0.79 | -0.45 | 0.46 | -1.87 | 0.17 | -1.52 | -1.75 |
| APRT | P07741 | 0.86 | -0.40 | -0.86 | -1.74 | -1.78 | -1.52 | -1.22 | -1.60 | -0.37 | -0.99 | -1.24 | -1.46 | -1.81 | 0.30 | 0.95 | 0.39 | 1.15 | 0.39 | 0.12 | 1.38 | -0.07 | 0.59 | -1.87 | -0.56 | 1.16 | 0.79 | 0.96 | -0.04 | 0.53 | -0.54 | -0.90 |
| ARPC4 | P59998 | 1.36 | -0.24 | 0.00 | -0.66 | 0.08 | 0.10 | -0.22 | -1.29 | 0.18 | 0.08 | -0.51 | -0.17 | 0.09 | 1.43 | 0.26 | 0.85 | 1.12 | 0.49 | 0.32 | 1.52 | 0.37 | 0.43 | -0.24 | -0.07 | 1.49 | 0.00 | 0.93 | 0.42 | 0.93 | -0.50 | -1.58 |
| ARPC5 | O15511 | 1.09 | -1.55 | -1.02 | -1.57 | -2.46 | -1.33 | -1.62 | -1.49 | -0.21 | -1.01 | -1.57 | -1.33 | -1.01 | 2.48 | -0.06 | 0.43 | 0.89 | 0.15 | -0.19 | 1.21 | 0.22 | -0.64 | -1.07 | -0.32 | 2.05 | -0.08 | 2.06 | 0.08 | 0.63 | -1.76 | -2.07 |

| | | | | | | | | | | | | | | | | | | | | | | | | | | | | | | | | |
|---|---|---|---|---|---|---|---|---|---|---|---|---|---|---|---|---|---|---|---|---|---|---|---|---|---|---|---|---|---|---|---|---|
| ATIC | P31939 | 0.81 | -0.51 | -1.33 | -1.35 | -1.30 | -1.72 | -0.91 | -1.36 | -1.46 | -1.96 | -1.56 | -1.58 | -0.78 | -0.34 | 1.10 | 0.45 | 0.91 | 0.35 | -0.12 | 0.68 | -0.19 | 0.55 | -1.80 | -1.33 | 0.00 | 0.67 | 0.34 | -0.69 | 0.42 | -0.37 | -0.94 |
| AZU1 | P20160 | 1.94 | -1.55 | -1.74 | -1.07 | -1.27 | -1.49 | -1.59 | -1.74 | 2.13 | -1.05 | 0.52 | 0.71 | -0.16 | 3.47 | 0.40 | 1.87 | -1.12 | -0.34 | -1.43 | 2.43 | 3.38 | 0.27 | -0.24 | -1.47 | 4.72 | 0.96 | 5.06 | 1.77 | 2.45 | 0.50 | 0.00 |
| BCAP31 | P51572 | 0.21 | -1.27 | -1.36 | -1.91 | -1.77 | -1.65 | -1.76 | -1.36 | -1.31 | -1.46 | -2.07 | -1.15 | -1.16 | 0.10 | -1.67 | 0.00 | 0.21 | -0.40 | -1.55 | 0.57 | -1.65 | -1.81 | -2.51 | -1.80 | 0.38 | -1.36 | 0.72 | 0.04 | -0.65 | -1.55 | -1.77 |
| BLMH | Q13867 | 0.85 | 0.05 | 0.04 | -1.73 | 0.17 | 0.40 | 0.12 | -1.70 | 0.65 | -0.03 | 0.90 | 1.00 | 0.16 | 1.75 | 0.93 | 1.63 | 0.43 | 0.16 | -1.35 | 0.54 | 0.23 | 0.57 | 0.38 | 0.07 | 0.85 | 0.70 | 1.24 | 0.32 | 2.45 | 0.11 | -0.07 |
| BLVRA | P53004 | -0.25 | -0.19 | -0.89 | -1.93 | -1.72 | -1.80 | -1.52 | -1.37 | -1.52 | -1.28 | -1.51 | -1.63 | -1.51 | 0.76 | 1.21 | 0.26 | -0.54 | -0.20 | -0.25 | 0.14 | -1.55 | 0.50 | -1.84 | -1.49 | 0.45 | 0.95 | 0.62 | -1.64 | 0.21 | -0.91 | -1.02 |
| BPGM | P07738 | 0.79 | 0.29 | -1.70 | -1.91 | -0.88 | -1.48 | 0.07 | -0.86 | 0.03 | -1.06 | -1.61 | -1.79 | -1.50 | 0.69 | 2.35 | 1.48 | 0.49 | 1.03 | -1.47 | 0.86 | 0.20 | 2.01 | -1.91 | -1.22 | -0.80 | 2.32 | -0.65 | -1.67 | 1.69 | 1.03 | -0.63 |
| BPNT1 | O95861 | 0.38 | -1.93 | -0.06 | -1.70 | -1.18 | -1.32 | -1.43 | -1.47 | -1.59 | -1.42 | -0.97 | -1.16 | -1.41 | -0.19 | 0.03 | 0.23 | 0.68 | 0.08 | -1.31 | 0.47 | 0.08 | -0.40 | -1.88 | -0.62 | -0.70 | -0.13 | -0.50 | -1.29 | -0.34 | -1.68 | -2.13 |
| BROX | Q5VW32 | -0.03 | -0.26 | -1.30 | -1.81 | -1.78 | -1.35 | -1.79 | -1.60 | -1.52 | -0.26 | -1.54 | -1.39 | -1.59 | -0.01 | 0.56 | -0.27 | 0.20 | -0.14 | 0.90 | 0.34 | -0.50 | -0.12 | -1.68 | -0.32 | -0.09 | 0.33 | 0.14 | -0.84 | -0.21 | -1.02 | -1.59 |
| BST1 | Q10588 | -0.08 | -0.01 | -1.41 | 0.24 | -1.57 | -2.05 | -1.16 | -1.28 | -0.19 | -1.60 | -2.01 | -1.29 | -1.92 | 1.13 | -0.12 | 0.37 | -1.43 | -1.52 | -0.09 | 1.41 | 0.73 | 0.07 | -1.37 | -1.70 | 2.11 | -0.69 | 2.05 | -0.66 | 0.80 | -2.04 | -0.28 |
| BUB3 | O43684 | -0.24 | -1.60 | -1.47 | -1.88 | -1.85 | -1.68 | -2.10 | -2.14 | -1.23 | -2.28 | -1.78 | -1.80 | -0.82 | -0.40 | -1.78 | -0.41 | -0.23 | -1.45 | -1.18 | -0.02 | -1.58 | -1.41 | -1.98 | -1.16 | 0.21 | -1.10 | 0.22 | -1.55 | -1.50 | -1.37 | -1.87 |
| CACNA1G | O43497 | 0.86 | -1.77 | -1.55 | -1.20 | -1.18 | -1.57 | -1.49 | -1.47 | 0.69 | -1.98 | -1.80 | -1.21 | -1.32 | 1.73 | -1.31 | 0.84 | -1.53 | -1.61 | -1.91 | 1.95 | 1.37 | -1.88 | -1.54 | -0.87 | 3.56 | -0.04 | 3.76 | 0.84 | 1.36 | -0.59 | -1.63 |
| CALR | P27797 | 1.58 | -0.09 | 0.27 | -0.32 | 0.27 | 0.27 | 0.08 | -0.29 | 0.65 | 0.42 | -0.33 | -0.12 | 0.24 | 1.37 | 0.27 | 0.86 | 0.77 | 0.35 | -1.77 | 1.60 | 0.42 | -1.31 | -0.15 | 0.55 | 1.96 | -0.22 | 2.08 | 0.07 | 0.54 | -0.39 | -0.15 |
| CAMP | P49913 | 1.51 | -1.84 | -1.33 | -2.26 | -1.71 | -1.88 | -1.72 | 0.30 | 0.95 | -1.57 | 0.11 | 0.35 | 0.22 | 2.66 | 0.46 | 1.34 | -0.39 | -1.92 | 0.07 | 1.98 | 1.08 | 0.33 | -1.76 | -1.66 | 3.26 | 0.39 | 3.72 | 0.76 | 1.63 | -0.42 | -0.86 |
| CAP1 | Q01518 | 1.44 | -0.26 | -0.03 | -0.46 | -0.06 | 0.34 | 0.11 | -1.58 | 0.37 | -0.01 | -0.71 | -0.23 | 0.05 | 1.24 | 0.54 | 0.80 | 1.29 | 0.52 | 0.21 | 1.38 | 0.44 | -0.07 | -0.70 | -0.59 | 1.46 | 0.44 | 1.33 | 0.36 | 0.96 | -0.52 | -1.45 |
| CAPG | P40121 | 1.41 | 0.32 | -1.95 | -0.67 | -0.94 | -1.56 | -1.10 | -1.08 | -0.45 | -1.64 | -1.42 | -1.27 | -1.07 | 1.59 | -0.22 | 0.96 | 1.85 | 0.96 | -0.31 | 1.50 | 0.06 | -0.34 | -1.74 | -0.13 | 1.56 | -0.30 | 1.50 | 0.19 | 1.08 | 1.40 | -1.41 |
| CAPN1 | P07384 | 0.70 | -0.27 | -1.67 | -1.59 | -1.74 | -0.89 | -0.33 | -1.33 | 0.55 | -0.37 | -0.56 | -0.53 | -0.68 | 1.30 | 1.27 | 0.95 | 0.96 | 0.42 | -0.16 | 1.34 | 0.09 | 0.56 | -0.26 | -0.33 | 1.35 | 0.80 | 1.26 | 0.06 | 0.94 | 0.15 | -1.97 |
| CAPNS1 | P04632 | 0.61 | -1.35 | -1.56 | -0.69 | -0.58 | -1.73 | -1.73 | -2.06 | -0.10 | -1.59 | -0.59 | -1.68 | -0.87 | -0.13 | 0.56 | 0.37 | 0.79 | 0.23 | -1.60 | 0.70 | -0.03 | 0.00 | -1.65 | -0.73 | 0.18 | 0.04 | -0.14 | 0.17 | 0.27 | -1.09 | -1.67 |
| CBR1 | P16152 | 1.55 | 0.12 | -1.92 | -0.31 | -1.29 | -0.97 | -0.05 | -1.21 | -0.16 | -0.61 | -1.43 | -0.64 | -0.73 | 0.48 | 1.10 | 0.30 | 1.58 | 1.25 | 0.15 | 1.06 | 0.01 | 0.75 | -1.50 | -0.18 | 0.26 | 1.03 | 0.54 | 0.87 | 0.74 | -0.10 | -2.30 |
| CCS | O14618 | 0.18 | -1.08 | -1.48 | -1.76 | -0.76 | -1.86 | -1.28 | -1.66 | -1.31 | -1.62 | -1.52 | -1.79 | -1.63 | -0.06 | 1.15 | 0.24 | 0.11 | 0.08 | -1.29 | -0.06 | -0.35 | 0.86 | -1.92 | -1.67 | -0.32 | 1.09 | 0.01 | -1.99 | 0.53 | -0.59 | -1.22 |
| CD44 | P16070 | 0.75 | 0.52 | -1.98 | 0.72 | -0.51 | -0.52 | -0.51 | 0.10 | 0.89 | -0.53 | -0.48 | -0.57 | -0.45 | 0.68 | 0.54 | 1.25 | 0.45 | 0.10 | 0.88 | 1.00 | 0.29 | 0.83 | -1.84 | -0.43 | 0.33 | 0.50 | 0.17 | -0.30 | 1.07 | -1.35 | 0.56 |
| CD5L | O43866 | 1.31 | -0.25 | -1.92 | 0.87 | -1.60 | -1.12 | -1.34 | 2.02 | 1.23 | -0.36 | 0.24 | 0.11 | -0.29 | 1.43 | 1.77 | 2.18 | 0.17 | 0.33 | 2.07 | 1.01 | 1.22 | 2.01 | -1.50 | -1.63 | 1.65 | 1.63 | 0.22 | -0.34 | 1.76 | 0.38 | -0.06 |
| CD63 | P08962 | -0.02 | -0.21 | -1.74 | -1.17 | -1.61 | -0.12 | 0.07 | -1.33 | -0.17 | 0.69 | -0.92 | -0.79 | -2.44 | 0.55 | -0.68 | -0.39 | 0.48 | 0.60 | -1.91 | 1.79 | 0.47 | -1.83 | -0.32 | 0.46 | 1.49 | -1.81 | 2.16 | 0.62 | 0.39 | -0.77 | -1.36 |
| CDC42 | P60953 | 1.05 | 0.21 | 0.06 | -1.13 | -0.36 | -0.25 | -0.33 | -0.97 | 0.41 | -0.33 | -2.14 | -2.03 | -0.05 | 0.05 | 0.70 | 0.17 | 1.13 | 0.68 | 0.07 | 1.14 | 0.28 | 0.47 | -1.81 | -0.07 | 1.13 | 0.50 | 1.14 | 0.27 | 0.70 | -1.57 | 1.17 |
| CEACAM1 | P13688 | 0.33 | 0.49 | -0.54 | -1.63 | -0.76 | -0.43 | -1.80 | -1.71 | 0.01 | 0.68 | -0.04 | -0.07 | -1.48 | 0.53 | -1.06 | -1.30 | -0.05 | 0.05 | -1.99 | 1.43 | 1.35 | -1.28 | -1.62 | -0.18 | 2.09 | -1.66 | 2.38 | -0.10 | 0.51 | -1.51 | -1.75 |
| CEACAM5 | P06731 | 3.27 | 1.13 | 2.44 | -1.26 | -0.76 | 1.79 | 1.14 | -1.69 | 1.07 | 1.66 | 0.43 | 0.75 | 0.48 | 1.42 | 0.24 | 0.75 | 2.28 | 1.86 | -0.20 | 3.37 | 2.81 | -1.42 | 0.17 | 1.61 | 2.49 | 1.16 | 2.43 | 1.71 | 1.95 | 0.73 | -1.17 |
| CEACAM6 | P40199 | 1.08 | 1.41 | 1.44 | -1.28 | -1.44 | 0.88 | 0.09 | -1.95 | 0.68 | 0.39 | 0.00 | 0.32 | 1.16 | 1.76 | -0.77 | 0.11 | 1.31 | 0.47 | -1.70 | 2.46 | 3.00 | 1.48 | 0.04 | 1.20 | 2.89 | -0.46 | 3.83 | 1.03 | 1.69 | -0.01 | -1.53 |
| CEACAM7 | Q14002 | -0.12 | -1.67 | -1.30 | -2.12 | -1.75 | -1.49 | -1.36 | -1.15 | -1.80 | -1.57 | -1.83 | -1.39 | 0.32 | 0.24 | -1.62 | -1.45 | 0.86 | -0.10 | -1.59 | 0.99 | 0.94 | -1.68 | -1.30 | -1.59 | 1.47 | -1.48 | -0.03 | -1.31 | -0.27 | -1.60 | -0.80 |
| CEACAM8 | P31997 | -0.39 | -1.67 | -1.46 | 1.55 | -1.15 | -1.47 | -1.46 | -1.80 | 0.25 | -1.86 | -0.62 | -0.93 | -1.48 | 1.36 | -2.08 | -0.14 | 0.86 | 2.60 | -1.77 | 0.82 | 0.52 | -1.55 | -1.84 | -1.94 | 2.34 | 1.07 | 2.71 | 0.09 | 0.12 | -1.64 | -1.09 |
| CENPJ | Q9HC77 | 1.73 | 0.17 | -0.44 | -1.23 | 1.91 | -1.29 | -0.17 | -0.34 | 0.49 | -0.21 | -0.17 | -0.46 | 0.18 | 2.13 | -1.55 | 1.59 | 1.60 | 0.67 | -1.25 | 2.30 | 1.40 | -1.44 | 0.09 | 0.05 | 2.11 | -1.33 | 2.34 | 0.68 | 1.76 | 0.61 | -0.09 |
| CFL1 | P23528 | 2.26 | 0.75 | 0.66 | 0.28 | 0.42 | 0.53 | 0.38 | 0.29 | 0.25 | 0.58 | 0.41 | 0.31 | 0.72 | 1.87 | 1.71 | 1.44 | 2.12 | 1.63 | 0.93 | 2.23 | 1.34 | 1.22 | 0.51 | 0.67 | 2.07 | 1.48 | 2.20 | 1.24 | 1.59 | 0.51 | 0.03 |
| CHI3L1 | P36222 | 1.07 | 0.19 | -1.44 | 0.55 | -1.86 | -1.66 | -1.57 | -1.58 | 0.37 | -1.54 | -1.42 | -0.05 | -1.08 | 1.21 | -1.60 | 0.82 | -1.84 | -1.52 | 0.23 | 0.82 | -0.19 | -0.32 | 0.08 | -1.61 | 1.68 | -1.38 | 2.08 | -1.75 | 0.75 | -1.41 | 0.84 |
| CHIT1 | Q13231 | 0.55 | 0.87 | -1.55 | -0.96 | -0.92 | -1.13 | -1.99 | -0.22 | 0.00 | -1.66 | -2.12 | -1.23 | -1.72 | 1.45 | 1.01 | 0.56 | -1.22 | -1.44 | -1.52 | 0.47 | -1.50 | 0.03 | -1.39 | -1.21 | 1.66 | -1.67 | 2.12 | -0.90 | 0.00 | -2.02 | 1.73 |
| CLC | Q05315 | 0.70 | -1.30 | -2.09 | -1.68 | -2.06 | -1.55 | -1.87 | -1.78 | 0.56 | -1.32 | -1.92 | -1.65 | -1.87 | 0.54 | -1.00 | -0.77 | -1.54 | -1.10 | -1.89 | 0.06 | -1.38 | -1.34 | -1.67 | -1.65 | 0.30 | -0.23 | 1.06 | -2.34 | -0.41 | -1.70 | -1.75 |
| CLIC1 | O00299 | 2.01 | 1.00 | 0.27 | -0.08 | -0.03 | 0.18 | 0.34 | -0.19 | 0.65 | 0.51 | 0.03 | 0.04 | 1.01 | 2.23 | 1.35 | 0.93 | 2.46 | 1.75 | 0.35 | 2.25 | 1.48 | 0.84 | 0.15 | 0.46 | 2.04 | 1.23 | 2.22 | 1.15 | 1.31 | 0.39 | -0.35 |
| COPA | P53621 | 1.05 | -0.84 | -1.75 | -1.70 | -1.81 | -1.73 | -1.65 | -0.97 | -1.29 | -1.84 | -1.91 | 0.32 | -1.62 | 0.43 | 0.03 | -0.28 | 0.42 | 0.00 | -1.52 | 0.72 | 0.11 | -1.11 | -1.37 | -1.49 | 0.34 | -2.10 | 0.32 | 0.01 | -0.16 | -0.80 | -1.64 |
| COPB1 | P53618 | 1.29 | -1.43 | -2.13 | -1.94 | -1.60 | -1.10 | -2.10 | 0.62 | 0.11 | -1.91 | -1.27 | -1.75 | -1.95 | 0.32 | 0.16 | 0.33 | 0.58 | -0.17 | -1.72 | 1.04 | -1.67 | -1.99 | -1.82 | -1.75 | 0.54 | -1.29 | 0.46 | 0.28 | 0.29 | -1.39 | -1.92 |
| CORO1A | P31146 | 1.44 | -0.05 | -0.14 | -1.33 | -0.39 | -0.19 | -0.03 | -1.61 | -0.15 | 0.22 | -0.61 | -0.21 | -0.88 | 1.63 | 0.36 | 0.66 | 0.25 | -0.33 | 0.87 | 1.71 | 0.08 | -0.15 | -0.37 | -0.53 | 2.26 | 0.40 | 2.23 | 0.05 | 1.21 | -1.43 | -0.57 |
| CPPED1 | Q9BRF8 | -0.11 | -1.42 | -1.62 | -1.56 | -1.56 | -1.67 | -1.22 | -1.47 | -1.56 | -1.53 | -1.89 | -1.60 | -1.81 | 0.74 | -0.10 | -0.11 | 0.21 | -0.41 | -1.15 | 0.11 | -1.49 | -0.09 | -1.22 | -1.53 | 0.77 | 0.51 | 1.15 | -1.88 | 0.15 | -1.58 | -1.46 |
| CRISP3 | P54108 | 0.65 | -0.55 | 0.16 | 0.30 | -1.67 | -1.26 | -1.25 | -0.52 | 0.14 | -1.35 | -0.54 | -1.71 | 0.78 | 1.36 | -0.01 | 0.48 | 0.12 | -0.11 | 0.43 | 0.50 | -0.03 | 0.24 | -1.72 | -0.33 | 1.91 | -0.40 | 1.92 | -0.57 | 0.59 | -1.40 | 0.10 |
| CSTB | P04080 | 1.26 | 0.64 | -1.55 | -0.12 | -1.28 | -1.78 | -1.30 | -1.32 | 0.73 | -0.38 | -1.66 | -1.17 | -0.60 | 0.96 | 0.55 | 1.22 | 1.75 | 1.20 | -0.18 | 1.58 | 0.67 | -0.12 | -1.38 | -0.05 | 0.80 | 0.45 | 1.17 | 0.28 | 1.30 | -0.01 | 0.18 |
| CTSC | P53634 | 0.34 | 0.44 | -1.63 | -1.64 | -1.17 | -1.46 | -0.04 | -2.13 | 0.11 | 0.44 | 0.15 | -1.33 | -1.53 | 0.93 | 0.15 | 0.38 | 0.25 | 0.78 | -1.27 | 0.50 | 0.27 | -0.07 | -0.31 | -0.02 | 0.55 | 0.38 | 1.74 | 0.20 | 0.70 | -0.12 | -1.52 |
| CTSG | P08311 | 1.70 | -1.99 | -1.53 | -1.39 | -1.20 | -1.35 | -2.03 | 2.27 | 0.67 | -1.66 | -2.06 | -1.66 | -0.18 | 2.37 | 0.66 | 1.42 | -0.91 | -0.59 | -0.57 | 2.52 | 1.78 | 0.55 | -0.98 | 1.63 | 4.48 | 0.76 | 4.76 | 1.62 | 2.09 | 0.23 | 0.03 |
| CYBB | P04839 | 0.37 | -2.05 | -1.79 | -1.38 | -1.01 | -1.88 | -1.62 | -1.45 | 0.05 | -1.29 | -1.17 | 2.30 | -1.32 | 0.78 | -1.44 | 0.39 | -2.07 | -1.82 | -1.95 | 1.65 | 0.52 | -1.79 | -1.31 | -1.38 | 2.43 | -1.94 | 2.36 | 0.14 | 0.71 | -1.87 | -1.52 |
| CYCS | P99999 | 1.24 | -0.33 | -0.35 | -0.98 | -0.44 | -1.54 | -1.10 | -1.60 | -0.67 | -0.33 | -1.20 | -1.65 | -0.34 | 0.76 | 0.12 | 1.17 | 0.95 | 0.66 | -0.31 | 1.40 | 1.21 | -0.10 | -0.19 | -0.60 | 0.95 | -0.52 | 1.47 | -0.38 | 0.75 | -0.33 | -0.91 |
| DDB1 | Q16531 | -0.06 | -1.45 | -1.32 | -1.57 | -1.04 | -1.75 | -0.31 | -1.92 | -1.45 | -1.18 | -1.47 | -2.05 | -1.74 | -0.42 | 0.24 | -0.19 | -0.14 | -1.25 | -1.84 | 0.01 | -0.97 | -0.32 | -1.87 | -1.62 | 0.04 | -0.02 | -0.13 | -1.67 | -0.43 | -0.97 | -1.88 |
| DDT | P30046 | 0.75 | -0.59 | -1.50 | -1.64 | -1.30 | -1.76 | -0.60 | -1.65 | -1.43 | -1.56 | -1.60 | -1.33 | -1.65 | 0.10 | 1.29 | 0.83 | 0.90 | 0.53 | -1.14 | 0.54 | -0.66 | 0.70 | -1.47 | -1.36 | 0.44 | 1.14 | 0.04 | -0.64 | 0.71 | 0.24 | -1.85 |
| DDX17 | Q92841 | 0.61 | -0.97 | -0.61 | -1.38 | -0.77 | -1.33 | -0.83 | -1.49 | -0.40 | -1.10 | -1.99 | -0.52 | -1.30 | -0.27 | -0.46 | 0.13 | 0.29 | -0.74 | 0.13 | 0.29 | -0.25 | -0.58 | -1.86 | -0.85 | 0.35 | -0.42 | 0.42 | -0.65 | -0.13 | -1.49 | -1.12 |
| DDX39B | Q13838 | 0.95 | -0.31 | -0.22 | -1.39 | -0.33 | -0.27 | -0.23 | -0.66 | -0.51 | -0.16 | -0.78 | -1.12 | -0.31 | 0.51 | -0.07 | 0.36 | 0.96 | 0.25 | 0.16 | 1.08 | 0.14 | -0.38 | -0.58 | -0.31 | 1.01 | -0.29 | 0.88 | 0.05 | 0.18 | -0.48 | -0.92 |
| DDX55 | Q8NHQ9 | 0.83 | 0.49 | -1.47 | 1.27 | -1.37 | -1.44 | -1.83 | 0.59 | 1.15 | -1.49 | 1.36 | 0.88 | -0.67 | 2.62 | 0.57 | 1.04 | -1.90 | 0.02 | 1.13 | 1.17 | 0.86 | 1.10 | -0.09 | -2.03 | 1.47 | 0.62 | 2.02 | -2.06 | 1.28 | -1.23 | 1.41 |
| DEFA1 | P59665 | 3.91 | 1.16 | 0.81 | 0.39 | 0.51 | 0.86 | 0.97 | 0.25 | 3.71 | 1.02 | 0.64 | 0.73 | 1.62 | 5.32 | 2.39 | 4.11 | 1.49 | 1.53 | 0.91 | 5.52 | 4.84 | 2.09 | 1.30 | 1.30 | 6.74 | 2.57 | 7.31 | 3.57 | 4.64 | 2.28 | 1.89 |
| DEFA4 | P12838 | -1.01 | -1.36 | -1.10 | -2.03 | -1.63 | -0.86 | -1.32 | -1.21 | 0.32 | -1.59 | -1.84 | -1.22 | -1.62 | 0.82 | -1.34 | -1.38 | -1.73 | -1.26 | -1.53 | 1.22 | 0.39 | -1.47 | -1.78 | -1.73 | 3.21 | 0.86 | 3.20 | 1.65 | 0.50 | -1.41 | -1.40 |
| DMBT1 | Q9UGM3 | 0.13 | 1.98 | -1.13 | -1.41 | -1.70 | 3.83 | -0.74 | -2.08 | 0.36 | -1.63 | 0.37 | 0.41 | 1.33 | 1.87 | 0.44 | 0.08 | 0.98 | 0.76 | 1.81 | 4.43 | 4.53 | 0.78 | 1.25 | 0.91 | 2.82 | -0.12 | 3.56 | 0.55 | 2.91 | 0.04 | -0.20 |
| DNAJB11 | Q9UBS4 | 0.06 | -0.36 | -1.84 | -1.91 | -1.95 | -1.49 | -0.41 | -1.21 | -1.63 | -1.58 | -1.94 | -2.16 | -1.12 | -1.39 | 0.52 | -0.38 | -1.58 | 0.08 | -1.69 | 0.17 | 0.07 | -1.67 | -1.82 | -1.73 | 0.49 | -1.76 | 0.66 | -1.73 | -0.60 | -1.14 | -1.51 |
| DPP3 | Q9NY33 | 0.78 | -0.57 | -1.50 | -1.23 | -1.55 | -1.45 | -1.09 | -1.72 | -0.52 | -0.75 | -1.64 | -1.45 | -1.75 | 0.12 | 0.58 | 0.11 | 0.54 | 0.17 | -1.53 | 0.17 | -0.50 | 0.15 | -1.46 | -1.34 | 0.10 | 0.54 | 0.15 | 0.64 | 0.14 | -0.57 | -1.56 |
| EEF1B2 | P24534 | 1.21 | -0.97 | -0.82 | -1.43 | -0.63 | -0.71 | -0.75 | -1.14 | -1.02 | -1.86 | -0.97 | -1.91 | -1.87 | -0.01 | 0.02 | 0.30 | 0.79 | 0.37 | -0.24 | 0.55 | -0.71 | -0.44 | -0.47 | -0.23 | -0.07 | -0.71 | 0.13 | -0.18 | 0.01 | -0.90 | -1.88 |
| EEF1D | P29692 | 1.27 | -0.98 | -1.85 | -1.80 | -1.72 | -0.14 | -1.26 | -1.82 | -1.06 | -1.01 | -2.03 | -1.36 | -1.50 | 0.26 | 0.27 | 0.69 | 1.10 | 0.65 | 0.07 | 0.30 | -0.70 | 0.10 | -1.37 | -1.28 | 0.52 | 0.07 | 0.21 | -0.03 | 0.56 | -1.11 | -1.99 |
| EIF2S3 | P41091 | 0.40 | -1.41 | -1.37 | -1.61 | -1.87 | -1.16 | -1.42 | -1.66 | -0.18 | -0.80 | -1.93 | -1.67 | -1.80 | 0.11 | -0.01 | -0.12 | 0.14 | -0.53 | -1.17 | 0.19 | -1.01 | -0.16 | -1.80 | -1.77 | 0.06 | -0.08 | 0.24 | -0.52 | -0.36 | -1.11 | -1.43 |
| EIF6 | P56537 | 0.87 | -1.16 | -0.86 | -1.41 | -0.90 | -0.57 | -1.81 | -1.29 | -0.51 | -0.85 | -1.85 | -0.41 | -0.76 | -0.01 | 0.32 | 0.08 | 0.73 | 0.27 | -1.17 | 0.55 | 0.01 | 0.54 | -0.58 | -0.45 | 0.50 | 0.52 | 0.68 | -0.30 | 0.15 | -1.67 | -1.61 |

| | | | | | | | | | | | | | | | | | | | | | | | | | | | | | | | | |
|---|---|---|---|---|---|---|---|---|---|---|---|---|---|---|---|---|---|---|---|---|---|---|---|---|---|---|---|---|---|---|---|---|
| ELANE | P08246 | 2.53 | -0.50 | -1.78 | -1.83 | -1.49 | -1.72 | -0.62 | -1.40 | 1.78 | -0.38 | -1.54 | -0.64 | 0.80 | 3.67 | 1.19 | 2.03 | -0.01 | -0.08 | 0.06 | 3.19 | 2.64 | 0.74 | -0.78 | -0.45 | 5.21 | 1.47 | 5.17 | 1.99 | 2.49 | 0.77 | 0.44 |
| EML2 | O95834 | 0.66 | 0.07 | -1.76 | -1.67 | -1.61 | -2.09 | -1.50 | -0.94 | -2.18 | -1.75 | -1.11 | -1.33 | -1.38 | 0.39 | -0.34 | -0.39 | 0.48 | -0.06 | 0.06 | 0.34 | -0.05 | -0.39 | -0.16 | -1.80 | 0.14 | -0.46 | 0.12 | -0.68 | -0.12 | -2.30 | -2.05 |
| ENO1 | P06733 | 2.48 | 1.03 | 1.02 | 0.48 | 0.79 | 0.89 | 0.72 | 0.35 | 0.75 | 0.83 | 0.48 | 0.55 | 1.11 | 1.77 | 1.87 | 2.08 | 2.57 | 1.88 | 1.08 | 2.75 | 1.65 | 1.76 | 0.64 | 0.76 | 2.13 | 1.61 | 2.25 | 1.28 | 2.22 | 0.95 | 0.55 |
| EPB41 | P11171 | -0.34 | -0.17 | -1.42 | -2.02 | -1.38 | -1.29 | -1.13 | -1.74 | -1.45 | -1.56 | -1.84 | -1.28 | -1.79 | -0.07 | 1.57 | 0.56 | -0.53 | -0.15 | -1.10 | 0.10 | -0.12 | 1.15 | -2.09 | -1.46 | -0.58 | 1.35 | -0.27 | -1.98 | 0.74 | 0.17 | -2.03 |
| EPRS1 | P07814 | 0.71 | -0.11 | -0.16 | -1.66 | -1.21 | -1.51 | -1.71 | -1.32 | -0.34 | -1.30 | -0.84 | -1.04 | -1.27 | 0.08 | -0.37 | 0.08 | 0.30 | 0.09 | -0.68 | 0.21 | -0.06 | -0.49 | -1.69 | 0.04 | -0.06 | -0.66 | 0.12 | 0.50 | 0.04 | -1.80 | -1.57 |
| ERFL | A0A1W2PQ73 | 1.20 | 1.19 | -1.59 | 1.57 | -1.61 | -1.15 | -1.15 | 0.80 | -0.14 | -1.45 | -1.65 | -1.58 | -0.83 | 1.09 | 1.03 | 0.75 | 0.48 | 0.29 | 1.46 | 1.32 | 0.17 | 1.62 | -1.31 | -1.70 | 0.95 | 1.08 | 0.70 | -1.34 | 1.61 | -1.02 | 1.58 |
| ERP29 | P30040 | 1.86 | -0.61 | -1.53 | -2.22 | -1.14 | -0.95 | -0.94 | -1.84 | -0.36 | -0.80 | -1.05 | -1.60 | -1.27 | 0.69 | 0.16 | 0.41 | 0.59 | 0.54 | -0.33 | 0.38 | -0.45 | -0.20 | -1.57 | -0.84 | 1.28 | -0.32 | 1.25 | -1.61 | 0.29 | -0.26 | -1.43 |
| ETFA | P13804 | 0.47 | -1.43 | -1.73 | -1.51 | -1.14 | -0.56 | -2.06 | -0.39 | -1.66 | -1.64 | -1.68 | -1.41 | -1.41 | -0.45 | 0.66 | 0.12 | 0.61 | -0.09 | -1.49 | 1.30 | 0.04 | -0.64 | -1.86 | -0.03 | 0.86 | -1.50 | 0.86 | 0.44 | 0.52 | -1.31 | -1.80 |
| FABP3 | P05413 | -1.62 | 0.24 | -0.33 | -1.95 | -0.94 | -0.38 | -0.32 | -2.03 | -0.32 | -0.53 | -1.69 | -1.78 | -0.38 | -0.25 | -1.62 | 1.60 | -2.05 | -1.71 | -1.39 | 0.27 | -0.50 | -1.25 | 0.42 | -1.63 | -1.47 | -0.98 | -1.69 | -1.62 | -1.51 | -1.51 | -1.71 |
| FBP1 | P09467 | 0.63 | 0.13 | -1.31 | -1.42 | -1.55 | -1.84 | -1.44 | -1.38 | -0.97 | -1.99 | -1.66 | -1.31 | -1.97 | -1.18 | 0.34 | 0.60 | 1.42 | 0.99 | -1.47 | 0.99 | -0.62 | 0.06 | -1.68 | -0.26 | -0.04 | 0.38 | -0.07 | -0.56 | 0.55 | -1.90 | -1.81 |
| FCN1 | O00602 | -1.32 | -1.59 | -1.72 | -1.38 | -1.63 | -1.89 | -1.62 | -1.60 | 0.07 | -1.57 | -1.44 | -2.06 | -2.04 | -0.03 | -1.40 | 0.10 | -1.82 | -1.67 | -1.92 | -0.03 | -1.92 | -1.46 | -1.52 | -1.06 | 2.17 | -1.47 | 1.82 | -0.13 | 0.07 | -2.08 | -1.57 |
| FCN3 | O75636 | 0.50 | -1.82 | -1.88 | -0.86 | -1.24 | -1.47 | -1.37 | 0.50 | 0.49 | -1.40 | -2.21 | -1.71 | -1.37 | 0.65 | 0.56 | 1.05 | -0.56 | -0.29 | 0.81 | -0.09 | -0.66 | 0.21 | -1.32 | -2.04 | 0.53 | 0.67 | -1.78 | -1.49 | 0.90 | -1.29 | -1.80 |
| FERMT3 | Q86UX7 | 0.05 | -0.15 | -1.57 | -0.75 | -1.92 | -1.44 | -1.56 | -1.70 | -0.25 | -1.28 | -1.31 | -1.11 | 0.40 | 0.31 | 0.51 | 0.00 | -0.08 | -2.07 | -0.53 | 0.91 | -1.50 | -0.39 | -1.65 | -1.41 | 1.37 | 0.51 | 1.26 | -1.72 | 0.15 | -1.09 | -1.69 |
| FGR | P09769 | 0.70 | 0.52 | -1.40 | -1.72 | -1.49 | -1.23 | -1.89 | -1.39 | -1.79 | -1.46 | -1.16 | -1.05 | -0.22 | 0.76 | -1.95 | -0.03 | 1.61 | 1.07 | -0.20 | 1.45 | 0.68 | -1.34 | -2.27 | -1.43 | 2.19 | -2.13 | 2.02 | 0.78 | 0.93 | -2.03 | -1.26 |
| FKBP1A | P62942 | 1.15 | -0.09 | -2.00 | -1.79 | -1.15 | -1.75 | -1.26 | -1.23 | -1.59 | -1.90 | -1.77 | -2.18 | -0.98 | 0.23 | 0.99 | 0.86 | 1.27 | 0.76 | -0.13 | 0.98 | -0.30 | 0.76 | -1.29 | -0.85 | 0.63 | 0.91 | 0.44 | -0.69 | 0.87 | -0.08 | -1.96 |
| FLOT1 | O75955 | -0.55 | -0.44 | -1.60 | -1.35 | -1.84 | -1.32 | -1.37 | -1.97 | -1.76 | -1.71 | -1.50 | -1.76 | -1.69 | -1.62 | 0.79 | 0.44 | 0.26 | -0.17 | -0.21 | 1.07 | -0.32 | 0.55 | -1.56 | -1.04 | 0.28 | 0.65 | 0.27 | -0.65 | 0.54 | -0.77 | -1.26 |
| FLOT2 | Q14254 | -1.19 | -0.56 | -1.31 | -1.48 | -1.27 | -2.00 | -1.04 | -2.54 | -0.05 | -1.04 | -1.63 | -1.11 | -1.62 | -1.54 | 0.58 | 0.11 | 0.05 | -0.39 | -0.60 | 0.85 | -0.20 | 0.15 | -1.10 | -1.26 | 0.03 | 0.45 | 0.24 | -0.30 | 0.30 | -1.79 | -1.78 |
| GALE | Q14376 | 0.41 | 1.02 | -1.63 | -1.06 | -1.94 | -1.57 | -1.72 | 0.41 | -1.31 | -1.70 | -1.94 | -1.40 | -1.61 | 0.26 | -0.03 | 0.48 | 0.60 | 0.33 | -1.59 | 0.72 | -0.01 | -1.61 | -1.62 | 0.19 | -0.21 | 0.17 | -0.31 | -0.14 | -0.20 | -1.19 | -1.66 |
| GAPDH | P04406 | 2.99 | 1.45 | 1.52 | 0.86 | 1.40 | 1.68 | 1.22 | 0.70 | 1.01 | 1.30 | 1.12 | 1.10 | 1.57 | 2.81 | 2.66 | 2.32 | 2.72 | 2.01 | 1.35 | 3.33 | 2.02 | 2.36 | 1.33 | 1.23 | 2.98 | 2.51 | 3.10 | 1.63 | 2.76 | 1.44 | 0.66 |
| GBE1 | Q04446 | 0.12 | -1.48 | -1.74 | -1.46 | -1.29 | -1.42 | -0.70 | -1.33 | -0.24 | -1.59 | -2.03 | -0.91 | -1.61 | 0.64 | 0.00 | -0.24 | 0.01 | -1.28 | -1.68 | 0.40 | -1.29 | -0.20 | -0.93 | -1.89 | 0.71 | -0.11 | 0.68 | -1.76 | 0.12 | -1.48 | -1.45 |
| GC | P02774 | 3.03 | 3.06 | -1.72 | 3.25 | 0.37 | -0.46 | 0.96 | 2.56 | 2.32 | 0.96 | 0.17 | -0.61 | 2.58 | 2.70 | 2.70 | 3.22 | 2.29 | 2.12 | 2.89 | 3.21 | 1.65 | 3.25 | 0.22 | 0.37 | 2.39 | 2.47 | 1.90 | 0.74 | 3.32 | 1.12 | 3.21 |
| GCA | P28676 | -0.13 | -1.58 | -1.56 | 0.61 | -1.82 | -1.58 | -1.97 | -1.44 | 0.75 | -1.60 | -1.12 | -1.68 | -2.03 | 1.47 | -0.20 | 0.65 | -0.23 | -0.70 | -1.04 | 1.88 | 0.68 | -0.22 | 1.23 | -1.07 | 2.68 | -0.28 | 2.52 | 0.07 | 0.93 | -0.91 | -1.23 |
| GCLC | P48506 | -0.05 | -1.79 | -1.46 | -1.74 | -1.71 | -1.79 | -1.93 | -2.17 | -2.22 | -1.18 | -1.56 | -1.96 | -2.08 | -0.09 | 1.07 | 0.38 | -0.39 | -0.49 | -1.81 | -0.15 | -1.10 | 0.70 | -1.92 | -1.32 | -1.10 | 0.90 | 1.33 | -1.64 | 0.18 | -0.38 | -1.40 |
| GCLM | P48507 | -0.30 | -1.26 | -1.50 | -1.79 | -1.52 | -0.98 | -1.13 | -2.03 | -0.85 | -1.73 | -1.86 | -1.19 | -1.42 | -0.11 | 1.10 | 0.22 | -0.67 | -0.37 | -1.59 | 0.12 | -1.48 | 0.73 | -1.78 | -1.94 | 0.47 | 1.00 | 0.52 | -1.60 | 0.44 | -1.68 | -1.28 |
| GFUS | Q13630 | 1.65 | -1.61 | -1.73 | -1.52 | -1.73 | -1.57 | -0.72 | -1.60 | -1.45 | -0.34 | -1.63 | -1.57 | -1.71 | 0.65 | 1.37 | 0.60 | 1.22 | 0.80 | -1.20 | 1.36 | 0.37 | 0.74 | -0.33 | -0.18 | -0.03 | 1.32 | 0.33 | 0.21 | 0.62 | 0.46 | -1.46 |
| GLIPR2 | Q9H4G4 | -0.63 | -1.73 | -1.35 | -0.90 | -0.97 | -1.31 | -1.71 | -1.77 | -1.49 | -1.85 | -1.15 | -1.92 | -1.39 | 0.32 | 0.35 | 0.18 | -1.98 | -1.45 | -0.71 | 0.25 | -1.16 | 0.09 | 0.04 | -1.30 | 1.24 | 0.04 | 1.23 | -1.54 | 0.26 | -1.72 | -1.35 |
| GLO1 | Q04760 | 0.89 | -0.07 | -1.26 | -1.49 | -0.60 | -0.53 | -1.37 | -2.05 | -0.10 | -1.42 | 0.47 | 0.57 | -1.02 | 0.95 | 1.18 | 0.44 | 1.30 | 0.93 | -0.31 | 0.55 | 0.48 | 0.82 | -1.80 | -1.48 | 0.61 | 1.13 | 1.00 | -0.59 | 0.43 | 0.12 | -1.44 |
| GLOD4 | Q9HC38 | 0.99 | -0.42 | -1.74 | -2.00 | -1.71 | -1.59 | -2.03 | -1.05 | -0.88 | -0.93 | -1.54 | -1.17 | -1.30 | 0.84 | 0.87 | 0.38 | 1.29 | 0.76 | -0.10 | 0.34 | -0.23 | 0.57 | -1.56 | -0.31 | -0.64 | 0.72 | -0.35 | -0.48 | 0.36 | -0.55 | -1.82 |
| GMDS | O60547 | 1.83 | 0.07 | -1.46 | 0.22 | -1.85 | -0.92 | -0.08 | -1.60 | -1.29 | -1.27 | -1.79 | -2.02 | -1.95 | -0.25 | 0.41 | 0.37 | 1.47 | 0.89 | -1.75 | 1.26 | 0.36 | -0.43 | -1.16 | 0.09 | 0.04 | -0.34 | 0.49 | 0.73 | 0.19 | -1.47 | -1.32 |
| GMFG | O60234 | 0.43 | -2.30 | -1.61 | -1.12 | -1.27 | -1.88 | -1.03 | -1.91 | -1.57 | -1.55 | -1.63 | -1.96 | -1.50 | -0.14 | -0.16 | 0.03 | 0.22 | -0.55 | -2.02 | 0.82 | -1.74 | -0.02 | -1.47 | -1.68 | 1.23 | -0.54 | 1.13 | -1.47 | 0.13 | -1.25 | -1.84 |
| GNAI3 | P08754 | -0.55 | -0.74 | -1.16 | -1.72 | -1.82 | -1.89 | -1.26 | -1.35 | -1.80 | -0.42 | -2.32 | -1.73 | -1.76 | 0.48 | -0.54 | -0.35 | 0.21 | -0.14 | -1.46 | 0.78 | 0.57 | -1.67 | -1.17 | -0.58 | 1.19 | -0.50 | 1.26 | -0.24 | 0.03 | -1.01 | -1.33 |
| GNB2 | P62879 | -0.15 | 0.19 | -1.58 | -1.71 | -0.61 | -0.49 | -0.02 | -1.28 | -0.48 | 0.16 | -0.62 | -0.73 | 0.60 | -0.05 | -0.40 | -0.02 | 0.73 | 0.15 | -1.48 | 1.22 | 0.78 | -0.28 | 0.14 | 0.15 | 1.18 | -0.51 | 1.16 | 0.10 | 0.57 | -1.63 | -1.31 |
| GNPDA1 | P46926 | 0.30 | -1.55 | -1.42 | -1.52 | -0.93 | -1.21 | -1.61 | -1.80 | 0.29 | -1.88 | -1.11 | -1.51 | -1.43 | -0.28 | 0.49 | 0.66 | -0.05 | -0.65 | -1.65 | 0.13 | -1.75 | 0.28 | -1.67 | -1.76 | 0.20 | 0.36 | -0.23 | -1.92 | 0.28 | -1.78 | -2.29 |
| GPD2 | P43304 | -1.59 | -0.09 | -0.74 | -1.48 | -1.49 | -1.42 | -1.89 | -1.50 | -1.33 | -0.08 | -1.88 | -1.19 | -1.76 | 0.33 | -1.39 | -0.28 | -0.09 | -0.51 | -1.39 | 0.79 | -0.30 | 0.51 | -1.66 | -1.10 | 0.28 | -0.11 | 0.70 | -0.24 | -0.12 | -1.59 | -1.87 |
| GPRC5A | Q8NFJ5 | -1.12 | 1.00 | -1.61 | -2.03 | -1.41 | -0.82 | -0.21 | -1.55 | -1.71 | 0.48 | -1.76 | -1.67 | 0.46 | -0.27 | -0.41 | 0.00 | 0.71 | -0.19 | -1.70 | 1.91 | 0.69 | -1.33 | -1.22 | 0.41 | 0.06 | -1.01 | 1.53 | -1.78 | 1.15 | -1.70 | -1.78 |
| GPX1 | P07203 | -0.04 | -1.31 | -1.86 | -1.79 | -1.29 | -1.49 | -1.58 | -1.76 | -1.83 | -1.01 | -1.73 | -1.91 | -1.60 | 0.68 | 1.11 | 0.21 | 0.64 | 0.13 | -1.30 | 0.46 | -0.40 | 0.60 | -1.47 | -1.21 | -0.26 | 0.85 | 0.31 | -1.32 | 0.38 | -0.09 | -1.51 |
| GSR | P00390 | 1.19 | -0.05 | -1.74 | -0.45 | -0.80 | -0.89 | -1.24 | -1.46 | 0.01 | -0.62 | -0.27 | -0.41 | -0.58 | 0.76 | 1.09 | 1.19 | 1.61 | 0.85 | 0.19 | 1.17 | 0.71 | 0.65 | -0.77 | -0.05 | 1.18 | 0.98 | 1.68 | 0.08 | 0.93 | -0.19 | -0.46 |
| GSS | P48637 | 1.68 | -0.03 | -1.12 | -1.79 | -1.13 | -0.88 | -0.10 | -2.13 | -2.00 | -0.23 | -1.42 | -0.95 | -1.75 | 0.84 | 1.49 | 1.09 | 1.23 | 0.86 | 1.77 | 1.18 | 0.81 | 0.94 | -1.08 | 0.13 | 0.99 | 1.31 | 1.06 | 0.42 | 1.02 | 0.32 | -1.68 |
| GSTO1 | P78417 | 1.62 | 0.58 | -1.40 | -1.10 | -0.37 | -0.85 | 0.08 | -0.64 | 0.19 | -0.27 | -1.53 | -0.25 | 0.03 | 1.56 | 2.21 | 1.30 | 1.62 | 0.74 | -0.34 | 1.77 | 0.91 | 1.51 | -0.57 | -0.22 | 1.64 | 1.95 | 1.55 | 0.23 | 1.36 | 0.52 | -0.59 |
| GSTP1 | P09211 | 2.65 | 0.75 | -0.45 | -0.46 | -0.97 | -0.66 | -0.13 | -0.65 | -0.02 | 0.21 | -1.22 | -1.00 | 0.41 | 2.09 | 1.19 | 1.48 | 2.65 | 2.03 | -0.20 | 2.45 | 1.37 | 0.97 | -0.57 | 0.32 | 2.16 | 1.16 | 2.17 | 1.29 | 1.50 | 0.10 | -0.18 |
| GUSB | P08236 | 0.79 | -0.39 | 0.08 | -1.13 | -1.55 | -1.57 | 1.35 | -2.33 | 0.07 | -1.67 | -0.46 | -0.61 | -1.02 | 1.39 | -0.95 | -0.64 | 0.72 | 0.68 | -1.71 | 0.71 | 0.82 | 0.55 | -1.67 | -1.59 | 1.48 | -1.35 | 2.04 | -1.15 | -0.35 | -1.44 | -2.15 |
| GYG1 | P46976 | 0.27 | -1.68 | -1.77 | -1.04 | -1.48 | -1.52 | -1.83 | -1.02 | -0.16 | -1.50 | -1.67 | -1.56 | -2.02 | 1.77 | -0.07 | 0.27 | -0.09 | -2.05 | -1.09 | 0.62 | -1.79 | 0.21 | -1.87 | -1.52 | 1.02 | -1.12 | 1.04 | -1.32 | 0.58 | -1.42 | -1.46 |
| H1-0 | P07305 | 1.12 | -1.59 | -2.08 | -1.80 | -1.54 | -1.75 | -1.57 | -1.36 | -1.81 | -1.54 | -1.35 | -1.06 | -1.62 | -0.46 | -0.18 | 0.27 | 1.57 | 0.54 | -0.59 | 0.52 | -1.60 | -0.31 | -1.58 | -2.24 | -0.81 | -0.47 | -1.18 | -1.53 | 0.22 | -1.24 | -1.07 |
| H1-5 | P16401 | 1.38 | -0.80 | -0.44 | -1.84 | -0.18 | -0.80 | -0.58 | -1.78 | -1.00 | -1.30 | -0.89 | -1.30 | -0.62 | -0.80 | 0.32 | 0.88 | 1.02 | 0.40 | 0.57 | 1.76 | -0.70 | -0.44 | -0.44 | -0.59 | -0.18 | 0.30 | 0.22 | -0.63 | 1.40 | -0.83 | -1.79 |
| H2AC11 | P0C0S8 | 3.47 | 1.75 | 1.91 | 1.23 | 1.67 | 1.99 | 1.52 | 0.91 | 1.07 | 1.69 | 1.36 | 1.42 | 2.07 | 2.00 | 1.83 | 3.21 | 3.57 | 2.61 | 1.87 | 3.99 | 1.65 | 1.54 | 1.56 | 1.53 | 4.59 | 1.58 | 4.16 | 2.19 | 3.71 | 1.40 | 1.04 |
| H2BC12 | O60814 | 3.45 | 1.46 | 1.53 | 0.79 | 1.41 | 1.80 | 1.37 | 0.75 | 1.47 | 1.60 | 1.17 | 1.27 | 1.92 | 3.10 | 1.75 | 3.09 | 3.25 | 2.28 | 1.70 | 4.13 | 1.64 | 1.35 | 1.58 | 1.54 | 4.82 | 1.62 | 4.55 | 2.07 | 3.55 | 1.40 | 0.74 |
| H2BU1 | Q8N257 | 1.28 | 0.31 | 0.41 | 0.00 | 0.28 | 0.50 | 0.24 | -0.25 | 0.54 | 0.38 | 0.09 | 0.16 | 0.44 | 1.45 | 0.02 | 1.28 | 1.09 | 0.28 | -0.01 | 2.19 | 0.36 | -0.09 | 0.15 | 0.28 | 3.12 | 0.00 | 3.08 | 0.71 | 1.25 | 0.11 | -0.19 |
| H3-2 | Q5TEC6 | 1.06 | -1.70 | -1.60 | -1.69 | -1.48 | -1.48 | -1.45 | -0.20 | -0.66 | -1.33 | -1.32 | -1.79 | -1.77 | -0.17 | -0.41 | 1.19 | 0.81 | -0.19 | -0.64 | 1.68 | -0.28 | -0.63 | -1.08 | -1.15 | 1.68 | -0.50 | 0.71 | -0.25 | 1.22 | -0.59 | -1.31 |
| H3-3A | P84243 | 3.27 | 1.04 | 1.44 | 0.63 | 1.12 | 1.64 | 1.20 | 0.41 | 1.09 | 1.40 | 0.96 | 0.91 | 1.79 | 2.43 | 1.54 | 3.42 | 2.99 | 1.84 | 1.45 | 3.99 | 1.91 | 1.22 | 1.24 | 1.27 | 3.99 | 1.49 | 3.75 | 2.02 | 3.59 | 1.16 | 0.69 |
| H4C1 | P62805 | 3.24 | 1.63 | 1.87 | 1.10 | 1.69 | 2.04 | 1.57 | 0.89 | 1.39 | 1.76 | 1.31 | 1.42 | 2.14 | 2.22 | 1.75 | 3.41 | 3.12 | 1.99 | 1.39 | 4.23 | 1.85 | 1.25 | 1.36 | 1.47 | 4.30 | 1.57 | 4.35 | 2.26 | 3.45 | 1.39 | 0.96 |
| HK2 | P52789 | -0.07 | -1.51 | -1.70 | -2.11 | -0.93 | -1.66 | -2.11 | -1.87 | -1.44 | -1.40 | -1.21 | -1.00 | -1.34 | 0.03 | -0.78 | -0.65 | -0.17 | 0.42 | -1.49 | 0.68 | 0.15 | -1.76 | -0.99 | -0.97 | 0.13 | -1.32 | 0.14 | -1.11 | -0.05 | -0.01 | -1.55 |
| HK3 | P52790 | 0.32 | -1.85 | -1.91 | -1.35 | -1.65 | -1.96 | -1.83 | 0.56 | 0.02 | -1.62 | -1.64 | -1.47 | -1.81 | 1.10 | -0.44 | 0.03 | -1.63 | -1.59 | -1.18 | 0.80 | -1.61 | -1.67 | -1.32 | -1.66 | 1.24 | -1.51 | 0.95 | -1.78 | 0.35 | -1.73 | -2.34 |
| HLA-A | P04439 | 0.67 | -1.48 | -1.73 | -0.32 | -1.69 | -1.58 | -1.31 | -0.47 | -0.44 | -1.15 | -1.79 | -1.51 | -1.55 | 0.00 | -0.60 | 0.23 | 0.01 | -0.32 | -0.08 | 1.05 | 0.56 | 0.61 | -1.33 | 0.28 | 1.37 | -0.49 | 1.90 | 0.05 | 0.47 | -1.90 | -1.81 |
| HLA-DRB3 | P79483 | 0.26 | -0.57 | -0.79 | -1.34 | -0.81 | -0.82 | 0.00 | -1.46 | -0.36 | -2.16 | -1.59 | -1.69 | -0.34 | -1.02 | 0.40 | 0.35 | -0.27 | -0.93 | -1.44 | 0.35 | 0.23 | -1.99 | -0.25 | -1.82 | -0.41 | -0.59 | 0.48 | -2.01 | -1.52 | -1.60 | -1.60 |
| HMGB1 | P09429 | 1.64 | -1.77 | 0.19 | -1.80 | -1.48 | -1.71 | -1.89 | -1.54 | 0.75 | -1.79 | -1.75 | -1.71 | -1.70 | 1.81 | 0.78 | 1.13 | 2.04 | 0.99 | 0.90 | 1.40 | -0.92 | 0.39 | -1.29 | -1.49 | 1.16 | -1.73 | 0.81 | -1.53 | 1.17 | -1.35 | -1.67 |
| HMGB2 | P26583 | 0.56 | -1.01 | -0.84 | -1.59 | -1.75 | -1.56 | -1.33 | -1.46 | 0.12 | -1.21 | -1.77 | -1.69 | -1.69 | 1.26 | -0.70 | 0.14 | -0.08 | -0.24 | -0.26 | 0.93 | -1.60 | -0.83 | -1.82 | -1.60 | 0.73 | -0.39 | 0.45 | -0.67 | 0.45 | -1.52 | -1.69 |
| HMGB3 | O15347 | -0.31 | -1.77 | -1.37 | -1.38 | -1.88 | -1.20 | -1.41 | -1.34 | -0.42 | -1.82 | -1.31 | -1.58 | -1.54 | 0.39 | -0.45 | -0.20 | -0.91 | -0.85 | -0.75 | -0.61 | -1.03 | -1.62 | -1.38 | -1.49 | 0.39 | -1.74 | -0.03 | -1.71 | -0.61 | -1.88 | -1.52 |
| HNRNPC | P07910 | 0.66 | -0.75 | -0.38 | -2.01 | -0.47 | -0.27 | -0.51 | -0.73 | -0.29 | -0.55 | -0.87 | -1.87 | -0.50 | 0.56 | -0.28 | 0.69 | 0.67 | -0.04 | 0.83 | 1.24 | -0.03 | -0.14 | -0.31 | -0.32 | 0.84 | -0.23 | 0.83 | -0.14 | 0.27 | -0.71 | -1.69 |

| | | | | | | | | | | | | | | | | | | | | | | | | | | | | | | | | |
|---|---|---|---|---|---|---|---|---|---|---|---|---|---|---|---|---|---|---|---|---|---|---|---|---|---|---|---|---|---|---|---|---|
| HNRNPD | Q14103 | 0.95 | -1.27 | -1.69 | -1.42 | -1.67 | -1.62 | -1.85 | -1.02 | -1.58 | -1.15 | -1.47 | -1.46 | -1.69 | 1.13 | -0.08 | 0.27 | 1.06 | 0.15 | 0.26 | 0.43 | -1.44 | -0.45 | -1.51 | -1.64 | 0.50 | -0.62 | 0.57 | -1.61 | -0.14 | -1.46 | -1.64 |
| HNRNPF | P52597 | 0.37 | -0.46 | -1.42 | -1.40 | -1.89 | -1.50 | -1.34 | -1.77 | -1.32 | -1.63 | -1.36 | -1.81 | -1.81 | -0.59 | -0.24 | -0.40 | 0.42 | -0.40 | -0.27 | 0.58 | 0.57 | -1.22 | -0.91 | -2.15 | 0.11 | -0.61 | 0.40 | -1.43 | -0.71 | -1.39 | -1.34 |
| HNRNPUL2 | Q1KMD3 | 0.27 | -0.04 | -1.48 | -1.45 | -1.44 | -1.24 | -1.18 | -1.65 | -1.18 | -1.57 | -1.60 | -1.96 | -1.00 | 0.38 | -1.98 | -0.10 | 0.21 | -1.90 | -0.66 | 0.69 | -1.70 | -1.26 | -1.26 | -1.19 | 0.60 | -1.33 | 0.63 | 0.17 | -0.14 | -1.62 | -1.55 |
| ICAM3 | P32942 | -0.64 | -1.94 | -1.29 | -1.37 | -1.93 | -1.28 | -1.34 | -1.39 | -0.09 | -1.48 | -1.68 | -1.74 | -1.28 | 0.09 | -1.38 | 0.21 | 0.32 | -1.28 | 0.20 | 0.53 | -0.72 | -0.76 | -1.67 | -1.57 | 0.87 | -0.93 | 1.18 | -1.89 | 0.15 | -1.85 | -1.24 |
| ILF3 | Q12906 | 0.74 | -1.12 | -1.49 | -1.51 | -1.57 | -1.35 | -1.79 | -1.52 | 0.55 | -1.53 | -1.25 | -1.46 | -0.60 | 0.03 | -0.28 | 0.37 | 0.44 | -0.41 | -0.10 | 0.21 | -1.04 | -0.47 | -1.49 | -0.35 | -0.10 | -1.43 | -0.16 | -1.29 | -0.48 | -1.72 | -1.38 |
| IQGAP1 | P46940 | 1.83 | 0.56 | -1.47 | -1.63 | 1.09 | -1.43 | -2.16 | 1.47 | -1.69 | 0.75 | -1.73 | -0.04 | -1.86 | 1.23 | 0.12 | 1.15 | 1.53 | 0.90 | 0.53 | 1.69 | 0.70 | 1.04 | -0.03 | -1.29 | 1.31 | 0.63 | 1.19 | 0.75 | 1.22 | -2.22 | -1.95 |
| IST1 | P53990 | 0.20 | -0.25 | -0.67 | -1.60 | -1.38 | -1.54 | -0.14 | -1.57 | -1.41 | 0.04 | -1.03 | -1.03 | 0.27 | 0.33 | 0.40 | -0.25 | 0.61 | 0.64 | -0.62 | 0.89 | 0.19 | -0.03 | -1.41 | 0.65 | -0.11 | 0.30 | 0.28 | -0.18 | 0.31 | -1.69 | -1.11 |
| ITGB2 | P05107 | 0.03 | 0.21 | -1.88 | -1.79 | -1.92 | -1.64 | -1.46 | -1.85 | 0.65 | -1.25 | 1.11 | -1.63 | -1.23 | 1.40 | -1.50 | 0.71 | -0.92 | -1.51 | 0.39 | 1.82 | 1.49 | -1.71 | -0.20 | -1.70 | 3.17 | -0.16 | 3.58 | 0.69 | 1.04 | -0.30 | -1.24 |
| LAMP1 | P11279 | 0.41 | -0.20 | -1.90 | -0.27 | -1.91 | -0.23 | -0.28 | -0.61 | 0.18 | -0.19 | -0.25 | -0.04 | -1.95 | 0.42 | -0.42 | 0.27 | 0.63 | 0.28 | -0.21 | 1.22 | 1.32 | -0.47 | 0.15 | -0.07 | 1.66 | -0.44 | 2.24 | 0.59 | 0.90 | -1.36 | -0.10 |
| LAP3 | P28838 | 2.23 | 1.00 | 0.82 | -0.66 | 0.91 | 0.73 | -1.42 | -0.40 | 0.86 | -0.90 | -1.28 | 0.44 | -1.62 | 1.48 | 1.30 | 1.31 | 2.31 | 1.61 | -0.88 | 2.29 | 1.58 | 1.28 | 1.14 | -1.98 | 1.78 | 1.10 | 2.20 | 0.67 | 1.47 | 1.23 | -1.64 |
| LASP1 | Q14847 | 0.97 | 0.35 | -1.21 | -1.23 | -1.07 | -1.62 | -1.61 | -1.47 | -1.61 | -1.26 | -1.09 | -0.80 | -0.47 | -0.14 | 0.59 | -0.12 | 1.39 | 1.11 | -0.37 | 0.59 | 0.27 | 0.40 | -1.84 | -1.62 | 0.87 | 0.37 | 0.89 | -0.48 | 0.40 | -1.03 | -0.03 |
| LCN2 | P80188 | 2.72 | 3.79 | 0.00 | -0.54 | 0.99 | 1.27 | 2.15 | -1.06 | 1.88 | 2.71 | 0.53 | 0.41 | 3.60 | 3.76 | 0.83 | 1.55 | 1.89 | 2.65 | -1.85 | 2.49 | 2.54 | 0.08 | 1.22 | 2.37 | 3.67 | 1.29 | 3.77 | 1.95 | 1.90 | 0.92 | 0.25 |
| LCP1 | P13796 | 2.02 | 0.23 | -1.45 | 0.53 | -1.66 | 0.49 | -1.21 | -0.11 | 0.92 | -1.92 | -1.55 | 1.23 | 0.76 | 1.81 | 1.08 | 1.92 | 0.55 | 0.17 | 1.01 | 2.32 | 0.79 | 0.75 | 0.11 | 0.66 | 2.21 | 0.92 | 2.08 | -0.08 | 1.98 | -1.81 | 0.27 |
| LDHA | P00338 | 2.57 | 1.29 | 1.53 | 0.79 | 1.28 | 1.66 | 1.16 | 0.64 | 1.02 | 1.40 | 0.95 | 1.08 | 1.60 | 2.02 | 1.85 | 2.25 | 2.19 | 1.37 | 0.88 | 3.15 | 1.94 | 1.50 | 1.09 | 1.20 | 2.30 | 1.52 | 2.25 | 1.44 | 2.46 | 1.02 | 0.64 |
| LGALS1 | P09382 | 0.85 | 0.03 | -1.97 | 0.36 | -2.07 | -0.87 | -1.86 | -0.46 | -0.41 | -1.17 | -0.17 | -1.65 | -1.87 | -0.13 | -0.12 | 0.81 | 1.10 | -0.22 | -0.22 | 1.29 | -0.62 | 0.26 | -1.43 | -1.80 | 0.58 | -0.60 | 0.65 | -0.18 | 0.98 | -1.71 | -0.40 |
| LGALS3 | P17931 | 1.43 | 0.77 | -1.80 | -0.29 | -1.43 | -0.76 | -0.12 | -0.58 | -1.55 | -0.18 | -0.33 | -1.18 | 0.07 | -1.09 | 0.41 | 1.34 | 2.45 | 1.63 | -0.15 | 1.74 | 1.63 | 0.63 | -0.21 | 0.83 | 0.80 | 0.30 | 0.71 | 1.15 | 1.75 | 0.09 | -0.68 |
| LPGAT1 | Q92604 | 0.87 | 0.48 | -1.55 | -1.38 | -1.72 | -1.18 | 1.05 | -1.39 | -1.70 | -1.55 | -2.02 | -1.23 | -1.15 | 0.40 | 1.18 | 1.05 | -2.03 | -1.63 | -1.47 | 0.40 | 1.05 | 0.88 | -1.15 | -1.38 | 1.18 | 1.25 | 1.28 | -1.86 | 1.00 | 0.87 | -1.55 |
| LTA4H | P09960 | 1.18 | 0.79 | -1.65 | -1.29 | -0.73 | -1.26 | -1.46 | -1.95 | 0.22 | -1.44 | 0.79 | 0.81 | -0.29 | 1.29 | 0.85 | 0.84 | 0.80 | 0.34 | 0.46 | 1.28 | 0.36 | 0.36 | 0.93 | 0.99 | 1.24 | 0.71 | 1.02 | -1.66 | 0.96 | -1.91 | -0.85 |
| LTF | P02788 | 2.89 | 0.03 | -0.28 | 0.10 | -0.04 | -0.14 | 0.03 | 3.65 | 2.12 | 0.19 | 0.07 | 0.46 | 1.16 | 3.71 | 0.90 | 2.48 | 0.26 | 0.33 | -0.04 | 3.50 | 3.45 | 0.94 | 0.43 | 0.25 | 4.81 | 1.29 | 5.36 | 2.13 | 3.03 | 0.88 | 0.44 |
| MACROH2A1 | O75367 | 1.73 | -1.46 | -1.83 | -1.79 | -1.31 | -1.52 | -1.90 | -1.55 | -1.96 | -1.49 | -1.68 | -1.42 | -1.83 | 1.39 | -0.10 | 1.27 | 1.63 | 0.80 | 0.70 | 1.48 | 0.11 | -0.11 | 0.04 | -2.18 | 1.64 | 0.03 | 1.20 | 0.75 | 1.76 | -0.12 | -1.66 |
| MAP2K1 | Q02750 | -0.20 | -0.22 | -0.91 | -0.59 | -0.66 | -2.04 | -1.74 | -2.15 | -1.36 | -1.69 | -1.72 | -0.52 | -1.64 | -0.01 | -0.13 | -0.42 | -0.21 | -1.39 | -1.61 | 0.36 | 0.35 | -0.53 | -1.10 | -1.67 | 0.53 | -0.25 | 0.42 | -0.61 | -0.49 | -1.80 | -1.49 |
| MAPK15 | Q8TD08 | 0.53 | -0.91 | -0.61 | -1.22 | -1.11 | -0.79 | -0.72 | -1.36 | -0.67 | -0.97 | -0.78 | -1.91 | -1.73 | 0.55 | 0.05 | -0.14 | -1.97 | -0.10 | -0.69 | 0.80 | -0.17 | -0.25 | -0.83 | -1.32 | 0.85 | 0.03 | 0.78 | 0.20 | 0.14 | -0.64 | -1.84 |
| MMP9 | P14780 | 1.83 | 0.24 | -1.95 | 0.68 | -1.83 | -1.25 | -1.44 | 1.41 | 0.61 | -1.81 | 0.43 | -2.24 | -1.38 | 2.40 | 0.30 | 1.60 | 0.69 | 0.80 | 0.39 | 1.26 | 0.57 | 0.05 | -1.35 | 0.13 | 2.65 | 0.24 | 3.29 | 0.27 | 1.23 | 1.42 | -1.93 |
| MPO | P05164 | 2.35 | -1.91 | -1.46 | -1.65 | 2.74 | -1.15 | -1.58 | 2.50 | 1.73 | -0.28 | -0.26 | -0.27 | 0.20 | 3.16 | 0.61 | 2.29 | -0.57 | -0.35 | 1.55 | 3.29 | 2.90 | 0.41 | -1.42 | -0.36 | 4.67 | 0.88 | 5.00 | 1.75 | 2.78 | 0.53 | -0.03 |
| MSN | P26038 | 1.81 | 1.64 | 0.12 | 0.19 | 0.16 | -0.11 | 0.65 | -0.25 | 0.31 | 1.30 | -0.31 | 0.06 | 1.32 | 1.90 | 1.31 | 1.65 | 2.02 | 1.23 | 1.09 | 2.44 | 1.61 | 0.86 | -0.15 | 1.04 | 2.01 | 1.18 | 1.97 | 1.23 | 1.85 | 0.45 | 0.05 |
| MUC5B | Q9HC84 | 0.04 | 2.87 | 0.90 | 0.06 | 0.39 | 1.97 | 0.89 | -1.77 | 0.63 | 0.49 | 0.19 | -1.36 | 2.62 | 1.46 | 1.67 | -0.42 | 1.37 | 1.51 | -1.63 | 3.85 | 1.73 | 1.13 | 0.23 | 1.59 | 2.03 | 1.35 | 1.69 | 1.15 | 2.74 | -0.20 | 0.32 |
| MVP | Q14764 | 1.54 | -1.93 | -2.08 | -0.46 | -1.20 | -1.08 | -1.49 | -1.41 | -0.80 | -0.26 | -1.00 | -2.14 | -1.61 | 0.08 | -0.12 | 0.19 | 1.39 | 0.69 | -0.39 | 2.16 | 0.22 | -0.36 | -1.44 | -1.68 | 0.30 | 2.29 | 0.66 | 0.00 | 0.89 | -1.50 | -1.85 |
| MYDGF | Q969H8 | 1.16 | -0.66 | -1.82 | -1.59 | -1.47 | -1.68 | -1.89 | -1.65 | -0.30 | -0.30 | -1.56 | -1.51 | -1.14 | 0.17 | -0.38 | -0.50 | 0.07 | 0.31 | -1.45 | 0.35 | -0.80 | -1.29 | -0.97 | -1.82 | 1.01 | -1.49 | 0.59 | -1.56 | -0.33 | -1.70 | 0.13 |
| MYH14 | Q7Z406 | 1.09 | 0.58 | -1.35 | -1.49 | -1.72 | -1.99 | 0.08 | -1.43 | 0.01 | 0.00 | -1.39 | -1.71 | -1.61 | 0.18 | 0.18 | -0.03 | 1.29 | 0.69 | -1.94 | 1.15 | 0.76 | -2.00 | -2.07 | -2.09 | 0.08 | -1.54 | 0.51 | 0.59 | 0.56 | -1.48 | -1.62 |
| MYH9 | P35579 | 1.81 | -0.35 | -1.79 | -0.09 | -0.35 | -1.46 | -0.20 | 1.07 | 0.85 | 0.25 | -1.96 | -1.49 | -0.50 | 2.17 | 0.83 | 1.24 | 1.52 | 0.87 | 0.36 | 1.73 | 0.59 | 0.32 | 0.16 | -0.46 | 2.17 | 0.79 | 2.04 | 0.93 | 1.46 | -0.40 | -2.19 |
| MYL6 | P60660 | 1.62 | -0.87 | -0.67 | -1.76 | -1.67 | -1.19 | -0.80 | -1.77 | -0.29 | -1.56 | -1.81 | -1.01 | -0.70 | 1.45 | -0.08 | 0.52 | 1.46 | 1.06 | -0.11 | 1.07 | 0.57 | -0.35 | -0.82 | -0.59 | 1.65 | -0.14 | 1.90 | 0.40 | 0.77 | -0.76 | -1.54 |
| MYO1F | O00160 | -0.47 | -1.09 | -1.42 | -1.45 | -1.27 | -1.32 | -1.20 | -1.73 | -0.28 | -1.54 | -2.20 | -1.06 | -1.17 | 0.27 | -2.13 | -0.26 | -2.00 | -1.57 | -1.44 | 0.50 | -1.10 | -1.11 | -1.37 | -1.55 | 1.12 | -0.33 | 1.19 | -2.19 | -0.11 | -1.21 | -0.61 |
| NAGK | Q9UJ70 | 0.37 | -0.25 | -1.65 | -2.01 | -1.49 | -1.24 | -1.43 | -1.52 | -0.65 | -1.18 | -0.59 | -1.73 | -0.74 | 0.31 | -0.20 | -0.09 | 0.27 | -0.01 | -0.38 | 0.35 | -0.47 | -0.58 | -0.30 | -0.66 | 0.71 | -0.39 | 0.50 | -0.10 | -0.01 | -1.71 | -1.56 |
| NAMPT | P43490 | 1.37 | -1.11 | -1.76 | -1.95 | -1.29 | -1.66 | -1.60 | -1.80 | 0.46 | -0.89 | -1.38 | 1.39 | -1.73 | 1.42 | 0.15 | 1.19 | 0.78 | 0.29 | -1.60 | 1.60 | 0.22 | -0.36 | -2.08 | -2.42 | 1.92 | -0.02 | 1.74 | 0.02 | 1.26 | -1.59 | -1.94 |
| NAPRT | Q6XQN6 | 0.79 | -0.93 | -1.58 | -1.79 | -1.52 | -1.40 | -1.88 | -1.48 | -1.47 | -1.77 | -1.98 | -1.84 | -1.69 | 0.43 | 1.28 | 0.29 | 0.91 | 0.49 | -1.65 | 0.47 | -0.20 | 0.72 | -1.39 | -1.69 | 0.55 | 1.06 | 0.37 | -1.00 | 0.66 | -0.68 | -1.94 |
| NAXE | Q8NCW5 | 0.33 | -0.75 | -1.69 | -1.62 | -1.51 | -1.29 | -1.64 | -1.31 | -0.79 | -1.22 | -1.61 | -1.34 | -1.45 | -0.83 | 0.27 | 0.24 | 0.58 | 0.53 | -0.93 | -0.06 | -0.91 | 0.22 | -1.97 | -0.53 | -0.95 | 0.11 | -1.23 | -1.60 | 0.10 | -1.61 | -1.39 |
| NCF2 | P19878 | -0.11 | 0.15 | -1.61 | -1.51 | -0.93 | -1.68 | -1.27 | -1.36 | -1.57 | -1.46 | -1.50 | -1.83 | -1.41 | 0.63 | -1.78 | -0.31 | -1.53 | -1.76 | -1.58 | 0.67 | -1.95 | -1.11 | -1.49 | -1.26 | 1.22 | -1.38 | 1.49 | -1.38 | -0.04 | -1.67 | -1.86 |
| NCSTN | Q92542 | -0.97 | -0.90 | -1.78 | -1.64 | -1.35 | -0.87 | -0.89 | -0.61 | -0.57 | -1.27 | -0.59 | -0.24 | -1.86 | -0.26 | -0.90 | -0.27 | -0.41 | -0.71 | -1.13 | 0.62 | 1.09 | -1.20 | -0.93 | -0.96 | 1.43 | -0.73 | 1.98 | -0.17 | 0.10 | -0.65 | -1.56 |
| NDRG1 | Q92597 | 0.49 | -1.71 | -1.75 | -1.51 | -1.35 | -1.17 | -1.73 | -1.25 | -1.56 | -1.93 | -1.37 | -1.84 | -1.95 | -1.94 | -1.16 | 0.12 | 0.21 | -0.38 | -1.37 | 0.58 | -0.19 | -0.38 | -1.83 | -0.15 | -1.62 | -1.83 | -0.45 | -1.46 | -0.19 | -1.46 | -1.41 |
| NPM1 | P06748 | 0.77 | 0.18 | -0.07 | -1.32 | 0.52 | -0.03 | -0.09 | -1.68 | 0.17 | -0.05 | -0.11 | -0.44 | -0.28 | 1.11 | 0.36 | 1.20 | 1.28 | 0.30 | 0.58 | 1.32 | 1.00 | 0.22 | -0.19 | -0.25 | 1.27 | 0.33 | 1.88 | -0.05 | 0.65 | -0.20 | -1.51 |
| NUDT3 | O95989 | -0.30 | -1.05 | -1.28 | -1.68 | -1.30 | -1.33 | -1.90 | -1.85 | -1.93 | -1.87 | -0.84 | -1.80 | -1.76 | 0.07 | -0.49 | -0.52 | -0.13 | -1.13 | -1.61 | -0.01 | -1.46 | -1.46 | -1.56 | -1.69 | 0.05 | -0.68 | 0.09 | -1.30 | -0.74 | -2.20 | -2.07 |
| NUDT5 | Q9UKK9 | 0.10 | -1.12 | -2.14 | -1.79 | -1.47 | -1.72 | -1.94 | -1.54 | -1.36 | -1.18 | -0.94 | -1.58 | -1.55 | 0.03 | -0.06 | -0.37 | 0.08 | -0.23 | -1.33 | 0.02 | -1.06 | -0.94 | -1.16 | -1.61 | 0.00 | -0.14 | 0.19 | -1.84 | -0.71 | -1.25 | -1.91 |
| OLFM4 | Q6UX06 | 0.44 | 3.25 | -0.66 | -2.04 | -1.28 | 1.21 | 1.31 | -1.89 | 0.43 | 0.37 | -0.88 | -1.21 | 0.18 | 1.27 | 0.11 | 0.93 | 2.24 | 1.93 | -1.47 | 3.38 | 1.46 | 0.97 | 0.94 | 1.58 | 1.16 | 0.56 | 2.05 | 0.19 | 1.67 | 0.73 | -1.65 |
| PA2G4 | Q9UQ80 | 0.93 | -0.26 | -0.61 | -1.45 | -1.62 | -0.51 | -0.59 | -2.08 | -1.02 | -0.60 | -0.95 | -1.63 | -0.50 | 0.95 | 1.29 | 0.57 | 0.74 | 0.13 | -0.28 | 0.78 | -1.56 | 0.65 | -1.85 | -0.60 | 0.42 | 0.99 | 0.57 | -0.03 | 0.38 | 0.08 | 0.60 |
| PADI2 | Q9Y2J8 | 0.28 | -1.47 | -1.12 | -1.78 | -1.28 | -1.63 | -1.64 | -1.48 | -1.74 | -1.57 | -1.58 | -1.84 | -2.38 | -0.02 | -1.49 | -0.25 | -0.06 | 0.06 | -1.54 | 0.79 | -1.69 | -1.28 | -1.15 | -1.60 | 1.05 | -1.56 | 1.02 | -1.86 | 0.12 | -1.76 | 0.19 |
| PADI4 | Q9UM07 | 0.81 | -1.74 | -1.37 | -0.88 | -1.17 | -1.69 | -1.31 | -1.34 | 0.25 | -1.59 | -1.58 | -1.69 | -1.58 | 0.92 | -0.14 | 0.53 | -1.53 | -1.66 | -0.92 | 1.23 | -1.72 | -0.82 | -1.97 | -1.36 | 1.65 | -0.40 | 1.63 | -1.57 | 0.70 | -1.76 | -1.51 |
| PAFAH1B2 | P68402 | 0.79 | -0.43 | -1.44 | -1.27 | -1.21 | -1.04 | -1.00 | -1.83 | -0.75 | -0.92 | -1.26 | -1.23 | -0.90 | 0.95 | 0.87 | 0.61 | 1.22 | 0.58 | -0.29 | 0.64 | 0.40 | 0.56 | -0.85 | -0.95 | 0.68 | 0.67 | 0.92 | -0.19 | 0.21 | -0.55 | -1.70 |
| PARK7 | Q99497 | 1.63 | 0.28 | -1.57 | -0.68 | -0.22 | 0.00 | -0.39 | -0.03 | -0.49 | -0.32 | -0.66 | -0.78 | -0.15 | 0.15 | 1.90 | 1.27 | 1.75 | 1.43 | 0.31 | 1.30 | 0.08 | 1.46 | -0.69 | -0.06 | 0.67 | 1.68 | 0.35 | 0.06 | 1.41 | 0.34 | -0.59 |
| PCBP1 | Q15365 | 1.22 | -0.45 | -0.40 | -0.96 | -0.68 | -0.41 | -0.77 | -1.55 | -1.68 | -0.46 | -0.69 | -0.65 | -0.42 | 0.16 | 0.79 | 0.62 | 1.42 | 0.71 | 0.16 | 1.24 | 0.36 | 0.19 | -0.65 | -0.57 | 1.32 | 0.47 | 1.26 | -0.05 | 0.72 | -1.31 | -1.57 |
| PCMT1 | P22061 | 0.80 | -0.74 | -2.13 | -1.08 | -1.14 | -1.79 | -1.47 | -1.43 | -1.89 | -1.25 | -1.44 | -1.50 | -1.72 | -0.52 | 1.37 | 0.39 | 0.60 | 0.38 | -0.33 | 0.47 | -0.63 | 1.07 | -1.63 | -0.98 | 0.16 | 1.30 | 0.07 | -1.86 | 0.67 | -0.26 | -1.64 |
| PDCD10 | Q9BUL8 | -0.06 | -0.18 | -1.59 | -2.22 | -2.02 | -1.59 | -1.69 | -1.44 | -1.49 | -0.73 | -1.68 | -2.20 | -0.32 | 0.53 | -0.44 | -0.51 | 0.22 | 0.09 | -0.85 | 0.18 | -1.02 | -1.36 | -1.33 | -0.01 | 0.11 | -0.42 | 0.40 | -0.52 | -0.19 | -1.71 | -1.04 |
| PDCD6 | O75340 | -0.27 | -1.27 | -1.29 | -1.48 | -1.44 | -1.53 | -0.65 | -1.26 | -1.02 | -0.71 | -1.34 | -1.56 | -1.34 | -0.72 | -0.22 | -0.39 | -0.23 | -0.92 | -1.71 | 0.48 | 0.39 | -0.58 | -1.75 | -0.51 | 0.33 | -0.37 | 0.43 | -0.65 | -0.46 | -1.93 | -1.75 |
| PDXK | O00764 | 0.61 | -0.44 | -1.34 | -1.80 | -1.14 | -2.03 | -1.49 | 0.14 | -1.60 | -1.44 | -2.26 | -1.23 | -1.72 | 0.32 | -0.17 | 0.42 | 1.02 | 0.45 | -1.87 | 0.84 | -0.44 | -1.27 | -1.42 | -1.77 | 0.94 | -0.23 | 1.22 | -0.01 | 0.57 | -1.41 | -1.19 |
| PEBP1 | P30086 | 1.16 | 0.32 | -2.04 | -1.64 | -0.68 | -0.58 | -0.34 | -1.99 | -1.19 | -0.07 | -1.45 | -0.51 | -0.08 | 0.31 | 1.24 | 0.44 | 1.44 | 0.99 | 0.14 | 0.73 | 0.18 | 0.72 | -0.34 | -0.68 | 0.43 | 0.98 | 0.24 | -0.42 | 0.25 | -0.14 | -0.76 |
| PGAM1 | P18669 | 1.78 | 0.21 | -0.61 | -1.96 | -1.80 | -0.17 | -1.56 | -1.60 | -0.06 | -1.16 | -1.43 | -1.40 | -0.75 | 1.96 | 1.27 | 1.13 | 1.71 | 1.20 | 0.49 | 1.80 | 0.65 | 0.98 | -0.59 | -1.21 | 1.90 | 1.00 | 1.83 | 0.22 | 1.52 | 0.17 | -0.29 |
| PGD | P52209 | 1.73 | 0.31 | 0.33 | 0.03 | 0.25 | 0.27 | 0.16 | -0.17 | 0.50 | 0.23 | -0.21 | 0.08 | 0.27 | 2.49 | 1.45 | 1.22 | 0.96 | 0.71 | 0.01 | 1.81 | 0.64 | 0.89 | 0.18 | 0.13 | 2.35 | 1.42 | 2.33 | 0.69 | 1.56 | 0.24 | -0.07 |
| PGM1 | P36871 | 1.45 | 0.07 | -1.61 | -1.64 | 1.61 | -1.56 | -0.93 | -1.21 | 0.77 | 0.70 | -1.57 | -1.64 | -1.55 | 1.50 | 0.51 | 0.52 | 1.50 | 0.88 | -1.81 | 1.36 | 0.43 | 0.34 | -1.41 | 0.73 | 1.09 | 0.23 | 0.69 | 0.35 | 0.87 | -1.36 | -1.35 |
| PGP | A6NDG6 | -0.45 | -2.10 | -1.85 | -1.83 | -1.26 | -1.71 | -1.44 | -1.84 | -1.40 | -1.45 | -0.81 | -1.84 | -1.33 | 0.48 | -0.05 | -0.40 | -0.95 | -1.29 | -2.21 | -0.55 | -1.33 | -0.36 | -1.79 | -1.73 | -0.01 | -0.15 | 0.00 | -1.74 | -0.30 | -1.22 | -1.36 |

| | | | | | | | | | | | | | | | | | | | | | | | | | | | | | | | | |
|---|---|---|---|---|---|---|---|---|---|---|---|---|---|---|---|---|---|---|---|---|---|---|---|---|---|---|---|---|---|---|---|---|
| PPA1 | Q15181 | 1.25 | -0.55 | -1.80 | -2.04 | -1.39 | -1.20 | -1.01 | -1.59 | -1.91 | -1.80 | -1.16 | -0.50 | -1.72 | 0.13 | 0.82 | 0.17 | 1.51 | 0.71 | -0.24 | 0.83 | 0.17 | 0.20 | -1.66 | -1.85 | 0.25 | 0.46 | 0.56 | 0.11 | 0.47 | -1.44 | -1.64 |
| PPIA | P62937 | 2.76 | 1.62 | 1.63 | 0.56 | 1.29 | 1.91 | 0.69 | 0.76 | 1.11 | 0.93 | 1.17 | 1.42 | 1.20 | 2.66 | 2.24 | 1.81 | 2.84 | 2.25 | 1.54 | 2.45 | 2.05 | 1.98 | 0.96 | 0.95 | 2.72 | 2.09 | 2.82 | 1.46 | 2.03 | 1.07 | 0.73 |
| PPIB | P23284 | 1.91 | 0.47 | 0.03 | 0.22 | -0.20 | -0.01 | 0.19 | -0.75 | 0.41 | 0.14 | -0.31 | -0.12 | -0.13 | 0.99 | 0.52 | 1.01 | 1.19 | 0.87 | -0.36 | 1.38 | 0.73 | -0.16 | -0.41 | -0.11 | 1.56 | 0.19 | 1.57 | 0.14 | 0.87 | -0.12 | -1.65 |
| PPP1CB | P62140 | 0.21 | -0.64 | 0.00 | -1.94 | -0.66 | -0.66 | -0.91 | -1.13 | -0.86 | -0.71 | -1.42 | -1.82 | -0.74 | -0.07 | -0.49 | -0.38 | 0.08 | -0.68 | -0.99 | 0.78 | -0.34 | -0.96 | -1.37 | -0.56 | 0.73 | -0.48 | 0.56 | -0.49 | -0.31 | -1.70 | -1.26 |
| PREP | P48147 | 0.36 | -1.62 | -1.61 | -1.60 | -0.91 | -1.23 | -1.81 | -1.03 | -0.55 | -1.08 | -1.82 | -1.41 | -1.14 | 0.33 | 0.01 | -0.24 | 0.40 | -0.12 | -2.17 | 0.34 | -0.13 | -0.61 | -1.57 | -1.61 | 0.11 | -0.18 | 0.40 | -0.33 | 0.16 | -1.06 | -1.36 |
| PRG2 | P13727 | 0.26 | -1.83 | -1.66 | -2.00 | -1.54 | -1.26 | -2.18 | 0.06 | 1.63 | -1.89 | -1.28 | -1.09 | -2.22 | 0.88 | 0.38 | 0.15 | -1.56 | -0.26 | 0.39 | 0.10 | 0.24 | 0.65 | -1.35 | -1.51 | 2.56 | 0.39 | 1.69 | 0.12 | 0.75 | -1.62 | -1.80 |
| PRKAR1A | P10644 | -0.32 | -1.71 | -2.18 | -1.54 | -1.44 | -0.95 | -1.35 | -2.13 | -1.66 | -2.22 | -0.69 | -1.47 | -2.19 | -0.38 | 0.04 | -0.54 | -0.49 | -1.52 | -1.29 | 0.41 | -1.30 | -0.79 | -1.74 | -1.05 | 0.83 | -0.29 | 0.96 | -1.72 | -0.14 | -1.24 | -1.35 |
| PRTN3 | P24158 | 2.34 | -0.56 | -0.88 | -1.39 | -1.80 | -0.50 | -0.51 | -0.89 | 1.86 | -0.49 | -0.04 | 0.75 | 0.49 | 4.44 | 0.85 | 1.24 | -0.24 | 0.02 | -0.29 | 2.53 | 2.17 | 0.35 | 0.04 | -0.27 | 4.40 | 0.97 | 4.57 | 1.21 | 1.80 | 0.36 | -0.12 |
| PSMA1 | P25786 | 1.38 | -0.32 | -0.57 | -0.87 | -0.44 | -0.56 | -1.90 | -1.53 | -0.47 | -0.71 | -0.66 | -0.85 | -0.71 | 0.60 | 1.41 | 1.19 | 1.06 | 0.67 | -0.10 | 1.25 | 0.68 | 0.94 | -0.36 | -0.33 | 0.91 | 1.28 | 1.06 | 0.23 | 1.22 | 0.13 | -1.05 |
| PSMA2 | P25787 | 0.96 | -0.45 | -1.61 | -1.65 | -1.37 | -1.71 | -1.77 | -1.19 | -0.32 | -1.18 | -1.24 | -1.88 | -1.31 | 0.43 | 0.95 | 0.64 | 0.57 | -0.07 | -0.44 | 0.92 | -0.07 | 0.30 | -1.68 | -1.37 | 0.54 | 0.85 | 0.61 | -0.49 | 0.63 | -0.10 | -1.27 |
| PSMA3 | P25788 | 0.61 | -1.33 | -1.71 | -1.72 | -0.36 | -0.08 | -0.18 | -1.17 | -0.77 | -0.03 | -0.38 | -0.20 | -0.17 | 0.50 | 0.72 | 0.47 | 0.27 | -0.15 | -0.92 | 0.71 | 0.02 | 0.37 | -1.55 | -1.23 | 0.53 | 0.66 | 0.59 | -0.29 | 0.33 | -0.58 | -0.86 |
| PSMA6 | P60900 | 0.94 | -0.53 | -1.28 | -1.19 | -0.80 | -1.00 | -0.59 | -1.71 | -0.49 | -0.66 | -1.83 | -0.47 | -0.52 | 0.94 | 1.02 | 0.62 | 0.54 | 0.27 | -1.06 | 0.94 | 0.20 | 0.25 | -1.24 | -0.15 | 0.89 | 0.86 | 1.23 | 0.17 | 0.42 | -0.38 | -1.60 |
| PSMB2 | P49721 | 1.00 | -1.23 | -1.25 | -1.59 | -1.87 | -0.87 | -1.36 | -1.75 | -1.66 | -1.51 | -1.41 | -1.71 | -1.28 | -0.30 | 0.77 | 0.58 | 0.48 | 0.17 | 0.06 | 0.75 | -0.22 | 0.22 | -0.64 | -0.56 | -0.01 | 0.55 | 0.28 | -0.42 | 0.51 | -0.45 | -1.91 |
| PSMB5 | P28074 | -0.33 | -1.58 | -1.29 | -1.38 | -0.79 | -1.30 | -0.97 | -1.50 | -1.54 | -1.55 | -1.45 | -1.53 | -1.72 | -0.03 | 0.72 | 0.22 | -0.63 | -0.83 | -1.37 | -0.14 | -0.49 | -0.09 | -1.36 | -1.36 | -0.65 | 0.51 | -0.30 | -1.27 | -0.28 | -0.56 | -1.71 |
| PSMB7 | Q99436 | 0.49 | -1.38 | -1.47 | -1.67 | -1.38 | -1.50 | -0.98 | -1.51 | -1.71 | -0.62 | -1.83 | -1.78 | -1.31 | -0.17 | 0.71 | 0.35 | -0.02 | -0.37 | -1.53 | 0.44 | -0.80 | 0.13 | -1.75 | -0.87 | 0.37 | 0.48 | 0.63 | -0.75 | 0.19 | -0.58 | -1.67 |
| PSME1 | Q06323 | 1.33 | -1.45 | -1.72 | -1.21 | -1.72 | -1.13 | -1.25 | -0.28 | -0.66 | -1.37 | 0.56 | -1.19 | -1.48 | 1.13 | 1.27 | 0.53 | 1.19 | 0.74 | 0.08 | 1.02 | 0.22 | 0.75 | -1.53 | -1.38 | 0.72 | 0.93 | 0.89 | 0.03 | 0.77 | -0.82 | -1.72 |
| PSME2 | Q9UL46 | 0.73 | 0.23 | -1.25 | -2.01 | -1.45 | -1.27 | -2.23 | -1.58 | -1.73 | -0.80 | -1.62 | -1.70 | -1.16 | 0.23 | 0.72 | 0.11 | 0.55 | 0.02 | -0.14 | 0.43 | -0.78 | 0.21 | -1.15 | -0.88 | -0.32 | 0.40 | -0.31 | -1.10 | 0.31 | -1.48 | -1.57 |
| PTBP1 | P26599 | 0.83 | -0.28 | -1.47 | 0.85 | -1.75 | -1.51 | -2.28 | -1.58 | -1.19 | -1.79 | -1.66 | -1.63 | -1.37 | 0.41 | 0.09 | 0.20 | 0.78 | 0.08 | -0.13 | 0.34 | -0.20 | -0.27 | -1.77 | -1.12 | 0.14 | -0.16 | 0.20 | -0.91 | -0.19 | -1.81 | 0.77 |
| PTGES3 | Q15185 | 1.03 | 0.05 | 0.18 | -0.56 | 0.16 | 0.19 | 0.13 | -0.53 | -0.08 | 0.11 | -0.25 | -0.37 | 0.12 | 1.22 | 0.97 | 0.60 | 1.26 | 0.78 | 0.13 | 1.44 | 0.65 | 0.57 | -0.20 | 0.32 | 1.41 | 0.71 | 1.50 | 0.65 | 0.54 | 0.15 | -1.03 |
| PTMA | P06454 | 1.48 | 0.39 | -0.66 | -0.47 | -0.56 | -0.37 | -0.56 | -1.50 | -0.82 | -0.56 | -0.82 | -1.69 | 0.19 | 0.49 | 0.30 | 1.68 | 2.04 | 1.24 | 0.93 | 0.24 | -0.42 | 0.15 | -0.42 | -0.60 | 0.30 | 0.04 | 0.54 | -0.25 | 0.65 | -1.77 | -1.21 |
| PTPRJ | Q12913 | -0.19 | 0.44 | -1.52 | -1.57 | -1.55 | -1.60 | -1.45 | -0.35 | 0.22 | 0.57 | -0.97 | -1.71 | -1.82 | 0.06 | -1.38 | 0.01 | -0.33 | -0.29 | -0.16 | 0.54 | -0.26 | -0.39 | -1.89 | 0.11 | 0.86 | -0.71 | 1.08 | -1.37 | 0.38 | -2.02 | -0.60 |
| RAB11B | Q15907 | 1.31 | 0.27 | 0.17 | -0.43 | 0.14 | 0.25 | 0.03 | -0.20 | 0.10 | 0.33 | -0.38 | -0.16 | 0.52 | 1.44 | 0.74 | 0.78 | 1.17 | 0.79 | -0.19 | 2.03 | 1.10 | 0.23 | 0.29 | 0.42 | 1.64 | 0.69 | 1.57 | 1.16 | 1.29 | 0.04 | -0.69 |
| RAB21 | Q9UL25 | 0.07 | -1.31 | -1.46 | -1.87 | -1.32 | -1.80 | -1.65 | -1.85 | -0.20 | 0.16 | -1.31 | -1.33 | -1.47 | 0.99 | -0.12 | -0.03 | -0.24 | -0.97 | -1.38 | 0.46 | -0.40 | -1.01 | -0.68 | -0.03 | 0.88 | -0.37 | 1.16 | -0.27 | -0.16 | -1.16 | -1.49 |
| RAB27A | P51159 | 0.17 | -1.57 | -1.51 | -1.81 | -1.76 | -1.58 | -1.37 | -0.59 | -2.03 | -0.10 | -1.32 | -1.25 | -0.96 | 0.35 | -0.42 | -0.72 | 0.15 | 0.18 | -1.49 | 1.36 | -0.06 | -1.29 | -1.64 | -1.83 | 1.41 | -0.60 | 1.22 | 0.96 | 0.12 | -1.20 | -0.95 |
| RAB33B | Q9H082 | -1.51 | 0.03 | -0.86 | -0.80 | -1.29 | -1.39 | -1.23 | -1.45 | -1.62 | -1.56 | -1.80 | -1.84 | -1.93 | -0.10 | 0.78 | 0.34 | 0.82 | -1.96 | -0.70 | 1.41 | 0.69 | -0.27 | -1.53 | -1.24 | 1.07 | 0.31 | 1.25 | 1.13 | 0.55 | -1.28 | -1.56 |
| RAB35 | Q15286 | 0.68 | 0.52 | 0.49 | -1.73 | -1.11 | 0.59 | -1.47 | -1.85 | -1.37 | -1.89 | -1.50 | -1.69 | -1.46 | 0.24 | 0.80 | 0.18 | 0.83 | -2.01 | -1.77 | 1.59 | 0.82 | -1.75 | 0.25 | -1.34 | 0.91 | 0.69 | 1.45 | 0.98 | 0.57 | -1.97 | -1.43 |
| RAP1B | P61224 | 1.06 | 0.89 | 0.33 | 0.28 | -0.03 | 0.20 | 0.17 | -1.73 | 0.32 | 0.59 | -0.21 | -0.40 | 0.58 | 1.27 | 1.15 | 0.94 | 1.05 | 0.81 | 0.57 | 1.83 | 1.01 | 0.64 | -0.18 | 0.41 | 2.11 | 1.04 | 2.21 | 0.83 | 1.26 | 0.40 | -0.24 |
| RAP2B | P61225 | -1.71 | -1.01 | -1.42 | -2.02 | -2.32 | -1.61 | -1.67 | -1.15 | -1.75 | -0.84 | -1.53 | -2.13 | -0.99 | -0.41 | 0.33 | -0.26 | -0.39 | -1.10 | -1.41 | 0.48 | -0.29 | 0.63 | -1.40 | -1.10 | 0.63 | 0.11 | 0.72 | -0.56 | -0.01 | -1.10 | -1.29 |
| RCC2 | Q9P258 | -0.12 | -1.90 | -1.63 | -1.76 | -1.85 | -0.72 | -1.83 | -1.16 | -2.14 | -1.55 | 0.14 | -1.69 | -1.74 | 0.30 | -1.71 | 0.07 | -0.12 | -1.02 | 0.11 | 0.03 | -1.25 | -0.15 | -1.71 | -1.30 | 0.42 | 0.26 | 0.25 | -1.31 | -0.77 | -1.39 | 0.17 |
| REG3A | Q06141 | 1.74 | 1.18 | -1.77 | -1.62 | -0.27 | -0.10 | 1.79 | -1.72 | 0.89 | -0.26 | 1.53 | -1.80 | -0.51 | 0.10 | 0.48 | -1.21 | 1.80 | 1.35 | -1.79 | 0.92 | 0.15 | -1.75 | -2.10 | 0.06 | 0.20 | 0.15 | -1.64 | 0.45 | -1.52 | -0.21 | -1.29 |
| RHOA | P61586 | 1.50 | 0.48 | -0.12 | -0.25 | -0.05 | -0.01 | -0.14 | -0.50 | 0.39 | 0.05 | -0.46 | -0.50 | 0.13 | 1.43 | 0.79 | 0.70 | 1.34 | 0.89 | 0.27 | 1.91 | 0.84 | 0.37 | -0.21 | 0.30 | 2.09 | 0.62 | 2.39 | 0.91 | 1.12 | -0.16 | -0.39 |
| RNASE2 | P10153 | 0.72 | 0.70 | -1.31 | -0.62 | -1.67 | -1.77 | -1.71 | -1.55 | 1.16 | -1.52 | -0.40 | -0.35 | -1.84 | 1.97 | -2.06 | 0.29 | -1.68 | -1.49 | -0.42 | 0.20 | 0.34 | -0.69 | -1.20 | -1.17 | 2.33 | -0.23 | 2.65 | -0.46 | 0.25 | -1.46 | -1.94 |
| RNPEP | Q9H4A4 | 1.09 | -1.32 | -1.64 | -1.85 | -1.21 | -1.57 | -1.78 | -1.29 | -1.74 | -1.35 | -1.48 | -1.51 | -1.81 | 0.27 | 0.25 | -0.09 | 0.92 | 0.53 | -0.84 | 0.39 | 0.04 | -0.17 | -1.39 | 0.40 | 0.56 | -0.32 | 0.64 | -0.13 | 0.18 | -1.09 | -1.54 |
| RPIA | P49247 | -0.17 | -2.14 | -1.96 | -2.03 | -1.11 | -1.39 | -1.51 | -1.19 | -0.76 | -1.16 | 0.05 | 0.14 | -1.95 | 0.54 | 1.14 | 0.51 | 0.01 | -0.45 | -1.31 | 0.20 | -0.22 | 0.61 | -1.88 | 0.84 | 0.39 | 0.87 | 0.71 | -1.43 | 0.41 | -0.14 | -1.82 |
| RPL28 | P46779 | 0.33 | -2.01 | -1.85 | -1.94 | -1.21 | -1.25 | -0.92 | -1.49 | -1.21 | -2.04 | -1.51 | -2.37 | -1.30 | -2.47 | -0.16 | 0.37 | 0.46 | -0.24 | 0.38 | 0.87 | -0.41 | -0.44 | -1.26 | -1.79 | 0.26 | -0.74 | 0.79 | 0.10 | 0.04 | -1.50 | -1.81 |
| RPL30 | P62888 | 0.74 | -1.42 | -0.64 | -1.66 | -1.79 | -0.68 | -1.06 | -1.19 | -0.65 | -0.68 | -1.28 | -1.63 | -0.83 | 0.06 | 0.06 | 0.19 | 0.40 | -0.25 | -0.34 | 0.85 | -0.20 | -0.83 | -1.57 | -0.76 | 0.02 | -0.89 | 0.60 | -0.28 | -0.29 | -1.30 | -0.10 |
| RPS12 | P25398 | 1.22 | -1.86 | -1.28 | -1.87 | -1.57 | -1.43 | -1.61 | -1.43 | -0.52 | -1.14 | -1.44 | -1.66 | -0.41 | 0.13 | 0.11 | 0.60 | 0.88 | 0.31 | 0.24 | 1.01 | 0.06 | -0.02 | -1.41 | -1.73 | 0.45 | -0.12 | 0.95 | -0.55 | 0.56 | -1.59 | -2.04 |
| RPS21 | P63220 | 1.15 | -1.31 | -1.73 | -1.09 | -1.76 | -1.56 | -1.04 | -1.58 | -0.86 | -1.58 | -0.86 | -1.30 | -1.48 | -0.26 | 0.08 | -0.21 | 1.08 | 0.44 | 0.00 | -0.09 | -0.04 | -0.12 | -1.04 | -0.85 | 0.13 | -0.36 | 0.57 | -0.57 | -0.31 | -1.25 | -1.32 |
| S100A11 | P31949 | 1.10 | -0.51 | -1.26 | -1.90 | -1.19 | -0.77 | -0.77 | -2.22 | 0.18 | -1.17 | -2.09 | -1.70 | 0.09 | 1.30 | -0.35 | 0.65 | 1.19 | 0.79 | -0.76 | 1.49 | 1.16 | -0.71 | -0.55 | -0.15 | 1.47 | -0.74 | 1.66 | 0.18 | 0.78 | -0.61 | -1.30 |
| S100A6 | P06703 | 1.20 | 0.32 | -0.43 | -1.02 | -1.34 | 1.11 | -0.12 | -0.83 | -0.19 | 0.24 | -0.84 | -0.78 | 0.56 | 1.00 | 0.64 | 0.50 | 2.03 | 1.46 | -1.54 | 1.41 | 1.97 | 0.60 | 0.30 | 0.30 | 1.79 | 0.80 | 1.67 | 1.02 | 0.91 | 0.36 | -1.16 |
| S100A8 | P05109 | 3.33 | 0.40 | -0.19 | -0.89 | -0.09 | -0.20 | 0.20 | 1.09 | 3.03 | 0.19 | 0.49 | 1.41 | 2.07 | 4.29 | 1.19 | 2.92 | 0.26 | 0.57 | -0.07 | 3.77 | 3.15 | 0.90 | 0.43 | 0.36 | 4.58 | 1.30 | 5.19 | 1.84 | 2.97 | 0.61 | 0.30 |
| S100A9 | P06702 | 2.93 | 0.30 | -0.12 | -0.87 | -0.19 | -0.28 | 0.08 | -0.85 | 2.85 | 0.27 | 0.58 | 1.61 | 1.44 | 4.19 | 1.04 | 2.94 | 0.37 | 0.62 | 0.04 | 3.43 | 3.20 | 0.81 | 0.62 | 0.13 | 4.44 | 1.22 | 5.10 | 1.76 | 2.77 | 0.68 | 0.29 |
| SDCBP | O00560 | -0.20 | 0.78 | -1.62 | 0.05 | -1.43 | -1.08 | 0.06 | -1.03 | -1.68 | 0.88 | -1.49 | -1.72 | 0.70 | 0.36 | -0.44 | 0.00 | 0.45 | 0.51 | -0.66 | 1.41 | 0.87 | -1.46 | -1.53 | 0.60 | 0.58 | 0.05 | 0.82 | 0.23 | 0.67 | -2.14 | -0.76 |
| SERPINA2 | P20848 | 0.13 | -0.40 | -1.65 | -0.22 | -1.18 | -1.67 | -1.17 | -2.08 | 0.06 | -1.64 | -0.60 | -0.61 | -2.09 | 1.61 | -0.97 | -0.49 | -1.37 | -1.31 | -0.66 | 0.79 | -0.25 | -1.82 | 0.06 | -1.83 | 1.63 | -1.87 | 0.91 | -1.69 | 0.20 | -1.21 | 0.68 |
| SERPINB1 | P30740 | 2.19 | 0.18 | -2.00 | -0.15 | 1.41 | -0.84 | 0.77 | -1.36 | 1.33 | 0.52 | 0.56 | -0.28 | -0.71 | 2.83 | 0.89 | 1.33 | 1.74 | 1.00 | -0.21 | 2.42 | 1.50 | 0.34 | 0.19 | 0.04 | 3.50 | 0.77 | 3.65 | 1.26 | 1.84 | 0.17 | -1.19 |
| SERPINB6 | P35237 | 1.66 | -0.34 | 0.15 | -1.71 | -0.21 | 0.24 | -0.65 | -0.25 | 0.51 | 0.04 | 0.16 | 0.15 | -0.92 | 1.83 | 0.35 | 0.28 | 1.70 | 0.81 | -1.40 | 1.68 | 1.45 | -0.15 | 0.10 | 0.46 | 1.66 | 0.01 | 1.95 | 0.90 | 1.01 | -1.81 | -1.91 |
| SERPINB9 | P50453 | -0.46 | -1.70 | -1.11 | -1.54 | -0.47 | -1.27 | -1.61 | -1.74 | -0.49 | -1.54 | -1.42 | -1.04 | -1.01 | 0.29 | -0.66 | -0.31 | -0.49 | -1.75 | -0.32 | -0.07 | 0.50 | -0.44 | -0.88 | -1.83 | -0.41 | -0.83 | 0.53 | -1.49 | -0.26 | -1.38 | -1.80 |
| SERPIND1 | P05546 | 2.20 | 0.08 | -1.74 | 1.58 | -1.10 | -1.09 | -1.93 | 1.61 | 1.44 | -1.39 | -1.75 | -2.15 | -1.76 | 1.71 | 2.20 | 2.25 | 0.71 | 0.45 | 1.97 | 1.54 | 0.13 | 2.30 | -0.22 | -1.07 | 1.12 | 2.09 | 0.59 | -0.39 | 2.26 | 0.42 | 1.83 |
| SFN | P31947 | 1.31 | -1.38 | -2.00 | -1.02 | -1.79 | -1.43 | -1.30 | -1.72 | 0.43 | -1.83 | -1.42 | -1.44 | -1.64 | 1.08 | 1.08 | 0.95 | 2.08 | 1.13 | -0.10 | 1.38 | 0.77 | -1.33 | 0.93 | -0.25 | 1.22 | 0.94 | 0.79 | 0.72 | 0.85 | -0.16 | -1.51 |
| SH3BGRL | O75368 | 0.37 | -0.35 | -1.44 | -0.02 | -1.37 | -1.26 | -1.38 | -1.86 | -1.70 | -1.47 | -1.90 | -1.34 | -1.35 | 0.78 | 0.49 | -0.04 | 0.53 | 0.16 | -1.65 | -0.05 | 1.08 | 0.23 | -1.34 | -1.40 | 0.80 | 0.37 | 0.61 | -2.11 | -0.14 | -0.50 | -1.67 |
| SH3BGRL3 | Q9H299 | 1.86 | 0.49 | -1.55 | -1.52 | -1.22 | -1.68 | -1.66 | -1.54 | -2.06 | -1.57 | -1.93 | -1.86 | -1.87 | 0.42 | 1.30 | 0.83 | 1.62 | 1.22 | 0.73 | 1.29 | -1.40 | 0.75 | -0.67 | -0.48 | 0.12 | 1.14 | -0.10 | -0.01 | 1.33 | 0.13 | -0.24 |
| SNRNP70 | P08621 | -0.27 | -1.07 | -1.64 | -1.26 | -1.46 | -1.43 | -1.25 | -2.11 | -1.55 | -1.99 | -1.52 | -2.00 | -1.80 | -0.10 | -1.44 | -0.48 | -0.62 | -1.91 | -0.85 | -0.63 | -1.22 | -1.30 | -2.11 | -1.46 | 0.16 | -2.32 | 0.12 | -1.62 | -1.29 | -1.49 | -1.95 |
| SNRPD1 | P62314 | 0.43 | -1.67 | -1.27 | -1.14 | -1.35 | -1.65 | -1.83 | -1.41 | -1.40 | -1.43 | -1.18 | -1.04 | -1.04 | 0.68 | -1.93 | 0.35 | 0.06 | -1.48 | -1.87 | 0.35 | 0.01 | -1.09 | 0.69 | -1.32 | 0.52 | -1.19 | 0.51 | -1.69 | 0.37 | -1.51 | -1.42 |
| SRI | P30626 | 1.23 | -0.56 | -1.69 | -1.30 | -1.65 | -1.17 | -1.02 | -1.63 | -1.53 | -1.71 | -1.55 | -1.68 | -1.84 | -0.87 | 1.00 | 0.29 | 1.27 | 0.48 | -1.93 | 0.87 | 1.11 | 0.65 | -2.03 | -0.37 | 0.70 | 0.97 | 0.40 | 0.41 | 0.41 | -0.07 | -1.35 |
| SSRP1 | Q08945 | -0.65 | -1.80 | -1.86 | -1.77 | -1.64 | -1.27 | -1.21 | -1.81 | -1.12 | -1.38 | -1.51 | -1.61 | -1.53 | 0.11 | -2.06 | -0.14 | -1.06 | -1.87 | -1.31 | -0.75 | -1.42 | -1.69 | -1.88 | -1.25 | -0.39 | -1.52 | -0.48 | -1.54 | -1.95 | -1.57 | -1.55 |
| STIP1 | P31948 | 0.52 | -1.96 | -1.50 | -1.94 | -1.63 | -1.65 | -1.61 | -0.53 | -0.10 | -1.52 | -1.20 | -1.67 | -0.63 | 0.26 | 1.08 | 0.31 | 0.60 | 0.31 | -0.07 | 0.13 | -0.39 | 0.59 | -1.72 | -0.63 | 0.28 | 0.75 | 0.30 | -0.05 | 0.23 | -1.05 | -1.43 |
| STOM | P27105 | -0.24 | 0.18 | 0.18 | -0.67 | 1.11 | -2.32 | -0.09 | -1.59 | 0.11 | 0.22 | 1.01 | 1.11 | 1.66 | 0.34 | 1.81 | 1.32 | 0.92 | 0.97 | -1.66 | 2.07 | 0.91 | 1.30 | -0.66 | 0.39 | 2.46 | 1.69 | 3.48 | 0.25 | 1.50 | 0.60 | -1.71 |
| SUMO2 | P61956 | 0.81 | -1.14 | -1.20 | -1.57 | -1.72 | -1.69 | -1.55 | -1.52 | -1.54 | -1.64 | -1.40 | -1.17 | -1.18 | 0.62 | 0.47 | 0.49 | 1.09 | 0.53 | 0.11 | 0.34 | -1.90 | 0.02 | -1.48 | -2.07 | 0.20 | 0.16 | 0.22 | -1.47 | 0.41 | -2.06 | -1.11 |

| | | | | | | | | | | | | | | | | | | | | | | | | | | | | | | | | |
|---|---|---|---|---|---|---|---|---|---|---|---|---|---|---|---|---|---|---|---|---|---|---|---|---|---|---|---|---|---|---|---|---|
| SYNGR2 | O43760 | -0.10 | -1.67 | -1.55 | -1.60 | -1.46 | -1.70 | -1.56 | -1.37 | -1.71 | -0.98 | -1.44 | 0.36 | -1.61 | -0.46 | -0.51 | 0.12 | -0.17 | -2.14 | -1.61 | 1.26 | 0.20 | -1.53 | -1.40 | -1.69 | 0.84 | -1.88 | 1.19 | 0.16 | -0.05 | -1.32 | -1.00 |
| TACSTD2 | P09758 | -1.69 | -1.20 | -1.48 | -1.38 | -1.35 | -1.44 | -1.24 | -1.55 | -1.83 | -1.45 | -1.77 | -1.76 | -1.82 | -1.91 | -1.48 | -0.29 | 0.03 | -0.18 | -1.76 | 0.35 | 0.33 | -1.49 | -1.46 | -1.40 | -0.84 | -1.30 | -0.25 | -1.14 | -0.18 | -2.01 | -1.42 |
| TAGLN2 | P37802 | 1.50 | -0.21 | -2.07 | -2.15 | -1.23 | -1.99 | -0.83 | -2.24 | -1.38 | -1.02 | -1.20 | -1.64 | -0.44 | -0.80 | 1.08 | 0.51 | 2.22 | 1.50 | 0.40 | 1.11 | 0.42 | 0.71 | -1.26 | -0.14 | 0.00 | 0.89 | 0.24 | 0.46 | 0.99 | -0.49 | -1.29 |
| TALDO1 | P37837 | 1.89 | 0.30 | -0.06 | -0.40 | -0.25 | -0.20 | -0.24 | -0.77 | 0.50 | -0.24 | -0.20 | 0.48 | -0.08 | 1.95 | 1.60 | 1.42 | 1.48 | 0.88 | 0.68 | 1.80 | 0.97 | 1.18 | -0.14 | -0.43 | 2.03 | 1.47 | 2.19 | 0.28 | 1.68 | 0.28 | -0.44 |
| TCN1 | P20061 | 0.82 | 1.15 | -1.38 | -1.36 | -1.70 | -1.40 | 0.40 | -1.89 | -0.46 | 0.47 | -1.38 | -1.63 | 1.23 | 1.20 | 1.29 | -1.00 | 0.18 | 1.54 | -1.87 | 1.35 | 0.57 | -1.42 | -1.42 | 1.08 | 1.24 | 0.12 | 1.12 | 0.69 | 1.05 | -0.67 | -1.47 |
| TKT | P29401 | 2.24 | 0.53 | 0.45 | -0.10 | 0.33 | 0.42 | 0.31 | -0.14 | 0.34 | 0.38 | 0.04 | 0.19 | 0.50 | 2.65 | 1.18 | 1.46 | 1.78 | 1.18 | 0.58 | 2.27 | 1.00 | 0.84 | 0.12 | 0.27 | 3.02 | 1.05 | 2.97 | 0.80 | 1.94 | 0.38 | -0.35 |
| TLN1 | Q9Y490 | 0.77 | -2.08 | -1.37 | -1.36 | -1.57 | -1.14 | -1.00 | -1.58 | 0.19 | -1.22 | -1.85 | -1.60 | -1.93 | 0.60 | 0.65 | 0.31 | 0.60 | -0.09 | -0.43 | 0.57 | -1.54 | -0.45 | -0.64 | -1.65 | 0.48 | 0.89 | 0.31 | -1.28 | 0.45 | -1.64 | -2.15 |
| TM9SF3 | Q9HD45 | 0.10 | 0.49 | 0.57 | -1.72 | 0.34 | -0.95 | -1.18 | -1.88 | -1.70 | -1.84 | -1.64 | -1.74 | -1.75 | -1.05 | -0.35 | -0.33 | -0.07 | -0.52 | -1.34 | 1.45 | -0.02 | -1.18 | -2.00 | -1.95 | 0.43 | -0.96 | 1.04 | -0.15 | 0.11 | -1.37 | -1.27 |
| TMEM30A | Q9NV96 | -1.28 | -1.05 | -0.54 | -1.65 | -0.51 | -0.69 | -1.28 | -0.98 | -0.64 | -0.36 | -1.51 | -1.17 | -0.47 | -0.13 | -0.66 | -0.09 | -0.10 | 0.21 | -0.98 | 0.95 | 0.82 | -1.16 | -0.38 | -1.61 | 1.72 | -0.69 | 2.08 | -0.12 | 0.41 | -1.14 | -1.07 |
| TP53I3 | Q53FA7 | 0.40 | -1.74 | -1.34 | -1.18 | -1.61 | -0.84 | -1.67 | -0.53 | -1.80 | -1.52 | -2.07 | -1.11 | -2.05 | 0.26 | 0.04 | -0.06 | 0.72 | 0.17 | -0.31 | 0.14 | -0.96 | -0.13 | -1.98 | -1.56 | 0.32 | 0.05 | -0.24 | -0.40 | 0.14 | -1.84 | -2.10 |
| TPP1 | O14773 | 0.50 | -0.17 | -1.69 | -1.90 | -1.77 | -1.35 | -1.93 | -1.61 | -1.54 | -0.81 | -1.82 | -0.99 | -1.21 | 0.07 | -0.34 | 0.21 | 0.29 | 0.53 | -1.87 | 0.03 | -0.36 | 0.26 | -1.66 | -1.69 | 0.51 | -1.33 | 0.69 | -0.87 | 0.30 | -0.45 | -1.83 |
| TSPAN3 | O60637 | -0.80 | -1.23 | -1.44 | -1.52 | -1.06 | -0.41 | -1.71 | -1.88 | -1.74 | -1.63 | -1.99 | -1.82 | -1.54 | -0.98 | -1.29 | -0.40 | 0.42 | 0.32 | -1.74 | 1.34 | 0.96 | -1.59 | -1.47 | 0.38 | 0.38 | -1.51 | 1.61 | 0.81 | 0.97 | 0.21 | -1.60 |
| TTN | Q8WZ42 | 0.48 | -0.54 | -1.66 | 0.46 | 0.45 | -1.22 | -1.60 | 0.29 | 0.07 | -1.57 | -0.03 | 0.06 | 0.63 | 0.85 | 0.54 | 1.12 | -0.25 | -1.41 | 0.98 | 1.00 | 0.11 | 1.09 | -1.53 | -1.55 | 0.31 | 0.50 | -0.26 | 0.95 | 1.12 | 0.33 | 1.00 |
| TUT4 | Q5TAX3 | 1.53 | 1.57 | -1.55 | -0.31 | -1.76 | -1.79 | 0.87 | 0.05 | 0.09 | 0.48 | -1.67 | -1.08 | 0.95 | 1.09 | 2.79 | 2.19 | 1.35 | 2.08 | 0.11 | 1.49 | 2.20 | 1.66 | -0.54 | -0.79 | -0.73 | 0.52 | -0.48 | 0.21 | 2.58 | 1.86 | 0.02 |
| TXN | P10599 | 1.78 | 0.54 | 0.23 | -0.07 | -0.10 | -0.07 | 0.24 | -0.34 | 0.14 | 0.04 | 0.11 | 0.11 | 0.03 | 1.49 | 1.67 | 1.19 | 2.07 | 1.41 | -0.37 | 1.59 | 0.79 | 1.20 | -0.16 | 0.35 | 1.38 | 1.30 | 1.60 | 1.04 | 1.28 | 0.52 | -0.22 |
| TXNDC17 | Q9BRA2 | 1.30 | 0.06 | -1.59 | -1.56 | -1.42 | -1.74 | -1.76 | -1.58 | -0.49 | 0.11 | -1.26 | -1.35 | -1.12 | 0.48 | 0.87 | -0.02 | 1.29 | 0.90 | -1.58 | 0.77 | 0.70 | 0.41 | -1.29 | -1.51 | 0.38 | 0.50 | 0.23 | 0.07 | 0.34 | -0.38 | -1.04 |
| TXNL1 | O43396 | 0.93 | -0.91 | -1.48 | -1.82 | -2.08 | -1.34 | -1.24 | -0.50 | -0.73 | -1.44 | -2.28 | -1.26 | -1.15 | 0.34 | 1.26 | 0.50 | 0.85 | 0.32 | -0.51 | 0.25 | 0.04 | 0.84 | -0.57 | -1.47 | 0.43 | 1.00 | 0.48 | -0.26 | 0.52 | -0.14 | -1.44 |
| TXNRD1 | Q16881 | 0.32 | -0.51 | -1.17 | -1.32 | 0.00 | -1.19 | -1.55 | -1.41 | -0.02 | -0.56 | -1.53 | -0.58 | -1.97 | 1.14 | 0.31 | 0.32 | 0.20 | -0.69 | -0.74 | 0.43 | 0.11 | -0.21 | -0.90 | -1.17 | 0.93 | 0.14 | 0.79 | -0.47 | 0.15 | -1.79 | -1.33 |
| UBE2V1 | Q13404 | 0.96 | -0.42 | -1.54 | -1.44 | -0.37 | -0.55 | -0.30 | -1.49 | -0.03 | -0.21 | -0.54 | -1.63 | -0.46 | 0.56 | 1.37 | 0.80 | 0.76 | 0.58 | -0.10 | 0.51 | -0.04 | 0.89 | -0.53 | -1.27 | 0.49 | 1.26 | 0.50 | -0.05 | 0.67 | 0.35 | -1.32 |
| UBE2V2 | Q15819 | 0.21 | -1.88 | -2.15 | -1.88 | -1.62 | -1.25 | -1.70 | -1.71 | -1.11 | -1.73 | -2.15 | -1.75 | -1.61 | -0.58 | 0.64 | 0.18 | 0.08 | -0.45 | -1.47 | -0.05 | -1.68 | 0.13 | -1.20 | -1.25 | -0.34 | 0.44 | -0.17 | -2.00 | -0.42 | -1.67 | -1.59 |
| UGP2 | Q16851 | 1.10 | -0.68 | -0.72 | -1.61 | -0.82 | -0.93 | -0.74 | -0.97 | 0.08 | -0.70 | -1.44 | -1.47 | -0.90 | 0.86 | 0.63 | 0.77 | 0.84 | 0.35 | -0.26 | 1.41 | 0.34 | 0.23 | -0.78 | -0.33 | 1.07 | 0.49 | 1.16 | 0.09 | 0.82 | -0.49 | -1.32 |
| USP14 | P54578 | 0.40 | -1.05 | -1.78 | -1.62 | -1.27 | -1.74 | -0.78 | -1.72 | -1.72 | -1.73 | -2.03 | -1.60 | -1.58 | -0.16 | 1.23 | 0.34 | 0.20 | -0.19 | -1.44 | 0.52 | -0.80 | 0.56 | -1.64 | -1.16 | -0.61 | 1.05 | -0.31 | -0.92 | 0.23 | -0.21 | -1.70 |
| VCL | P18206 | 0.91 | -0.49 | -0.79 | -0.71 | 0.39 | 1.27 | -0.32 | -1.12 | -0.20 | -1.93 | 0.40 | -0.07 | -1.06 | 0.73 | 0.50 | 0.90 | 1.07 | 0.54 | 0.29 | 1.24 | 0.35 | -0.05 | -0.26 | -0.23 | 0.68 | 0.56 | 0.68 | -0.04 | 0.88 | -0.15 | -1.48 |
| VILL | O15195 | -0.15 | -1.79 | -1.39 | -1.27 | -1.49 | -1.96 | -1.28 | -1.88 | -0.39 | -1.67 | -2.09 | -1.49 | -1.54 | -0.35 | 1.58 | 0.77 | 0.40 | 0.18 | -1.93 | 0.59 | 0.34 | 1.65 | -1.79 | -1.35 | -1.79 | 1.86 | -0.10 | 0.41 | 0.52 | 0.26 | -1.52 |
| VPS26A | O75436 | 0.20 | -0.53 | -0.65 | -0.28 | -0.89 | -0.38 | -1.56 | -2.11 | -0.67 | -0.61 | -1.93 | -2.16 | -1.53 | 0.30 | -1.67 | 0.00 | -0.14 | -0.33 | -1.27 | 0.49 | -0.06 | 0.01 | -1.69 | -1.54 | 0.27 | -0.07 | 0.04 | -0.09 | -0.38 | -1.10 | -1.37 |
| VPS4B | O75351 | 0.32 | -0.25 | -1.27 | -1.10 | -1.78 | -1.43 | -1.77 | -1.70 | -1.10 | -0.65 | -1.66 | -1.72 | -0.07 | 0.39 | 0.08 | -0.12 | 0.71 | 0.27 | -0.24 | 0.84 | -0.08 | 0.14 | -1.98 | 0.11 | 0.93 | -0.07 | 0.88 | -0.22 | 0.37 | -1.32 | -0.76 |
| WARS1 | P23381 | 1.17 | -0.54 | -1.76 | -0.11 | -0.86 | -1.13 | -1.48 | -1.99 | -1.42 | -0.95 | -1.86 | -1.64 | -0.80 | 0.85 | 0.40 | 0.72 | 0.28 | -0.20 | -0.40 | 0.44 | -0.23 | 0.20 | -1.91 | -1.58 | 0.55 | 0.24 | 0.48 | -0.48 | 0.53 | -1.53 | -1.60 |
| WDHD1 | O75717 | -0.48 | 0.17 | -2.18 | 0.28 | -1.58 | -1.52 | -1.42 | 0.32 | 0.42 | -1.59 | -1.44 | -1.43 | -1.98 | 0.01 | 0.15 | 0.84 | -0.07 | -1.88 | 1.05 | 0.01 | 0.42 | 1.15 | -0.73 | -1.82 | 0.41 | 0.72 | 0.34 | -2.01 | 1.40 | -1.27 | 0.74 |
| WDR1 | O75083 | 1.49 | 0.28 | 0.37 | -0.38 | 0.13 | 0.40 | 0.17 | -0.57 | -0.03 | 0.08 | -0.21 | 0.04 | 0.37 | 1.35 | 0.82 | 0.49 | 1.28 | 0.77 | 0.33 | 1.61 | 0.81 | 0.50 | -0.01 | 0.21 | 1.68 | 0.77 | 1.58 | 0.48 | 0.96 | 0.02 | -0.50 |
| XPO1 | O14980 | -0.15 | -0.14 | -1.45 | -1.98 | -0.93 | -1.76 | -1.36 | -2.00 | -2.18 | -1.67 | -1.27 | -1.39 | -1.77 | 0.13 | 0.19 | -0.08 | -0.40 | -1.67 | -1.53 | 0.33 | -0.61 | -0.40 | -1.81 | -1.73 | -0.07 | 0.04 | -0.16 | -1.72 | 0.13 | -1.57 | -1.28 |
| ZNF324 | O75467 | 0.41 | -1.41 | -1.32 | -1.60 | -1.67 | -1.54 | -1.81 | -1.81 | -1.24 | -1.53 | -1.77 | -1.30 | -1.74 | -0.22 | 1.12 | 0.40 | -0.13 | -0.50 | -1.08 | 0.28 | -0.56 | 0.24 | -1.52 | -0.59 | -0.20 | 0.75 | -1.37 | -1.55 | -0.17 | -0.43 | -1.58 |

Supplementary Table S2. Differentially expressed proteins between carcinoma and presumed benign samples identified in pancreatic cyst fluid. Pancreatic cyst fluid (PCyF) samples (n=31) were analyzed in a blinded manner using data-independent acquisition (DIA) mass spectrometry (MS) to acquire label-free quantitation (LFQ) intensity values. The LFQ values were acquired upon normalization of raw label-free quantitative nano-liquid chromatography-tandem MS data (peptide intensities) for all identified proteins in all samples (Materials and methods). A generic multi-protein panel of 377 differentially expressed proteins (DEPs) identified in PCyF was derived upon statistical analysis of differences in protein abundance, represented by Z-score values of normalized, $log_2$-transformed LFQ intensities, between carcinoma and presumed benign groups of the training set (two-tailed Student's t-test; FDR-adjusted p-value <0.05; S0=2.00; Fig. S2A). A final multiple-protein panel consisting of 89 DEPs (highlighted in grey background) was obtained using only DEPs reduced abundance in carcinomas (n=3) and those specifically detected in all carcinomas of the training set, but not in any presumed benign samples (n=86), as described in Fig. S1).

| Genes | Protein | Literature curation | | | | | | | Database curation | | Author assessment | |
|---|---|---|---|---|---|---|---|---|---|---|---|---|
| | | Association reported | mRNA expression | Protein expression | Cell type | Cancer cell process | Signaling pathway | PMID | GO Biological process (GO Accession; GO Evidence code) | GO Molecular function | Protein family | Cancer Hallmark |
| ALOX5 | Polyunsaturated fatty acid 5-lipoxygenase | Yes | Upregulation in PC tissue; Mainly expressed in macrophages infiltrating in PC tissue; High expression is associated with poor patient survival in PC | Upregulation in PC tissue; Expression in PC cell lines; Expression in macrophages | Cancer; Macrophages | Positive regulation of the proliferation, migration, invasion and expression of mesenchymal marker Vimentin, and negative regulation in expression of epithelial marker E-cadherin in PC cell lines | N/A | 36660936; 37348233 | leukocyte chemotaxis involved in inflammatory response (GO:0002232)<br>inflammatory response (GO:0006954; IDA; IMP; ISS)<br>regulation of inflammatory response (GO:0050727; IDA; IMP; ISS)<br>leukotriene biosynthetic process (GO:0019370; IDA; IMP) | oxidoreductase activity (GO:0016491; IDA; IBA) | Enzyme | Tumor Promoting Inflammation |
| AMY2A | Pancreatic alpha-amylase | Yes | Downregulation in acinar cells during acinar to ductal metaplasia; Downregulation in dedifferentiatied acinar cells which highly express genes associated with PC | Downregulation in PCyF from high grade dysplastic cysts/carcinomas compared to benign/low grade dysplastic cysts | Epithelial | N/A | N/A | 37425649; 34049974; 34459141 | carbohydrate metabolic process (GO:0005975; IBA) | hydrolase activity (GO:0016787; IDA; IBA) | Enzyme | N/A |
| AMY2B | Alpha-amylase 2B | Yes | Downregulation in PC tissue; Upregulation in aberrantly differentiated endocrine-exocrine (ADEX) tumor subtype | Downregulation in PCyF from high grade dysplastic cysts/carcinomas compared to benign/low grade dysplastic cysts | Epithelial | N/A | N/A | 31394630; 26909576; 34459141 | carbohydrate metabolic process (GO:0005975; IBA) | hydrolase activity (GO:0016787; IDA; IBA) | Enzyme | N/A |
| ANXA11 | Annexin A11 | Yes | N/A | Upregulation in PC tissue; Upregulation in PC-derived serum exosomes; Upregulation of protein levels in plasma EVs from PC patients; High protein expression in EVs from PC-derived organoids; Expression in PC cell lines | N/A | N/A | N/A | 30160795; 32990680 | cell division (GO:0051301; IMP)<br>cell cycle process (GO:0022402; IMP) | RNA binding (GO:0003723; HDA) | Transcriptional/Translational regulator | Sustaining proliferative signaling |
| ARHGDIB | Rho GDP-dissociation inhibitor 2 | Yes | Upregulation in PC tissue; High expression is associated with poor patient survival in PC | Upregulation in PC tissue; Upregulation in PC cells invading the perineural niche of PC tissue; Expression in PC cell lines; High expression is associated with poor patient survival in PC | Cancer | Positive regulation of the proliferation, migration, invasion (including perineural) and expression of mesenchymal marker Vimentin, and negative regulation in expression of epithelial marker E-cadherin in PC cell lines | N/A | 19509238; 35147897; 24788627; 25573518 | signal transduction (GO:0007165; IMP; IDA; IBA) | hydrolase activity (GO:0016787; IMP; IDA; IBA) | Enzyme | N/A |
| ARPC1B | Actin-related protein 2/3 complex subunit 1B | Yes | Expression in PC cell lines | Upregulation in PC tissue lysates | Cancer | Positive regulation of the migration and invasion of PC line | N/A | 19145645; 33388318 | cytoskeleton organization (GO:0007010; IDA; IBA) | structural constituent of cytoskeleton (GO:0005200; IDA)<br>actin binding (GO:0003779; IDA) | Structural protein | N/A |
| ARPC3 | Actin-related protein 2/3 complex subunit 3 | Yes | Expression in PC cell lines | Upregulation in PC tissue lysates; High expression in acinar-to-ductal metaplasia lesions and in PC tissue | Cancer/Epithelial | Positive regulation of the migration of PC line | N/A | 23267127; 33388318 | cytoskeleton organization (GO:0007010; IDA; IBA) | structural constituent of cytoskeleton (GO:0005200; IDA)<br>actin binding (GO:0003779; IDA; IBA) | Structural protein | N/A |
| BAX | Apoptosis regulator BAX | Yes | Upregulation in PC tissue; Expression in cancer cells (especially in those forming duct-like structures) of PC tissue; Expression in PC cell lines; High expression is associated with longer patient survival in PC | Upregulation in PC tissue; Expression in PC cell lines; High expression is associated with longer patient postoperative survival in PC | Cancer/Epithelial | Positive regulation of apoptosis of PC cell line | N/A | 9863489; 27074573 | regulation of apoptotic process (GO:0042981; IDA; IMP; IBA)<br>regulation of programmed cell death (GO:0043067; IDA; IMP; IBA) | channel activity (GO:0015267; IDA; IBA)<br>transporter activity (GO:0005215; IDA; IBA) | Transporter | Resisting cell death |
| CASP6 | Caspase-6 | Yes | Expression in PC cell lines; Low expression is associated with longer patient survival in PC | Expression in PC cell lines | Cancer | Positive regulation of the proliferation, migration and invasion of PC cell lines | N/A | 36882663; 32682809 | regulation of apoptotic process (GO:0042981; IDA)<br>regulation of programmed cell death (GO:0043067; IDA)<br>epithelial cell differentiation (GO:0030855; IEP) | hydrolase activity (GO:0016787; IDA) | Enzyme | Resisting cell death/ Unlocking phenotypic plasticity |
| CDA | Cytidine deaminase | Yes | Upregulation in PC tissue; Expression in PC cell lines; High expression is associated with poor patient survival in PC; Upregulation in PC cell line upon incubation with tumor-activated macrophage-conditioned medium in the presence of gemcitabine | Expression in PC cell lines; *Upregulation after gemcitabine treatment in orthotopic tumor mouse model; Downregulation in macrophage-depleted mice treated with gemcitabine* | Cancer | Positive regulation of the proliferation, colony formation, cell clycle regulation and negative regulation of apoptosis in PC cell lines | N/A | 23995783; 31871136; 38294491; 36084875 | regulation of cell growth (GO:0001558; IDA) | hydrolase activity (GO:0016787; IDA; IBA) | Enzyme | Evading growth suppressors |
| CDH17 | Cadherin-17 | Yes | Expression in PC cell lines | Expression in in primary and metastatic PC tissue; Expression in sclerosing and non-sclerosing pancreatic neuroendocrine tumour (PanNET) tissue; Expression in exocrine-like tumors from PC tissue; Expression in PC cell lines | Cancer/Epithelial | Positive regulation of the proliferation, colony formation and migration of PC cell lines | N/A | 22904132; 31004701; 25307987; 27846146 | cell migration (GO:0016477; IBA)<br>cell-cell junction organization (GO:0045216; IBA)<br>cell-cell adhesion (GO:0098609; IBA; ISS) | cell adhesion molecule binding (GO:0050839; IPI; IBA) | Structural protein | Activating invasion and metastasis |
| CMPK1 | UMP-CMP kinase | Yes | Single nucleotide polymorphisms of CMPK1 involved in gemcitabine metabolic pathway and associated with overall survival, time-to-progression and disease progression of PC patients treated with gemcitabine-based chemotherapy | Upregulation in gemcitabine clone of PC cells; High expression is associated with poor patient survival in PC | Cancer | N/A | N/A | 22838950; 36084875 | nucleobase-containing compound metabolic process (GO:0006139; IDA) | kinase activity (GO:0016301; EXP; IDA; IBA) | Enzyme | N/A |
| COMT | Catechol O-methyltransferase | Yes | Expression in PC cell lines | Upregulation in PC tissue; Expression in PC cell lines; High expression PanIN tissue; Expression in tissues increases with pathological stage (from normal tissue to PanIN lesions to PC); Expression in ductal epithelia and pancreatitis tissue; High expression is associated with longer patient survival in PC | Cancer | Negative regulation of the proliferation, cell clycle (S phase) regulation and positive regulation of apoptosis in PC cell lines | PI3K/Akt | 23015122; 25711924; DOI: 10.1097/JP9.0000000000000006 | catecholamine metabolic process (GO:0006584; IDA; IBA) | transferase activity (GO:0016740; ISS; IBA) | Enzyme | N/A |
| CSK | Tyrosine-protein kinase CSK | Yes | Upregulation in PC tissue | N/A | Cancer | *Negative regulation of the colony formation of tumor-derived cells acquired from orthotopic pancreatic carcinoma mouse model* | N/A | 21242978; 38235873 | regulation of immune response (GO:0050776; IBA)<br>immune response (GO:0006955; IBA) | kinase activity (GO:0016301; IPI; IBA) | Enzyme | Tumor Promoting Inflammation |
| CTNNB1 | Catenin beta-1 | Yes | N/A | Expression in PC tissue; Low protein expression is associated with higher tumor histological grade in PC tissue; Low protein expression is associated with with poor patient survival in PC; High protein expression is associated with better prognosis and lower risk of death from PC. *Upregulation caused ductal lesions and increased pancreas mass and induced tumorigenesis in pancreatic carcinoma mouse model; Conditional knockout suppressed the tumorigenesis and growth of pancreatic carcinomas, and significantly prolongs the survival time in mice* | Cancer/Epithelial | *Positive regulation of the proliferation of tumor-derived cells acquired from mouse model ; Positive regulator of tumorigenesis in mouse models* | Wnt;Notch | 27411302; 18725219; 25517963 | regulation of epithelial to mesenchymal transition (GO:0010717; IGI; IMP)<br>cell-cell junction organization (GO:0045216; IMP)<br>cell-cell adhesion (GO:0098609; IMP; IBA)<br>regulation of apoptotic process (GO:0042981; IDA)<br>regulation of programmed cell death (GO:0043067; IDA) | transcription regulator activity (GO:0140110; IMP; IDA) | Transcriptional/Translational regulator | Unlocking phenotypic plasticity/Activating invasion and metastasis |
| CYBA | Cytochrome b-245 light chain | Yes | Upregulation in PC tissue; High expression in IPMNs compared to nonneoplastic pancreatic ductal epithelium | N/A | N/A | N/A | N/A | 14982844; 37222951; 15477757 | inflammatory response (GO:0006954; IMP)<br>innate immune response (GO:0045087; IMP; ISS)<br>regulation of phagocytosis (GO:0050764; IDA) | oxidoreductase activity (GO:0016491; IMP) | Enzyme | Tumor Promoting Inflammation |

| | | | | | | | | | | | |
|---|---|---|---|---|---|---|---|---|---|---|---|
| CYRIB/FAM49B | CYFIP-related Rac1 interactor B | Yes | Low expression in PC tissue; Expression in PC cell lines | Low expression in PC tissue; Low expression is associated with higher tumor lymph node invasion in PC; Expression in PC cell lines | Cancer/Epithelial | Negative regulation of the proliferation, migration, invasion and intracellular reactive oxygen species (ROS) in PC cell lines | PI3K/Akt;MAPK/ERK | 29059164 | regulation of leukocyte activation (GO:0002694; IBA; ISS)<br>regulation of leukocyte mediated immunity (GO:0002703; ISS) | major histocompatibility complex (MHC) protein complex binding (GO:0023023; ISS)<br>GTPase binding (GO:0051020; IPI) | Signal transducer/modulator | Tumor Promoting Inflammation |
| DBNL | Drebrin-like protein | Yes | Upregulation in PC tissue of patients with lymph node metastasis | N/A | N/A | N/A | N/A | 17414055 | cell junction organization (GO:0034330; ISS) | cell adhesion molecule binding (GO:0050839; HDA)<br>actin binding (GO:0003779; ISS) | Structural protein | Activating invasion and metastasis |
| EIF3B | Eukaryotic translation initiation factor 3 subunit B | Yes | Expression in PC tissue; Expression in PC cell lines | Upregulation in PC tissue; Expression in PC cell lines; High expression is associated with higher tumor histological grade in PC tissue; High exoression is associated with poor patient survival in PC | Cancer | Positive regulation of the proliferation, migration, cell cycle arrest and negative regulation of apoptosis in PC cell lines | PI3K/Akt | 33996561; 33459066 | regulation of translational initiation (GO:0006446; IDA)<br>translational initiation (GO:0006413; IDA; IBA) | translation initiation factor activity (GO:0003743; IDA; IBA)<br>RNA binding (GO:0003723; IDA) | Transcriptional/Translational regulator | Sustaining proliferative signaling |
| ERLIN2 | Erlin-2 | Yes | Expression in PC tissue; Member of a gene signature which distinguish normal from PC tissues samples; High expression of signature ERLIN2 belongs to is associated with poor patient survival in PC | N/A | Cancer | N/A | N/A | 27240826 | cellular response to stress (GO:0033554; IMP; IDA; IBA)<br>signal transduction (GO:0007165; IMP; IBA)<br>regulation of cholesterol metabolic process (GO:0090181; IDA; IMP) | enzyme binding (GO:0019899; IPI)<br>Lipid binding (GO:0008289; IBA) | Signal transducer/modulator | Deregulating cellular energetics |
| EWSR1 | RNA-binding protein EWS | Yes | Expression in PC tissue; EWSR1 fusions PanNETs associated with mTOR pathway activation; *EWSR1-FLI1 fusion oncogene expression is associated with loss of the acinar cells and induces acinar-to-ductal metaplasia in the pancreas of mice* | N/A | N/A | N/A | mTOR | 28199314; 33051146 | N/A | RNA binding (GO:0003723; HDA; IBA)<br>transcription regulator activity (GO:0140110; IBA) | Transcriptional/Translational regulator | N/A |
| GART | Trifunctional purine biosynthetic protein adenosine-3 | Yes | Expression in PC tissue; *Upregulated in pancreatic cancer cells grown as tumors (in pancreatic carcinoma mouse model) compared to cell cultured in vitro (possible association with the tumor environment)* | N/A | N/A | N/A | N/A | 34447748; 33152324 | nucleobase-containing compound metabolic process (GO:0006139; IDA; IBA) | ligase activity (GO:0016874; IDA; IBA)<br>transferase activity (GO:0016740; IDA; IBA) | Enzyme | N/A |
| GLRX | Glutaredoxin-1 | Yes | Expression in PC cell lines; Upregulation in PC cell line upon treatment with DNA methyltransferase inhibitor (5Aza-dC) or histone deacetylase inhibitor (SAHA) | Upregulation in PC tissue | N/A | N/A | N/A | 10757123; 19035457 | N/A | oxidoreductase activity (GO:0016491; EXP; IBA) | Enzyme | N/A |
| GPX4 | Phospholipid hydroperoxide glutathione peroxidase | Yes | Expression in PC cell lines | Expression in PC cell lines; Upregulation in PC cell lines upon tratment with gemcitabine | Cancer | Negative regulation of the proliferation and positive regulation of apoptosis and ferroptosis in PC cell lines | mTOR; STAT3; HSPA5 | 32457486; 35859150; 28130223 | regulation of programmed cell death (GO:0043067; ISS) | oxidoreductase activity (GO:0016491; IMP; IDA; IBA) | Enzyme | Resisting cell death |
| GRB2 | Growth factor receptor-bound protein 2 | Yes | Expression in PC cell lines | Expression in PC tissue; Expression in PC cell lines; High expression is associated with poor patient survival in PC | Cancer | Positive regulation of the proliferation, colony formation, migration and cell cycle regulation in PC cell lines | PI3K/Akt; MAPK/ERK; JNK | 26885689; 33987255; 21508312 | leukocyte mediated immunity (GO:0002443)<br>innate immune response (GO:0045087; IDA)<br>DNA damage response (GO:0006974; IMP) | signaling receptor binding (GO:0005102; IPI; IBA)<br>GTPase regulator activity (GO:0030695; IDA) | Signal transducer/modulator | Avoiding immune destruction/ Genome Instability and Mutation |
| HINT1 | Histidine triad nucleotide-binding protein 1 | Yes | N/A | Downregulation in PC tissue | N/A | N/A | N/A | 31927104 | programmed cell death (GO:0012501; IMP) | hydrolase activity (GO:0016787; IMP; IDA; IBA) | Enzyme | N/A |
| HNRNPL | Heterogeneous nuclear ribonucleoprotein L | Yes | Upregulation in PC tissue; Expression in PC cell lines | Expression in cells of PC tissue; Expression in PC cell lines; Positive correlation with tumor TNM stages and pathologic grade; High expression is associated with poor patient survival in PC | Cancer | Positive regulation of the colony formation, migration, cell cycle regulation and mesenchymal markers N-cadherinin expression in PC cell lines | MAPK/ERK | 31355266; 27689400; 34779408 | regulation of RNA splicing (GO:0043484; IMP; IBA) | RNA binding (GO:0003723; HDA; IDA; IBA)<br>transcription regulatory region nucleic acid binding (GO:0001067; IDA) | Transcriptional/Translational regulator | N/A |
| ITGA2 | Integrin alpha-2 | Yes | Upregulation in PC tissue; High expression is associated with poor survival in PC; High expression is associated with poor survival and disease-free survival in gemcitabine-treated PC patients | Upregulation in PC tissue; Expression in PC cell lines; High expression is associated with poor patient survival, positive lymph node status, high tumour grade (G3/G4) and perineural invasion in PC; High expression is associated with poor survival, disease-free survival, increased risk of death and increase of relapse in gemcitabine-treated PC patients; High protein is associated with poor survival and progression-free survival survival in metastatic gemcitabine-treated PC patients; Expression is correlated with TNM stage, lymph node metastasis and local invasion in PC | Cancer | Positive regulation of the proliferation, colony formation, migration, invasion, DNA repair and expression of mesenchymal markers (E-cadherin, N-cadherin, Snail) and negative regulation of apoptosis in PC cell lines | PI3K/Akt; STAT3;FAK; NHEJ | 35396216; 36765586; 35193647; 34007003; 30569514; 37881431; 35998796; 31818309; 37907515 | cell-matrix adhesion (GO:0007160; IBA; IMP)<br>cell-cell adhesion (GO:0098609; IBA)<br>cell migration (GO:0016477; IMP) | cell adhesion molecule binding (GO:0050839; IMP; IBA) | Structural protein | Activating invasion and metastasis |
| KPNB1 | Importin subunit beta-1 | Yes | N/A | Expression in PC cell lines | Cancer | Positive regulation of the proliferation and migration of PC cell lines | N/A | 28811376; 38195622 | mitotic cell cycle process (GO:1903047; IMP)<br>cell cycle process (GO:0022402; IMP) | RNA binding (GO:0003723; HDA)<br>nuclear import signal receptor activity (GO:0061608; IDA; IBA) | Transcriptional/Translational regulator | Evading growth suppressors |
| LITAF | Lipopolysaccharide-induced tumor necrosis factor-alpha factor | Yes | Downregulation in PC tissue; Expression in PC cell lines; Upregulation in metastatic compared to primary PC tissue | Upregulation in PC tissue; Upregulation in metastatic compared to primary PC tissue and in PC patients with lymph node metastasis; Low expression in PC tissue is associated with poor disease-free survival | N/A | Negative regulation of the proliferation and positive regulation of apoptosis and cell cycle arrest in PC cell lines | N/A | 29423035; 37612267 | regulation of cytokine production (GO:0001817; IBA) | transcription regulatory region nucleic acid binding (GO:0001067; IDA)<br>DNA-binding transcription factor activity (GO:0003700; IMP) | Transcriptional/Translational regulator | Tumor Promoting Inflammation |
| LYPLA1 | Acyl-protein thioesterase 1 | Yes | Upregulation in PC tissue | N/A | N/A | N/A | N/A | 30137376 | regulation of autophagy (GO:0010506; IDA) | hydrolase activity (GO:0016787; IDA; IBA) | Enzyme | Resisting cell death |
| MAPK14 | Mitogen-activated protein kinase 14 | Yes | Upregulation in PC tissue; Upregulation in peripheral blood of PC patients; High expression is increasing with advanced stage and associated with poor survival in PC | Upregulation in PC tissue; Upregulation of phospho-MAPK14 in PC tissue; Expression in PC cell lines | Cancer | Positive regulation of the proliferation, colony formation, migration, invasion, cell cycle progression and negative regulation of apoptosis in PC cell lines | N/A | 31828036; 34194669; 35449823; 32988878; 38285896; 38395519; DOI:10.1016/j.imu.2020.100346 | regulation of cytokine production involved in inflammatory response (GO:1900015; IDA)<br>regulation of inflammatory response (GO:0050727; IDA)<br>cell differentiation (GO:0030154; IMP; ISS)<br>DNA damage response (GO:0006974; IMP) | kinase activity (GO:0016301; IDA; IBA) | Enzyme | Tumor Promoting Inflammation/ Unlocking phenotypic plasticity/ Genome Instability and Mutation |

| | | | | | | | | | | | | |
|---|---|---|---|---|---|---|---|---|---|---|---|---|
| MNDA | Myeloid cell nuclear differentiation antigen | Yes | Methylation in or near MNDA gene was significantly associated with pancreatic cancer risk | N/A | N/A | N/A | N/A | 32430337 | regulation of apoptotic process (GO:0042981; IMP) regulation of programmed cell death (GO:0043067; IMP) regulation of immune response (GO:0050776; IBA) innate immune response (GO:0045087; IBA) DNA damage response (GO:0006974; IMP) | DNA binding (GO:0003677; IBA) | Transcriptional/Translational regulator | Resisting cell death/ Tumor Promoting Inflammation/ Genome Instability and Mutation |
| MTAP | S-methyl-5'-thioadenosine phosphorylase | Yes | Deep gene deletion is prevalent in PC patients and associated with poor survival in PC | Downregulation in PC tissue; Loss of MTAP activity in PC tissue; Expression in PanIN tissue and PC cell lines; Loss of expression is associated with poor survival in PC | Cancer/Epithelial | Positive regulation of the proliferation, colony formation, DNA repair, mitochondrial respiration, metabolic reprogramming (by enhancing the Warburg effect) and negative regulation of glycolysis in PC cell lines | RioK1 | 15662124; 15534104; 15832197; 38054349; 34385182; 31257072 | N/A | transferase activity (GO:0016740; IBA) | Enzyme | N/A |
| MUC4 | Mucin-4 | Yes | N/A | Upregulation in PC tissue; Absence in normal pancreatic tissue; Expression is increased with increasing grade of PanIN; Progressive increase in expression from PanIN-I to PDAC; High expression is associated with poor survival in PC; Low expression is associated with favorable survival in PC patients receiving adjuvant gemcitabine treatment | Cancer | Positive regulation of the proliferation, colony formation, migration, invasion, epithelial-to-mesenchymal transition and negative regulation of apoptosis in PC cell lines | PI3K/Akt; MAPK/ERK; JNK; NF-κB | 28460571; 17019794; 16049287; 12090430; 14744777; 17595659; 17406026; 22580602; 26035354; 18381409; 19738614; 28270046; 16049287; 27185392 | epithelial structure maintenance (GO:0010669; IMP) | signaling receptor binding (GO:0005102; IBA) | Signal transducer/modulator | Unlocking phenotypic plasticity |
| NHERF1 | Na(+)/H(+) exchange regulatory cofactor NHE-RF1 | Yes | Downregulation in PC tissue | Downregulation in PC tissue; Expression in PC cell lines | Cancer | Negative regulation of the proliferation, colony formation, cell cycle progression, migration, invasion, and positive regulation of apoptosis in PC cell lines | PI3K/Akt; Wnt | 22622406; 23544174; 25120624; 26622484; 29725468 | regulation of cell population proliferation (GO:0042127; IDA; IMP) regulation of cell cycle (GO:0051726) regulation of apoptotic process (GO:0042981; IDA) epithelial cell differentiation (GO:0030855; IMP; ISS) | signaling receptor binding (GO:0005102; IPI; IBA) | Signal transducer/modulator | Sustaining proliferative signaling/ Resisting cell death/ Unlocking phenotypic plasticity |
| NIT2 | Omega-amidase NIT2 | Yes | N/A | Downregulation in the secretome of PC cell lines | Cancer | N/A | N/A | 20687567 | asparagine metabolic process (GO:0006528; IDA; IBA) | hydrolase activity (GO:0016787; IDA; IBA) | Enzyme | N/A |
| PGLYRP1 | Peptidoglycan recognition protein 1 | Yes | Upregulation in PC tissue | Upregulation in serum of PDAC patients; Variable expression PC tissue and absence of expression in healthy tumour-adjacent pancreas; Expression in primary PDAC cells | Cancer/Epithelial | Negative regulation of the phagocytosis and T cell cytotoxic killing (immune evasion) in primary PDAC cells | NF-κB | 37529485; 38754953 | humoral immune response (GO:0006959; IDA) immune response (GO:0006955; IDA) | signaling receptor activity (GO:0038023; IDA; IBA | Signal transducer/modulator | Tumor Promoting Inflammation |
| PGM2 | Phosphoglucomutase-2 | Yes | Upregulation in PC tissue | Expression in PC cell lines | Cancer | Positive regulation of the metabolic activity (glucose and uridine metabolism) in PC cell line | N/A | 37198494 | nucleobase-containing compound metabolic process (GO:0006139; IBA) | isomerase activity (GO:0016853; IDA; IBA) | Enzyme | N/A |
| PLA2G1B | Phospholipase A2 | Yes | Downregulation in PC tissue and PC cell lines | N/A | N/A | N/A | N/A | 29288362; 32272917 | innate immune response in mucosa (GO:0002227; IDA) innate immune response (GO:0045087; IDA) humoral immune response (GO:0006959; IDA) regulation of immune response (GO:0050776; IDA; IBA) immune response (GO:0006955; IDA; IBA) leukocyte mediated immunity (GO:0002443; ISS) leukotriene biosynthetic process (GO:0019370; ISS) positive regulation of cell population proliferation (GO:0008284; IDA; IBA) | hydrolase activity (GO:0016787; IDA; IBA) signaling receptor binding (GO:0005102; IPI; IDA; IBA) | Enzyme | Tumor Promoting Inflammation/ Sustaining proliferative signaling |
| PLBD1 | Phospholipase B-like 1 | Yes | Upregulation in PC tissue; Upregulation in blood of PC patients; High expression is associated with poor survival in PC | Expression in PC cell lines | Cancer | N/A | N/A | 33133160; 21653254; 29617451 | lipid metabolic process (GO:0006629; IBA) | hydrolase activity (GO:0016787; IBA) | Enzyme | N/A |
| PLCB3 | 1-phosphatidylinositol 4,5-bisphosphate phosphodiesterase beta-3 | Yes | Gene mutation (missense) in PC tissue | N/A | N/A | N/A | N/A | 18772397 | lipid metabolic process (GO:0006629; IDA; IBA) signal transduction (GO:0007165; IBA) | hydrolase activity (GO:0016787; IDA; IBA) cell adhesion molecule binding (GO:0050839; HDA) | Enzyme | N/A |
| PLS1 | Plastin-1 | Yes | Upregulation in PC tissue; High expression is associated with poor survival in PC | Upregulation in PC tissue | Cancer | Positive regulation of the proliferation, migration, invasion, metabolic activity (glucose uptake, pyruvate production, lactate production), expression of glycolysis-related proteins and negative regulation of apoptosis in PC cell lines | N/A | 17412869; DOI: 10.23736/S2724-542X.23.03010-9 | epithelial cell differentiation (GO:0030855; ISS) | actin binding (GO:0003779; IBA) | Structural protein | Unlocking phenotypic plasticity |
| PSMB4 | Proteasome subunit beta type-4 | Yes | N/A | Upregulation in EVs from plasma samples of patients with metastatic PC | N/A | N/A | N/A | 36993200 | proteasome-mediated ubiquitin-dependent protein catabolic process (GO:0043161; IBA) | N/A | N/A | N/A |
| PSMB8 | Proteasome subunit beta type-8 | Yes | Upregulation of lncRNA (PSMB8-AS1) in PC tissue and PC cell lines | N/A | Cancer | Positive regulation of the proliferation, migration, invasion, metabolic activity (glucose uptake, pyruvate production, lactate production), expression of glycolysis- and immune evasion-related proteins and negative regulation of apoptosis in PC cell lines | STAT1 | 29796634; 32891166 | cell differentiation (GO:0030154; IMP) | hydrolase activity (GO:0016787; IBA) | Enzyme | Unlocking phenotypic plasticity |
| PSMB9 | Proteasome subunit beta type-9 | Yes | Upregulation in PC tissue and PC cell lines | N/A | Cancer | N/A | N/A | 34957503 | proteasome-mediated ubiquitin-dependent protein catabolic process (GO:0043161; IBA) | hydrolase activity (GO:0016787; IBA) | Enzyme | N/A |
| PTPRC | Receptor-type tyrosine-protein phosphatase C | Yes | High expression of mRNA levels in tissue of PC patients with prolonged survival; High expression of mRNA levels in dendritic cells, macrophages, natural | Upregulation in PC tissue; High expression in border and distal regions of PC tissue; High expression in leukocyte-derived EVs (CD45+) in blood of PC patients is associated with prolonged survival in PC | Immune cells | N/A | N/A | 31381571; 24652403; 33727309; 36230671 | regulation of immune response (GO:0050776; IMP; ISS) immune response (GO:0006955; IMP; ISS) | signaling receptor binding (GO:0005102; ISS) | Signal transducer/modulator | Tumor Promoting Inflammation/ |

| | | | | | | | | | | | | |
|---|---|---|---|---|---|---|---|---|---|---|---|---|
| | | | killer and T cells of tissue of PC patients with good prognosis | | | | | | regulation of leukocyte activation (GO:0002694; IMP; ISS) regulation of phagocytosis (GO:0050764; IDA; ISS) regulation of cell cycle (GO:0051726; ISS) cell cycle process (GO:0022402; IMP) | | | Sustaining proliferative signaling |
| RAB31 | Ras-related protein Rab-31 | Yes | Upregulation in PC tissue | Upregulation in PC tissue; High expression in blood of PC patients are associated with poor disease-free survival and overall survival | N/A | N/A | N/A | 31258775 | regulation of phagocytosis (GO:0050764; ISS) | GTPase activity (GO:0003924; IBA) | Enzyme | Tumor Promoting Inflammatio n |
| RAC2 | Ras-related C3 botulinum toxin substrate 2 | Yes | Upregulation in PC tissue and metastatic PC tissue; Expression in PC cell lines; *Downregulation inhibited tumor growth and lymph node metastasis in mouse xenograft model* | Upregulation in the secretome (identified in culture medium) of PC cell lines | Cancer | *Positive regulation of tumor growth (proliferation), angiogenesis, invasion and metastasis in pancreatic carcinoma mouse xenograft model* | N/A | 16215274; 33564087; 24770346; 30900162 | regulation of leukocyte chemotaxis (GO:0002688; IMP) regulation of leukocyte migration (GO:0002685; IMP) regulation of cell-substrate adhesion (GO:0010810; IMP) | GTPase activity (GO:0003924; IBA) | Enzyme | Tumor Promoting Inflammatio n |
| RHOG | Rho-related GTP-binding protein RhoG | Yes | Upregulation in human pancreatic epithelial nestin-expressing (HPNE)-KRAS (G12D mutation) cells in response to ethanol conditioning | Expression in PC cell lines and gemcitabine resistant PC cell lines; Upregulation in human pancreatic epithelial nestin-expressing (HPNE)-KRAS (G12D mutation) cells in response to ethanol conditioning | Cancer/Epitheli al | N/A | N/A | 27832197; 35454872; 35617275 | cell migration (GO:0016477; IDA; IMP) | GTPase activity (GO:0003924; IDA; IBA) | Enzyme | Activating invasion and metastasis |
| RNASE3 | Eosinophil cationic protein | Yes | Gene mutation in IPMN tissue (whole-exome sequencing) | N/A | N/A | N/A | N/A | 22355676 | innate immune response in mucosa (GO:0002227; IDA; IBA) innate immune response (GO:0045087; IDA; IBA) humoral immune response (GO:0006959; IDA; IBA) immune response (GO:0006955; IDA; IBA) | hydrolase activity (GO:0016787; IBA) | Enzyme | Tumor Promoting Inflammatio n |
| RPL15 | 60S ribosomal protein L15 | Yes | Downregulation in PC tissue; Expression in PC cell lines; Low expression is associated with poor survival in PC | Downregulation in PC tissue; Expression in PC cell lines; Low expression is associated with poor survival in PC | Cancer | Negative regulation of the invasion and expression of mesenchymal markers, positive regulation of expression of epithelial markers and apoptosis in PC cell lines | N/A | 26498693 | cytoplasmic translation (GO:0002181; IBA) | RNA binding (GO:0003723; HDA; IBA) cell adhesion molecule binding (GO:0050839; HDA) | Transcriptional/Translat ional regulator | N/A |
| RUVBL2 | RuvB-like 2 | Yes | Upregulation in PC tissue; *Upregulation in PDAC tumours compared to normal tissue in allografted mouse model* | Upregulation in PC tissue; Upregulation in EVs of blood from PC patients (PDAC was compared to chronic pancreatitis and IPMN) and in EVs of blood from metastatic PC patients | N/A | N/A | N/A | 35754815; 36993200; 38821858 | DNA damage response (GO:0006974; IDA; ISO) regulation of DNA repair (GO:0006282; IDA) regulation of DNA replication (GO:0006275; IMP) regulation of cell cycle (GO:0051726; IMP) | transcription regulator activity (GO:0140110; IDA) | Transcriptional/Translat ional regulator | Genome Instability and Mutation/ Sustaining proliferative signaling |
| S100A12 | Protein S100-A12 | Yes | | Upregulation in serum of patients with malignant and benign tumours compared to healthy controls; Upregulation in serum of patients developed grade B postoperative pancreatic fistulas (POPFs) | N/A | N/A | N/A | 31582902 | humoral immune response (GO:0006959; IDA; IBA) immune response (GO:0006955; IDA; IBA) cell migration (GO:0016477; IBA) | signaling receptor binding (GO:0005102; IPI; IDA; IBA) | Signal transducer/modulator | Tumor Promoting Inflammatio n/ Activating invasion and metastasis |
| S100P | Protein S100-P | Yes | Upregulation in PanIN, IPMN and PC tissue compared to normal tissue; Upregulation in pancreatic juice of IPMN and PC tissue; Upregulation in IPMN, invasive ductal carcinoma and PanIN cells compared to normal ductal cells; Expression in PC cell lines; High expression is associated with poor survival in PC | Upregulation in PC tissue; Upregulation correlates with increasing grade of PanINs; Expression in PC cell lines and in their culture media; | Cancer | Positive regulation of the proliferation, cell cycle progression, migration, invasion, and negative regulation of apoptosis in PC cell lines | N/A | 12750293; 16061848; 12950018; 11943709; 15632002; 17000674; 37853345 | cell migration (GO:0016477; IMP; IBA) | calcium-dependent protein binding (GO:0048306; IPI; IBA) | Signal transducer/modulator | Activating invasion and metastasis |
| SEPTIN9 | Septin-9 | Yes | Increased gene methylation in PC tissue; Expression is regulated by hypomethylation in PC tissue | N/A | N/A | N/A | N/A | 19497796; 31205116 | cell division (GO:0051301; IBA) cell cycle process (GO:0022402; IBA) | GTPase activity (GO:0003924; IBA) | Signal transducer/modulator | Genome Instability and Mutation |
| SNRPN | Small nuclear ribonucleoprot ein-associated proteins B and B' | Yes | Upregulation in PC tissue | Expression in PC cell lines | Cancer | Positive regulation of the proliferation, colony formation, cell cycle progression, and negative regulation of apoptosis in PC cell lines | N/A | 26261020; 32448196 | RNA processing (GO:0006396; IBA) | ribonucleoprotein complex binding (GO:0043021; IBA) | Transcriptional/Translat ional regulator | N/A |
| SSBP1 | Single-stranded DNA-binding protein, mitochondrial | Yes | N/A | Expression in PC cell lines | Cancer | N/A | N/A | 32764385 | regulation of DNA replication (GO:0006275; IDA) | DNA binding (GO:0003677; IMP; IDA; IBA) RNA binding (GO:0003723; HDA) | Transcriptional/Translat ional regulator | Sustaining proliferative signaling |
| STXBP2 | Syntaxin-binding protein 2 | Yes | N/A | Upregulation in serum of PC patients | N/A | N/A | N/A | 19199705 | regulation of leukocyte mediated immunity (GO:0002703; IMP; ISS) leukocyte activation involved in immune response (GO:0002366; ISS) | SNARE binding (GO:0000149; IPI; IBA) | Vesicle trafficking regulator | Tumor Promoting Inflammatio n |
| TARS1 | Threonine--tRNA ligase 1, cytoplasmic | Yes | N/A | Upregulation in PC tissue; High expression is associated with poor survival in PC | Cancer | Positive regulation of the migration of PC cell line | N/A | 29328069 | RNA metabolic process (GO:0016070; IDA; IBA; ISS) | ligase activity (GO:0016874; ISS; IDA; IBA) | Enzyme | N/A |
| TSN | Translin | Yes | N/A | Downregulation in invaded and noninvaded nerves of PC tissue | N/A | N/A | N/A | 30690892 | RNA metabolic process (GO:0016070; IDA) | RNA binding (GO:0003723; IBA) | Transcriptional/Translat ional regulator | N/A |
| TSPO | Translocator protein | Yes | N/A | Upregulation in PC tissue; Upregulation in IPMN tissue compared to benign lesions; Expression correlates directly with the grade of dysplasia across low, intermediate, and high-grade samples; Expression in PC cell lines | Cancer | N/A | N/A | 34390852; 32933996 | N/A | lipid binding (GO:0008289; IBA) | N/A | N/A |
| UBE2D3 | Ubiquitin-conjugating enzyme E2 D3 | Yes | Upregulation in PC tissue; High expression is associated with poor survival in PC | Expression in PC cell lines | Cancer | Positive regulation of the proliferation, colony formation, cell cycle progression, and negative regulation of apoptosis in PC cell lines | NF-κB | 35272681 | proteasome-mediated ubiquitin-dependent protein catabolic process (GO:0043161; IDA) | transferase activity (GO:0016740; IDA; IBA) | Enzyme | N/A |
| UBE2I | SUMO-conjugating enzyme UBC9 | Yes | Upregulation in PC tissue; High expression is associated with poor survival in PC | N/A | N/A | N/A | N/A | 32001555 | regulation of cell migration (GO:0030334; IMP) | transcription factor binding (GO:0008134; IPI) | Transcriptional/Translat ional regulator | Activating invasion and metastasis |
| VASP | Vasodilator-stimulated phosphoprotein | Yes | High expression is associated with poor survival in PC | Upregulation in PC tissue; Upregulation in PC tissue is associated with lymphovascular and perineural invasion | Cancer | Positive regulation of size (proliferation) of PC spheroids | FAK | 32001555 | morphogenesis of an epithelium (GO:0002009; IBA) | cell adhesion molecule binding (GO:0050839; HDA) | Structural protein | Unlocking phenotypic plasticity |

| | | | | | | | | | | | | |
|---|---|---|---|---|---|---|---|---|---|---|---|---|
| ABRACL | Costars family protein ABRACL | No | N/A | N/A | N/A | N/A | N/A | N/A | regulation of actin filament-based process (GO:0032970; IBA) | N/A | Structural protein | N/A |
| ADK | Adenosine kinase | No | N/A | N/A | N/A | N/A | N/A | N/A | nucleobase-containing compound metabolic process (GO:0006139; IBA) | kinase activity (GO:0016301; IBA) RNA binding (GO:0003723; HDA) | Enzyme | N/A |
| ALDH9A1 | Aldehyde dehydrogenase 9 family member A1 | No | N/A | N/A | N/A | N/A | N/A | N/A | cellular metabolic process (GO:0044237; IDA) | oxidoreductase activity (GO:0016491; IDA; IBA) | Enzyme | N/A |
| ALOX5AP | Arachidonate 5-lipoxygenase-activating protein | No | N/A | N/A | N/A | N/A | N/A | N/A | leukotriene biosynthetic process (GO:0019370; IDA; IBA) | transferase activity (GO:0016740; IBA) oxidoreductase activity (GO:0016491; IBA) lyase activity (GO:0016829; IBA) | Enzyme | Tumor Promoting Inflammation |
| ARHGEF1 | Rho guanine nucleotide exchange factor 1 | No | N/A | N/A | N/A | N/A | N/A | N/A | signal transduction (GO:0007165; IBA) | hydrolase activity (GO:0016787; IDA) | Enzyme | N/A |
| ARL1 | ADP-ribosylation factor-like protein 1 | No | N/A | N/A | N/A | N/A | N/A | N/A | vesicle-mediated transport (GO:0016192; IMP; IBA) | hydrolase activity (GO:0016787; IDA) | Enzyme | N/A |
| BPI | Bactericidal permeability-increasing protein | No | N/A | N/A | N/A | N/A | N/A | N/A | regulation of leukocyte activation (GO:0002694; IDA; IBA) innate immune response (GO:0045087; IBA) | lipopolysaccharide binding (GO:0001530; IDA; IBA) lipid binding (GO:0008289; IDA; IBA) | Granule protein | Avoiding immune destruction |
| C11orf54 | Ester hydrolase C11orf54 | No | N/A | N/A | N/A | N/A | N/A | N/A | N/A | hydrolase activity (GO:0016787; IDA; IBA) | Enzyme | N/A |
| CAB39 | Calcium-binding protein 39 | No | N/A | N/A | N/A | N/A | N/A | N/A | signal transduction (GO:0007165; IDA; IBA) | kinase activator activity (GO:0019209; IDA; IBA) | Signal transducer/modulator | N/A |
| COPG1 | Coatomer subunit gamma-1 | No | N/A | N/A | N/A | N/A | N/A | N/A | vesicle-mediated transport (GO:0016192; IBA) | N/A | Vesicle trafficking regulator | N/A |
| DNPH1 | 2'-deoxynucleoside 5'-phosphate N-hydrolase 1 | No | N/A | N/A | N/A | N/A | N/A | N/A | epithelial cell differentiation (GO:0030855; IEP) | hydrolase activity (GO:0016787; IDA; IBA; ISS) | Enzyme | Unlocking phenotypic plasticity |
| EFHD2 | EF-hand domain-containing protein D2 | No | N/A | N/A | N/A | N/A | N/A | N/A | cadherin binding (GO:0045296; HAD) | cell adhesion molecule binding (GO:0050839; HDA) | Structural protein | N/A |
| NIT1 | Deaminated glutathione amidase | No | N/A | N/A | N/A | N/A | N/A | N/A | amide catabolic process (GO:0043605; ISS) | hydrolase activity (GO:0016787; ISS) | Enzyme | N/A |
| PCBD1 | Pterin-4-alpha-carbinolamine dehydratase | No | N/A | N/A | N/A | N/A | N/A | N/A | N/A | lyase activity (GO:0016829; IBA) | Enzyme | N/A |
| PITPNB | Phosphatidylinositol transfer protein beta isoform | No | N/A | N/A | N/A | N/A | N/A | N/A | vesicle-mediated transport (GO:0016192; IMP) | transporter activity (GO:0005215; IMP; IDA; IBA) | Vesicle trafficking regulator | N/A |
| POR | NADPH--cytochrome P450 reductase | No | N/A | N/A | N/A | N/A | N/A | N/A | response to hormone (GO:0009725; IBA) | oxidoreductase activity (GO:0016491; IDA; IBA) | Enzyme | N/A |
| PSMB3 | Proteasome subunit beta type-3 | No | N/A | N/A | N/A | N/A | N/A | N/A | proteasome-mediated ubiquitin-dependent protein catabolic process (GO:0043161; IBA) | N/A | N/A | N/A |
| RETN | Resistin | No | N/A | N/A | N/A | N/A | N/A | N/A | N/A | N/A | N/A | N/A |
| RP2 | Protein XRP2 | No | N/A | N/A | N/A | N/A | N/A | N/A | vesicle-mediated transport (GO:0016192; IGI; IMP; IBA) | GTPase regulator activity (GO:0030695; IDA; IBA) | Enzyme | N/A |
| RPL23 | 60S ribosomal protein L23 | No | N/A | N/A | N/A | N/A | N/A | N/A | positive regulation of cell population proliferation (GO:0008284; IMP) regulation of cell cycle (GO:0051726; IMP) | RNA binding (GO:0003723; HDA; IBA) transcription factor binding (GO:0008134; IPI) | Transcriptional/Translational regulator | Sustaining proliferative signaling |
| SNRPD2 | Small nuclear ribonucleoprotein Sm D2 | No | N/A | N/A | N/A | N/A | N/A | N/A | RNA processing (GO:0006396; IDA) | RNA binding (GO:0003723; HDA) | Transcriptional/Translational regulator | N/A |
| SNRPD3 | Small nuclear ribonucleoprotein Sm D3 | No | N/A | N/A | N/A | N/A | N/A | N/A | RNA processing (GO:0006396; IDA) | RNA binding (GO:0003723; HAD; IPI; ISS; IBA) | Transcriptional/Translational regulator | N/A |
| TPMT | Thiopurine S-methyltransferase | No | N/A | N/A | N/A | N/A | N/A | N/A | cellular response to xenobiotic stimulus (GO:0071466; IDA) | transferase activity (GO:0016740; IDA; IBA) | Enzyme | N/A |
| WBP2 | WW domain-binding protein 2 | No | N/A | N/A | N/A | N/A | N/A | N/A | response to hormone (GO:0009725; IDA; IMP) | transcription regulator activity (GO:0140110; IMP; IDA; IBA) | Transcriptional/Translational regulator | N/A |

Supplementary Table S3. Association of pancreatic cyst fluid protein signature with pancreatic cancer based on literature and Gene Ontology database curation. The association of the 89 differentially expressed proteins (DEP) signature identified in pancreatic cyst fluid (PCyF) was assessed by literature and database curation. For literature curation, association of 89 DEPs with pancreatic cancer (PC) was based on published literature found on PubMed database and assessed using information about gene and protein expression, along with cell type, process and signaling pathway each DEP is involved (Materials and methods). The PMID or DOI for each study used during literature curation are also mentioned. For database curation, the gene ontology (GO) database[26] terms for biological process and molecular function were used to assign protein family and cancer hallmark for each DEP. Identifiers (IDs) for each GO term are mentioned in brackets. Results from animal studies are mentioned in *italics*. Abbreviations: Extracellular vesicles: EVs; Intraductal papillary mucinous neoplasm: IPMN; Pancreatic intraepithelial neoplasia: PanIN; Pancreatic neuroendocrine tumor: PanNET; EXP: Inferred from Experiment; IDA: Inferred from Direct Assay; IPI: Inferred from Physical Interaction; IMP: Inferred from Mutant Phenotype; IGI:Inferred from Genetic Interaction; IEP: Inferred from Expression Pattern; ISS: Inferred from Sequence or structural Similarity; IBA: Inferred from Biological aspect of Ancestor; PMID: PubMed unique identifier; DOI: Digital Object Identifier.
